# High daily dose Short COurse PrimaquinE after G6PD testing for the radical cure of *Plasmodium vivax* malaria in Indonesia and Papua New Guinea: The SCOPE implementation study protocol

**DOI:** 10.1101/2024.12.15.24318165

**Authors:** Jeanne Rini Poespoprodjo, Moses Laman, Ayodhia Pitaloka Pasaribu, Inge Sutanto, Erni Nelwan, Liony Fransisca, Enny Kenangalem, Faustina Helena Burdam, Vincent Jimanto, Framita Ainur, Azkarunia Pasidiaz Hutagalung, Sherley Angeline, Adela Putri, Ari Winasti Satyagraha, Minerva Theodora, William Pomat, Maria Ome-Kaius, Mary Malai, Cynthia Abegini, Irene Pukai, Sharol Ronkentuo, Yolyne Amdara, Leo Makita, Evelien Rosens, Paul Daly, Rachael Farquhar, Nicholas M. Douglas, Kylie Mannion, Grad Dip, Ella Curry, Annisa Rahmalia, Vanessa S Sakalidis, Jacklyn Adella, Grant Lee, Benedikt Ley, Angela Devine, Patrick Abraham, Julie A. Simpson, Katelyn Brown, Thy Do, Heike Huegel, Helen Demarest, Elodie Jambert, Tanyaporn Wansom, Stephan Duparc, Leanne J. Robinson, Ric N. Price

## Abstract

**Introduction:** *Plasmodium vivax* malaria remains an important threat to the public in the Asia Pacific region. Preventing *P. vivax* relapses is crucial for reducing morbidity from malaria and ultimately controlling and eliminating this species. Primaquine is the only widely available drug with antirelapse activity against dormant stages of *P. vivax.* Its widespread use in clinical practice is limited by its potential to cause severe haemolysis in patients with glucose-6-phosphate dehydrogenase (G6PD) deficiency.

**Methods and analysis:** The primary aims of this staged, binational, multicentre, before-and-after implementation study are to determine the safety, feasibility, and cost-effectiveness of a revised package of case management interventions for improved *P. vivax* radical cure. The interventions include: i) pre-treatment testing of patients for G6PD deficiency using a semi-quantitative point-of-care device from SDBiosensor (ROK); ii) prescription of high dose primaquine (7mg/kg total dose) either over 7 days for G6PD normal patients (≥70% activity) or 14 days for intermediate patients (30-<70% activity), or lower dose weekly primaquine over 8 weeks for deficient patients (<30% activity); iii) improved patient education processes; iv) routine community-based review on day 3 (and day 7 for Stage 1) and v) enhanced malariometric surveillance and community pharmacovigilance. Stage 1 of the study (800 patients) will be implemented at 4 community clinics across Indonesia and Papua New Guinea (PNG) and will focus on analysis of treatment safety. If safety of the intervention is confirmed during Stage 1, the study will proceed to Stage 2, in which patient recruitment will be expanded to 10 clinics across Indonesia and PNG, and the feasibility of the similar intervention package will be assessed, but with a single community-based review on day 3. Stage 2 will run for 12 months and recruit approximately 11,410 patients. Mixed methods analyses of Stage 2 data will focus on the operational feasibility and cost-effectiveness of the revised case management package, with effectiveness determined through analysis of individual-level risk of *P. vivax* recurrence and population-level changes in incidence (with comparison to the pre-implementation period). Feasibility will be assessed via qualitative observations, in-depth interviews and focus groups of health care workers and participants.

**Discussion:** The intervention package will provide critical information on the safety, feasibility and cost-effectiveness of achieving radical cure with G6PD testing prior to high dose primaquine treatment and community-based follow-up. The study results will inform national malaria programs aiming to eliminate *P. vivax* in Indonesia and PNG by 2030.

**Ethics and dissemination:** This study has been approved by the World Health Organization Ethics Committee (Indonesia: ERC 0003810, PNG: ERC 0003892); Menzies School of Health Research (HREC: 2023-4524), Alfred Health (Burnet Institute) (Project No: 18/23), University of Gadjah Mada (KE/FK/0079/EC/2023), University of Indonesia (KET-347/UN2.F1/ETIK/PPM.00.02/2023), University of Sumatra Utara (80/KEPK/USU/2023), Institute of Tropical Medicine (1655/23), PNG Institute of Medical Research (PNGIMR) (IRB: 22.02), and the PNG National Department of Health Medical Research Advisory Committee (MRAC: 22.66). The study was registered on clinicaltrials.gov for Indonesia: NCT05879224 and PNG: NCT05874271. All participants will provide written, informed consent to join the study. For qualitative observations of health workers, a waiver of consent was granted by all ethics committees, and health centres will be made aware that observations on staff will be carried out. Dissemination plans include presentations at scientific conferences, peer-reviewed publications, and reporting at the national and community level.

**Strengths and limitations of this Study:**

- The staged design of this study will enable confirmation of the safety of high daily dose primaquine and provide operational insights on G6PD testing and community-based reviews, thereby facilitating the adoption of optimal practices in preparation for wide scale roll-out.
- Stage 2 will follow real-world conditions with treatment and follow-up embedded within local clinic procedures and implemented by clinic healthcare staff and community-based health workers. These efforts will enable a realistic assessment of the interventions’ feasibility, cost-effectiveness, and scalability.
- Study sites in Indonesia and PNG are publicly funded facilities servicing populations from areas with markedly different malaria endemicity and workloads, supporting the generalisability of our results to regions across both countries with varying malaria epidemiology.
- Implementation of the intervention may be limited by the informed consent process, potentially restricting enrolment and coverage of the intervention across the study clinics.
- Multiple time-varying confounders, such as government-initiated malaria control activities, may influence malaria incidence and, therefore, impair the ability to determine the true population-level impact of the study interventions.

## BACKGROUND

*Plasmodium vivax* has become the predominant cause of malaria in the Asia-Pacific region (1). *P. vivax* is more difficult to eliminate than *P. falciparum* because it forms dormant liver stages (hypnozoites) that can reactivate weeks to months after the initial infection, resulting in recurrent blood stage infections, known as relapses. In some endemic regions, approximately 80% of *P. vivax* cases are caused by relapses (2). In 2022, a total of 101,300 cases of vivax malaria were reported in Indonesia and 211,000 cases in Papua New Guinea (PNG), with the most significant burden occurring in remote and rural areas (3). The governments of both countries have committed to eliminating *P. vivax* by 2030; accomplishing this ambitious goal will require innovative strategies to ensure the widespread availability of well-tolerated and effective radical cure of individuals harbouring blood and/or liver stages of the parasite.

The 8-aminoquinoline compounds, primaquine and tafenoquine, are the only licensed antimalarial drugs that kill hypnozoites and thus can prevent subsequent relapses of *P. vivax*. Both drugs can cause severe haemolysis in patients with glucose-6-phosphate dehydrogenase (G6PD) deficiency (4, 5). Tafenoquine was approved by the US and Australian health authorities in 2018 and is given as a single dose, facilitating patient adherence (6–8). Currently, tafenoquine can only be prescribed in combination with chloroquine to patients with >70% G6PD enzymatic activity, making access to this drug limited in the many endemic countries where routine G6PD testing is unavailable (9). Primaquine has been the primary treatment for preventing *P. vivax* relapses for almost 70 years and is used in combination with chloroquine and artemisinin-based combination therapies (ACTs) (5). Primaquine is widely available, but its population-level effectiveness is confounded by suboptimal dosing, poor adherence of patients to prolonged treatment courses (10) and the reluctance of healthcare providers to prescribe the drug due to concerns of severe haemolysis in G6PD-deficient patients (11).

The World Health Organization (WHO) antimalarial treatment guidelines currently recommend a low-dose primaquine regimen (total dose of 3.5mg/kg administered as 0.25mg/kg/day over 14 days or 0.5 mg/kg/day over 7 days) (12). The 14-day regimen is most commonly used in *P. vivax*-endemic countries (13). It is recommended that all patients are tested for G6PD deficiency before administration of primaquine. If an individual is diagnosed with G6PD deficiency (<30% enzyme activity), then an 8-week regimen of 0.75mg/kg primaquine administered once per week is recommended (12). In PNG, the national policy for radical cure is primaquine after pre-treatment testing for G6PD deficiency, combined with blood schizonticidal treatment with artemether-lumefantrine (14). The same radical cure regimen in Indonesia is combined with dihydroartemisinin-piperaquine (15). In practice, G6PD testing is rarely available and hence the national malaria control programmes (NMCPs) recommend a low daily dose of primaquine (0.25 mg/kg/day) administered over 14 days to reduce the risk of drug-induced haemolysis (11, 13). Adherence to this prolonged treatment course is typically poor when unsupervised (16, 17). Partial supervision, conversely, has been shown to improve adherence and anti-relapse effectiveness (10, 18, 19).

The antirelapse efficacy of primaquine is related to the total dose administered, whereas safety and tolerability are related to the daily dose administered (20). A recent meta-analysis of the antirelapse efficacy of primaquine demonstrated that the overall risk of recurrent *P. vivax* within 6 months was 51.0% (95% confidence interval (CI) 48.2–53.9%) in patients who were not treated with primaquine, 19.3% (95%CI: 15.9-21.9%) in patients treated with low-dose primaquine (∼3.5mg/kg total dose), and 8.1% (95% CI: 7.0-9.4%) for patients treated with high-dose primaquine (∼7mg/kg total dose). The benefits of the higher total dose regimen were apparent in both low and high relapse periodicity areas (21).

Reducing the duration of primaquine treatment has the potential to improve adherence, but requires the same total dose to be administered over a shorter period, thereby increasing the daily dose needed and the subsequent risk of drug induced haemolysis for patients. Two recent clinical trials conducted in Thailand, Afghanistan, Indonesia, Ethiopia and Vietnam (22, 23) have demonstrated that a short-course high daily dose of primaquine (1mg/kg/day over 7 days – PQ7) is non-inferior to the standard high dose regimen (0.5 mg/kg/day administered over 14 days with supervision – PQ14). However, PQ7 was associated with more adverse events, particularly gastrointestinal intolerability (22, 23). The latter can be mitigated by coadministration with food.

G6PD deficiency is an X-linked inherited trait. Whereas males are hemizygous normal or deficient, females can be homozygous normal, homozygous deficient or heterozygous for the G6PD gene (24). The latter is associated with a variable degree of intermediate deficiency and an increased risk of haemolysis (25, 26). A recent systematic review demonstrated that early indicators of significant haemolysis are typically apparent within 5 days of treatment commencement and usually preceed severe clinical compromise and the need for medical intervention (27). These findings suggest that routine clinical review within 3-5 days of starting primaquine may facilitate early detection of haemolysis so that treatment can be ceased and severe consequences of primaquine-induced haemolysis avoided (27).

Over the last decade a series of point-of-care tests for G6PD deficiency have been developed and marketed to screen for patients at risk of primaquine or tafenoquine-induced haemolysis (28–30). The SD Biosensor provides reliable differentiation of patients with deficient (<30%), intermediate (30-<70%) and normal (≥70%) G6PD activity within 2 minutes using one drop of capillary blood, therefore lending itself to pre-treatment testing.

In collaboration with Indonesian and PNG policymakers, the SCOPE study was devised to evaluate a revised and enhanced *P. vivax* case management protocol developed to improve access to well-tolerated and effective radical cure. The study is anticipated to inform Indonesian and PNG treatment guidelines and is relevant to other *P. vivax* endemic countries.

## METHODS AND ANALYSIS

### Introduction to Study Design and Setting

The SCOPE study is a pragmatic, staged, binational, multicentre, before-versus-after implementation study of a revised case management package aimed at optimising radical cure for patients with vivax malaria presenting to community health clinics in Indonesia and PNG (Figure 1). Ten publicly funded clinics (6 in Indonesia and 4 in PNG) servicing areas with widely disparate *P. vivax* incidence have been selected for the study (Figure 2). The annual caseloads range from 412 to 4,937 patients per annum and this spectrum of endemicity will support the generalisability of study results (Table 1). The revised case management protocol incorporates five novel components:

i. Pre-treatment testing of patients for G6PD deficiency using the semi-quantitative point-of-care SD Biosensor device.
ii. Prescription of high dose primaquine (7mg/kg total dose) over 7 days (G6PD normal patients - ≥70% activity, ≥6.1 U/gHb), or over 14 days (intermediate patients – 30-<70% activity, 4.1-6.0 U/gHb) or primaquine one a week for 8 weeks (deficient patients - <30% activity, ≤4.0 U/gHb) (Table 2).
iii. Improved patient education processes, including supervision of the first dose of primaquine and counselling regarding key risks and benefits of therapy as well as the necessity to take doses with food.
iv. Routine community-based review on day 3 (and day 7 for Stage 1) for the dual purpose of encouraging adherence to primaquine and detecting early warning signs of impending severe haemolysis and other adverse effects
v. Enhanced malariometric surveillance and community pharmacovigilance to support wider-scale use of the revised case management package

**Figure 1.**
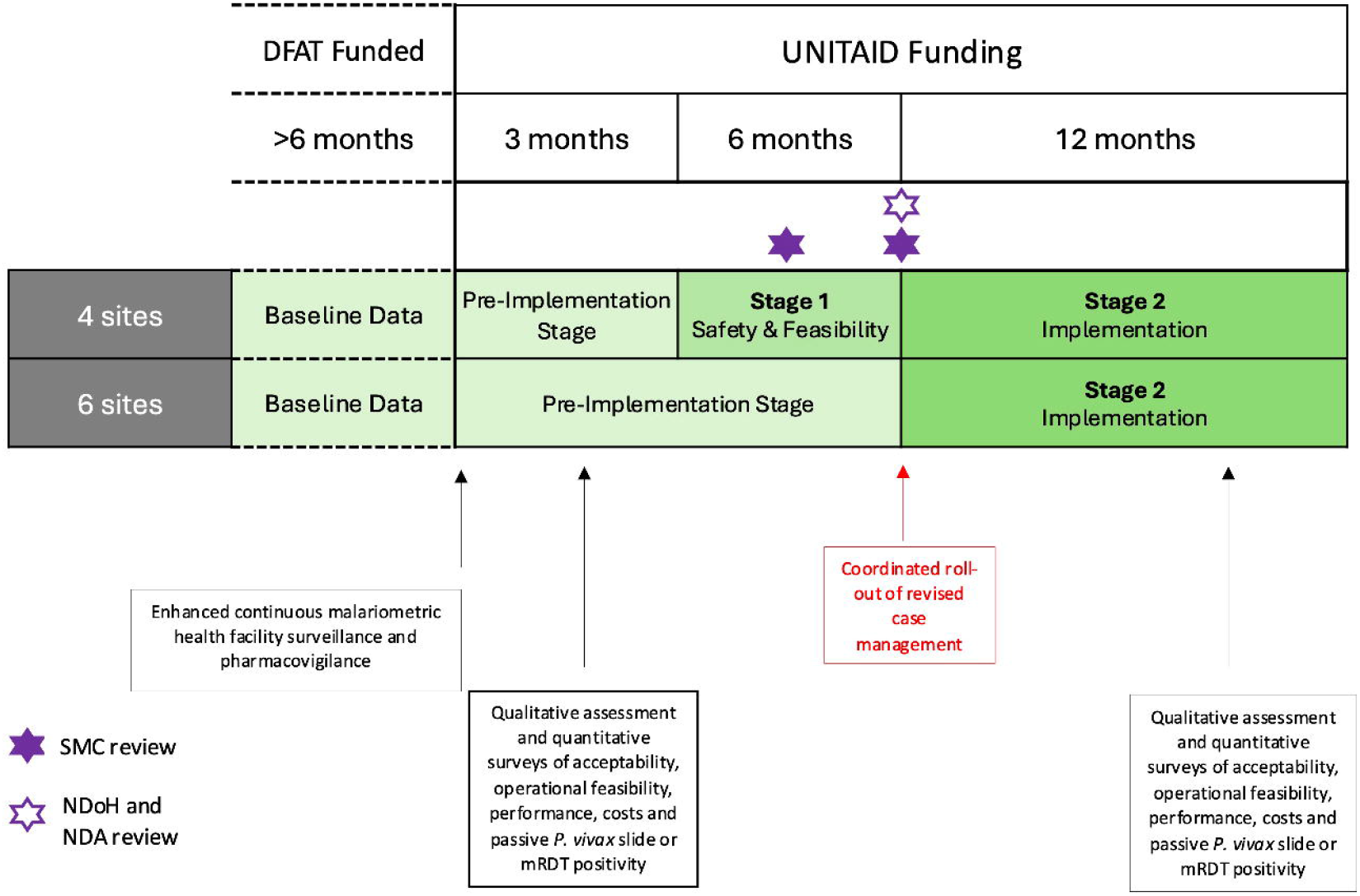
Study schedule.

**Figure 2.**
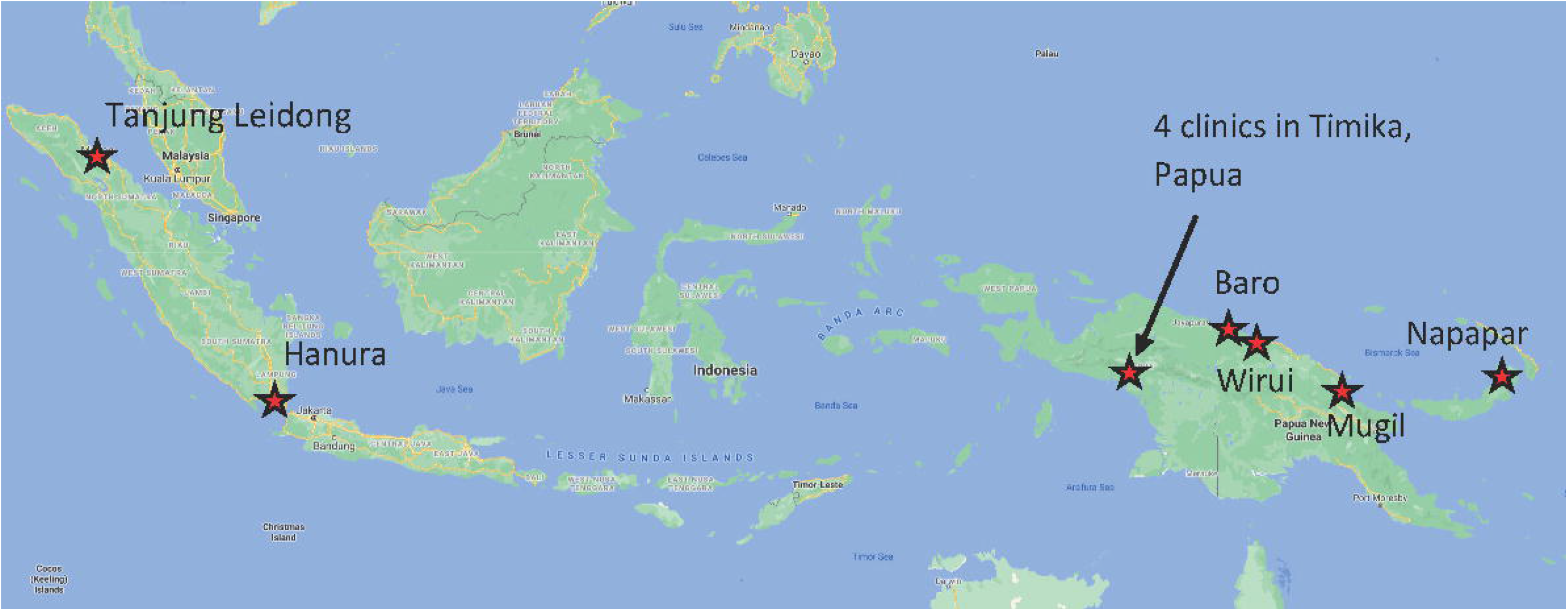
Location of study clinics.

**Table 1:**
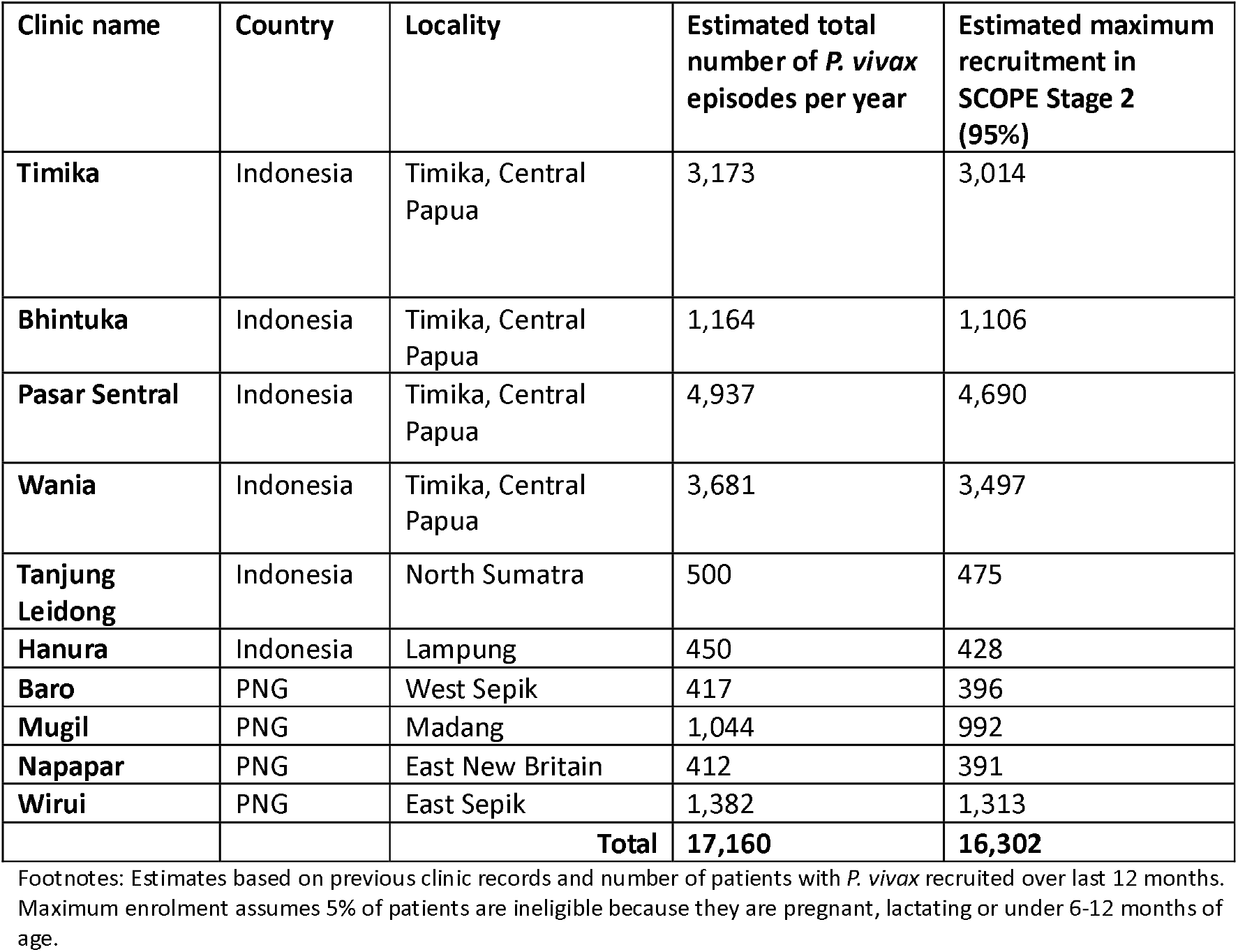
Study Clinics estimated *P. vivax* episodes per year.

| Clinic name | Country | Locality | Estimated total number of *P. vivax* episodes per year | Estimated maximum recruitment in SCOPE Stage 2 (95%) |
| --- | --- | --- | --- | --- |
| **Timika** | Indonesia | Timika, Central Papua | 3,173 | 3,014 |
| **Bhintuka** | Indonesia | Timika, Central Papua | 1,164 | 1,106 |
| **Pasar Sentral** | Indonesia | Timika, Central Papua | 4,937 | 4,690 |
| **Wania** | Indonesia | Timika, Central Papua | 3,681 | 3,497 |
| **Tanjung Leidong** | Indonesia | North Sumatra | 500 | 475 |
| **Hanura** | Indonesia | Lampung | 450 | 428 |
| **Baro** | PNG | West Sepik | 417 | 396 |
| **Mugil** | PNG | Madang | 1,044 | 992 |
| **Napapar** | PNG | East New Britain | 412 | 391 |
| **Wirui** | PNG | East Sepik | 1,382 | 1,313 |
|  |  | **Total** | **17,160** | **16,302** |

Footnotes: Estimates based on previous clinic records and number of patients with *P. vivax* recruited over last 12 months. Maximum enrolment assumes 5% of patients are ineligible because they are pregnant, lactating or under 6-12 months of age.

**Table 2.**
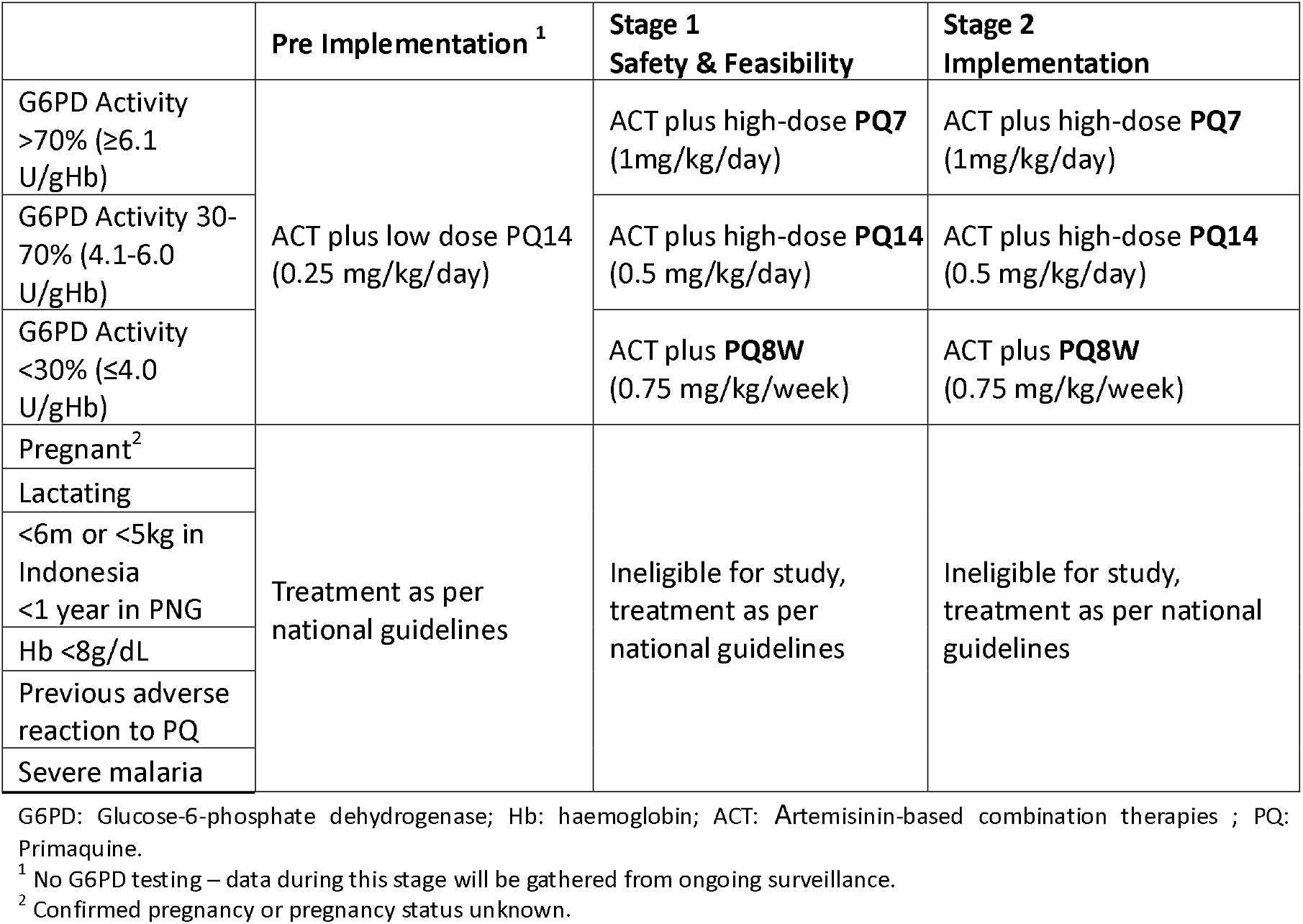
Summary of *P. vivax* patient eligibility and treatment in the different stages of the study.

The study uses a staged approach. Stage 1 aims to build confidence that short course, high dose primaquine can be implemented safely before progressing to Stage 2, in which a large-scale implementation study will be conducted of the revised case management protocol under conditions more closely matching real-world practice. The overarching aim of the study is to determine the safety, operational feasibility and cost-effectiveness of the revised case management protocol. The study will be conducted over 21 months, and the results will be reported in line with the CONSORT extension (31) and SPIRIT statement (32) (Supplementary data 1).

### Study components and data collection

#### Pre-implementation

The study will commence with a pre-implementation stage during which continuous malariometric surveillance at all 10 clinics will document clinic case numbers, building upon pre-existing surveillance data collection systems. Incidence data collected during the pre-implementation and Stage 1 recruitment periods (Figure 1) will provide a reference for comparing post-implementation incidence. Midway through the pre-implementation stage, 200 consecutive patients with fever presenting to each clinic will be recruited into a cross-sectional survey assessing the microscopy-based prevalence of *P. vivax* and G6PD activity (Table 3, Figure 3). The pre-implementation phase will last three months for the four clinics participating in Stage 1 of the study and 9 months for the remaining 6 clinics.

**Figure 3.**
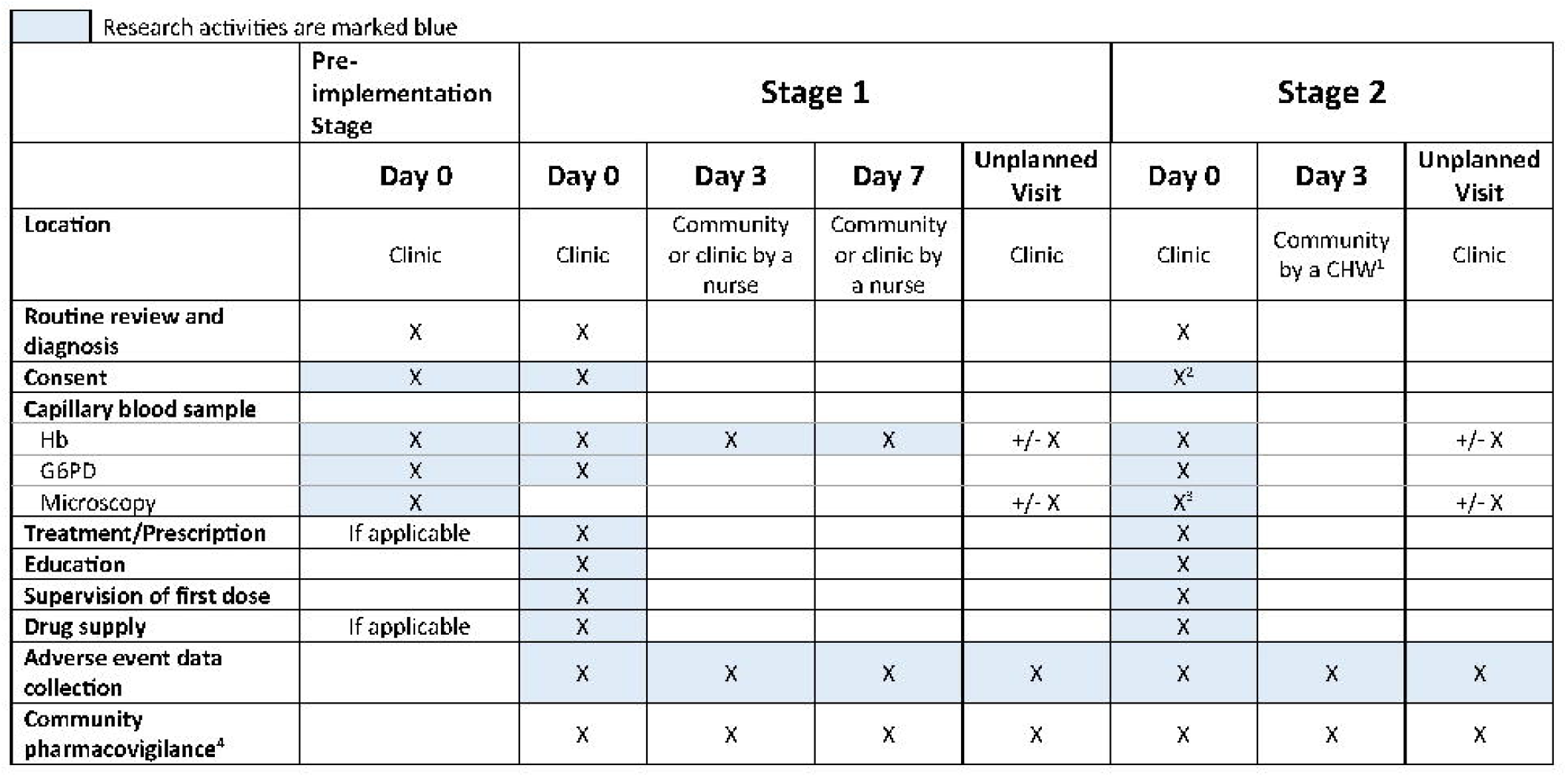
Schedule of enrolment, interventions, and assessments. ^1^Day 3 review may be a phone call if the CHW cannot access the patient at home, or at the clinic if the patient presents there. ^2^During Stage 2 200 consecutive febrile patients at each clinic will be consented for the post-implementation survey, which includes the following procedures: Hb, G6PD and Microscopy only. If positive for vivax malaria, patients will also be recruited to Stage 2. ^3^Microscopy will only be performed for 200 consecutive febrile patients at each clinic during post-implementation survey, not Stage 2. ^4^Any AESI/SAE identified during study participants’ primaquine treatment will be captured.

**Table 3.** Study Procedure and Data Collection in Pre-implementation Stage.

| Research Activity | Day of Collection | Study Procedures | Data Collection |
| --- | --- | --- | --- |
| Surveillance | Day 0 | Routine care only | Routine clinic data |
| Pre-implementation Survey | Day 0 | Written informed consent<br>Capillary blood sample for malaria diagnosis, G6PD and Hb | Malaria diagnosis, Hb and G6PD activity |
| Community Pharmacovigilance | Continuous | Passive detection of serious adverse events in patients presenting to clinic and hospital | Records of all serious adverse events (Supplementary Material 4) |
| Qualitative Interviews and Focus Groups | Continuous | Field testing In-depth interviews and focus group tools with key stakeholders<br>Observations of routine health care activities | Transcripts and summaries of interviews<br>Observations notes and memos |
| Health Economic Survey | Day 0 and Day 7 | Household cost survey per *P. vivax* episode at each clinic | Direct and indirect costs to patients and any caregivers |

G6PD: Glucose-6-phosphate dehydrogenase; Hb: haemoglobin.

#### Stage 1

Following three months of pre-implementation, Stage 1 of the SCOPE study will commence at two clinics in Papua, Indonesia and two in PNG, with a planned recruitment duration of 6-12 months. This study component aims to provide preliminary safety and feasibility data before progressing to Stage 2 at all ten clinics. A total of 800 patients with non-severe *P. vivax* mono- or mixed species infection diagnosed by microscopy or rapid diagnostic test will be enrolled (500 in Indonesia and 300 in PNG) and, following pre-treatment G6PD activity assessment using the SD Biosensor, patients will receive primaquine according to the dosing schedules outlined above (Table 2 and Supplementary Data 2). Community-based follow-up assessments by study nurses will be conducted on day 3 and day 7 of treatment (Supplementary Data 3; Level 2 Review Form), with G6PD activity and haemoglobin assessed on both days. Patients with severe adverse events (SAE) or adverse events of special interest (AESI) will be referred to the clinic for a health practitioner-led clinical review (Supplementary Data 3; Level 3 Review Form and SAE form) and referred to the hospital for medical management if appropriate. All patients will also receive enhanced education and counselling as per the study interventions specified previously (Table 4).

**Table 4.** Study Procedure and Data Collection in Stage 1.

| Research Activity | Day of Collection | Study Procedures | Data Collection |
| --- | --- | --- | --- |
| **Enrolment and Baseline** | **Day 0** | • Clinical review<br>• Capillary blood sample for malaria diagnosis by microscopy or rapid diagnostic test (clinic procedure)<br>• Written informed consent<br>• If malaria positive, a 2<sup>nd</sup> capillary blood sample for G6PD and Hb measurement<br>• Treatment prescribed<br>• Education package<br>• Supervision of first dose<br>• Supply of drugs | • Routine clinic data<br>• Day 0 Hb and G6PD activity, malaria diagnosis, PQ treatment |
| **Level 2 Review** | **Day 3 and 7** | • Capillary blood sample for Hb measurement<br>• Signs and symptoms questionnaire by research nurse | • Day 3 signs and symptoms<br>• Day 7 signs and symptoms<br>• Day 3 Hb<br>• Day 7 Hb<br>• SAEs and AESIs |
| **Level 3 Review** | **When referred from Level 2** | • Capillary blood sample Hb measurement<br>• Signs and symptoms questionnaire by medical practitioner | • Hb, MetHb<br>• Signs and symptoms<br>• SAEs and AESIs |
| **SAE Form** | **Hospitalisation or death** | • Capillary blood sample Hb measurement<br>• Other laboratory tests<br>• Clinical management<br>• Classification of SAE | • Hb, other laboratory tests<br>• Clinical management<br>• SAE classification |
| **Community Pharmacovigilance** | **Continuous** | • Passive detection of SAEs and AESIs | • All SAEs and AESIs |
| **Qualitative** | **Continuous** | • IDI, focus groups with patients, key stakeholders and community members of acceptability of revised case management strategy | • Transcripts and summaries of interviews<br>• Observations notes and memos |

G6PD: Glucose-6-phosphate dehydrogenase; Hb: haemoglobin; MetHb: Methaemoglobin SAE: serious adverse event; AESI: adverse event of special interest; IDI: in-depth interview.

#### Stage 2

The Stage 2 implementation study will run for 12 months and will recruit all eligible patients with *P. vivax* mono- or mixed species infection presenting to one of 10 study clinics across Indonesia and PNG. Conditions during this stage are designed to emulate real-world practice as closely as possible. Stage 2 will follow the same study procedures as Stage 1, except that clinic (rather than research) staff will perform consent procedures and G6PD testing, and follow-up will be reduced to a single clinical review (either in person or by phone) on day 3 (Supplementary Data 3; Level 1 Review Form). The latter will be conducted by a Community-Based Health Worker (CbHW) or village midwife rather than a research nurse. Patients with symptoms or signs of AESIs or SAEs will be referred to the clinic for a Level 2 or Level 3 clinical review (Supplementary data 3; Level 2 and Level 3 Review Form). The subsequent review and management of the patient will be the same as in Stage 1, with referral to the site medical practitioner as necessary. In Stage 2, patients re-presenting with vivax malaria will be eligible for repeat enrolment (Table 5).

**Table 5.** Procedure and Clinical Data Collection in Stage 2.

|  | Day of Collection | Study Procedures | Data Collection |
| --- | --- | --- | --- |
| **Enrolment and baseline** | Day 0 | • Clinical review<br>• Capillary blood for malaria diagnosis by microscopy or rapid diagnostic test (clinic procedure)<br>• Written informed consent<br>• If malaria positive, a 2<sup>nd</sup> capillary sample for G6PD and Hb measurement<br>• Treatment prescribed<br>• Education package<br>• Supervision of first dose of DHP and PQ | • Routine clinic data<br>• Day 0 Hb and G6PD activity |
|  |  | • |  |
| **Level 1 Review** | Day 3 | • Symptom questionnaire by CbHW | • Patient follow-up<br>• Symptoms and signs of patients referred to clinic<br>• SAEs and AESIs |
| **Level 2 Review** | When referred from Level 1 review or patient presenting to health facility | • Capillary blood sample for Hb measurement<br>• Signs and symptoms questionnaire by clinic nurse | • Day 3 signs and symptoms<br>• Day 3 Hb<br>• SAEs and AESIs |
| **Level 3 Review** | When referred from Level 2 | • Capillary blood sample Hb measurement<br>• Signs and symptoms questionnaire by medical practitioner | • Hb, Met Hb<br>• Signs and symptoms<br>• SAEs and AESIs |
| **Community Pharmacovigilance** | Continuous | • Passive detection of SAEs and AESIs | • All SAEs and AESIs |
| **Qualitative Interviews, focus group discussions (FGDs), Photovoice and observations** | Continuous | • IDI, focus groups with key stakeholders and community members of acceptability of revised case management strategy<br>• Photovoice (PNG only) with community members of revised case management strategy | • Transcripts and summaries of interviews and FGDs<br>• Observations notes and memos<br>• Photos, summaries of transcripts from photovoice FGDs. |
| **Health Economic Survey** | Day 0 and Day 7 for household costs<br>Continuous for provider costs | • Household cost survey per *P. vivax* episode at each clinic<br>• Ingredients-based provider costs at selected clinics | • Direct and indirect costs to patients and any caregivers<br>• Provider costs for each component of the revised case management strategy |
| **Post-implementation Survey** | 6 months into Stage 2 | • Written informed consent<br>• Capillary blood sample for malaria diagnosis, G6PD and Hb | • Malaria diagnosis, Hb and G6PD activity |

G6PD: Glucose-6-phosphate dehydrogenase; Hb: haemoglobin; MetHb: Methaemoglobin; SAE: serious adverse event; AESI: adverse event of special interest; IDI: in-depth interview; CbHW: community-based health worker.

### Health Economics

Detailed cost questionnaires will be completed by 30 consecutive patients (or their guardians) presenting with *P. vivax* malaria at each clinic (total of 300 patients) to determine the direct and indirect costs to households. In Indonesia, this will be completed during the pre-implementation stage and the final 6 months of Stage 2. In PNG, only one data collection timepoint per clinic will be completed. Data will also be collected on the costs of community engagement meetings, patient education, clinical reviews, pharmacovigilance, staff training, serious adverse events (where relevant) and malaria surveillance.

### Social Science

During pre-implementation, purposive sampling of key stakeholders will be used to identify participants for in-depth interviews to determine the acceptability and operational feasibility of G6PD testing. Focus group discussions and in-depth interviews with malaria program officers at the clinics, provincial and district health office representatives and health care providers will be held during Stage 1 and Stage 2 to gather information on the practicality of the revised case management, challenges for implementation, and inefficiencies. Community-based health workers will also be involved in focus group discussions. In PNG, a subset of community members that test positive with *P. vivax* will be invited to be involved photovoice, to critically reflect on the clinical reviews and PQ adherence during day 3 and day 7 follow ups. Participant observations, informal conversations and community engagement methods will be undertaken at the health clinics and certain sites in the villages at several points during Stage 1 and Stage 2, across both PNG and Indonesia. The number of participants involved in the social science research will vary by site, but the aim will be to reach a broad spectrum of demographics, including participants of different ethnicity, sex and age (Table 6).

**Table 6.**
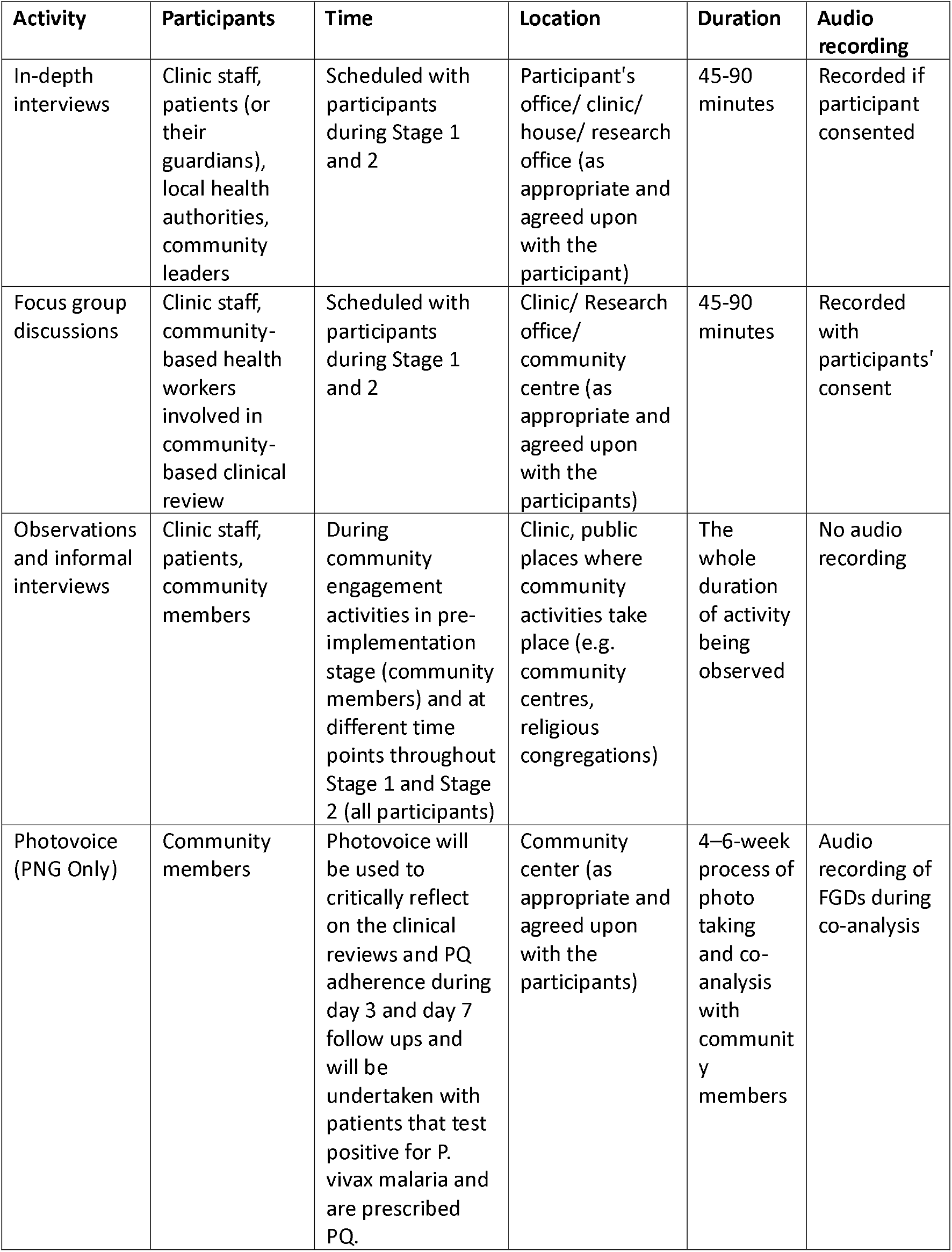
Qualitative research activities.

### Study objectives and related endpoints

The overall objective of the SCOPE study is to determine the safety, feasibility and cost-effectiveness of high-dose primaquine after point of care G6PD testing. The study has additional subsidiary objectives with associated endpoints, subdivided by study stage and objective category addressing operational feasibility, cost-effectiveness, safety and pharmacovigilance (Table 7). The two primary endpoints in Stage 1 include the proportion of patients experiencing at least one SAE during treatment and the proportion of patients experiencing at least one AESI during treatment. In Stage 2 the primary endpoint is the proportion of patients with *P. vivax* malaria who correctly receive all components of revised case management (including G6PD testing, treatment with PQ according to the revised case management protocol and G6PD activity, patient education, supervision of the first dose of PQ and community review on day 3).

**Table 7.**
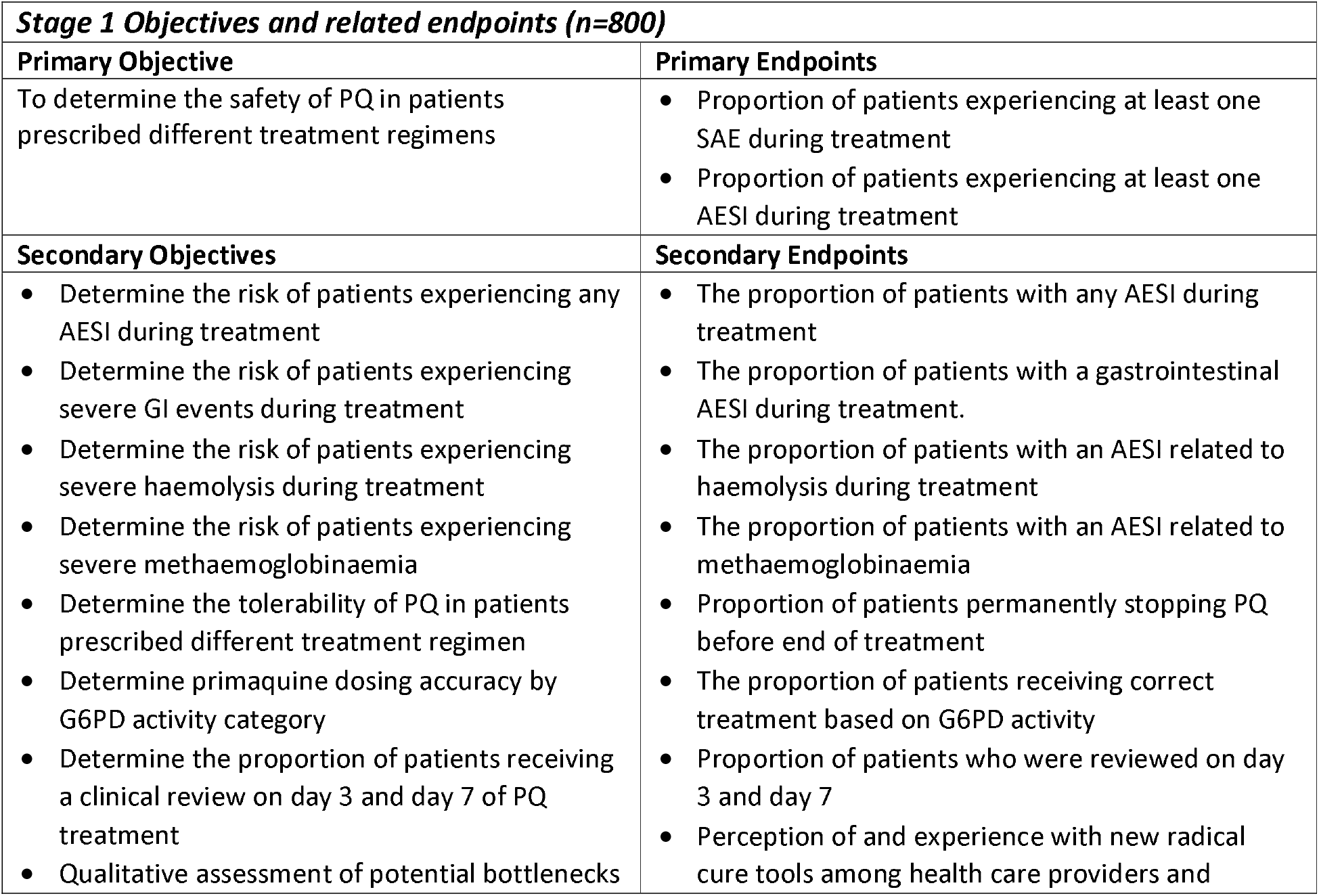
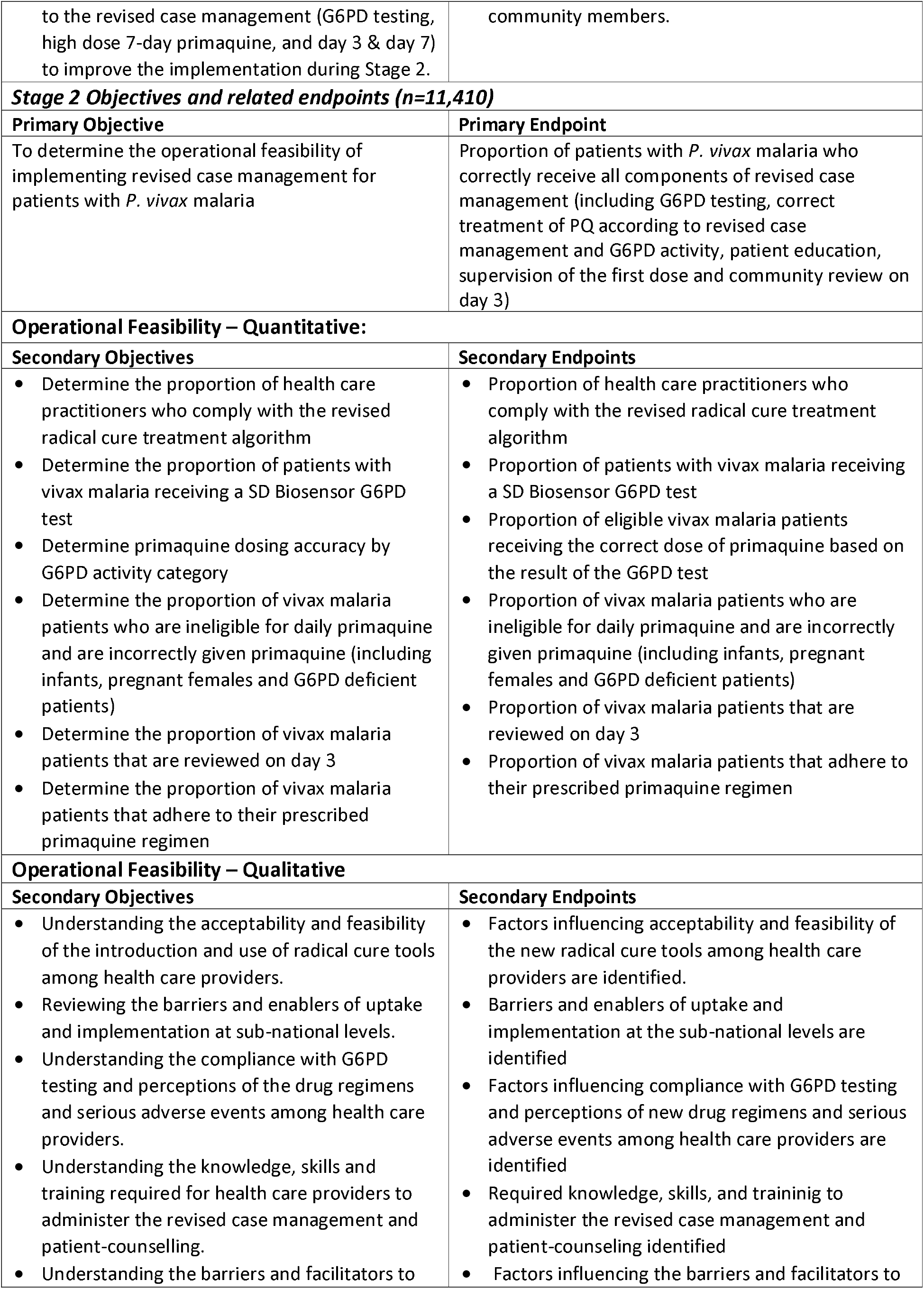
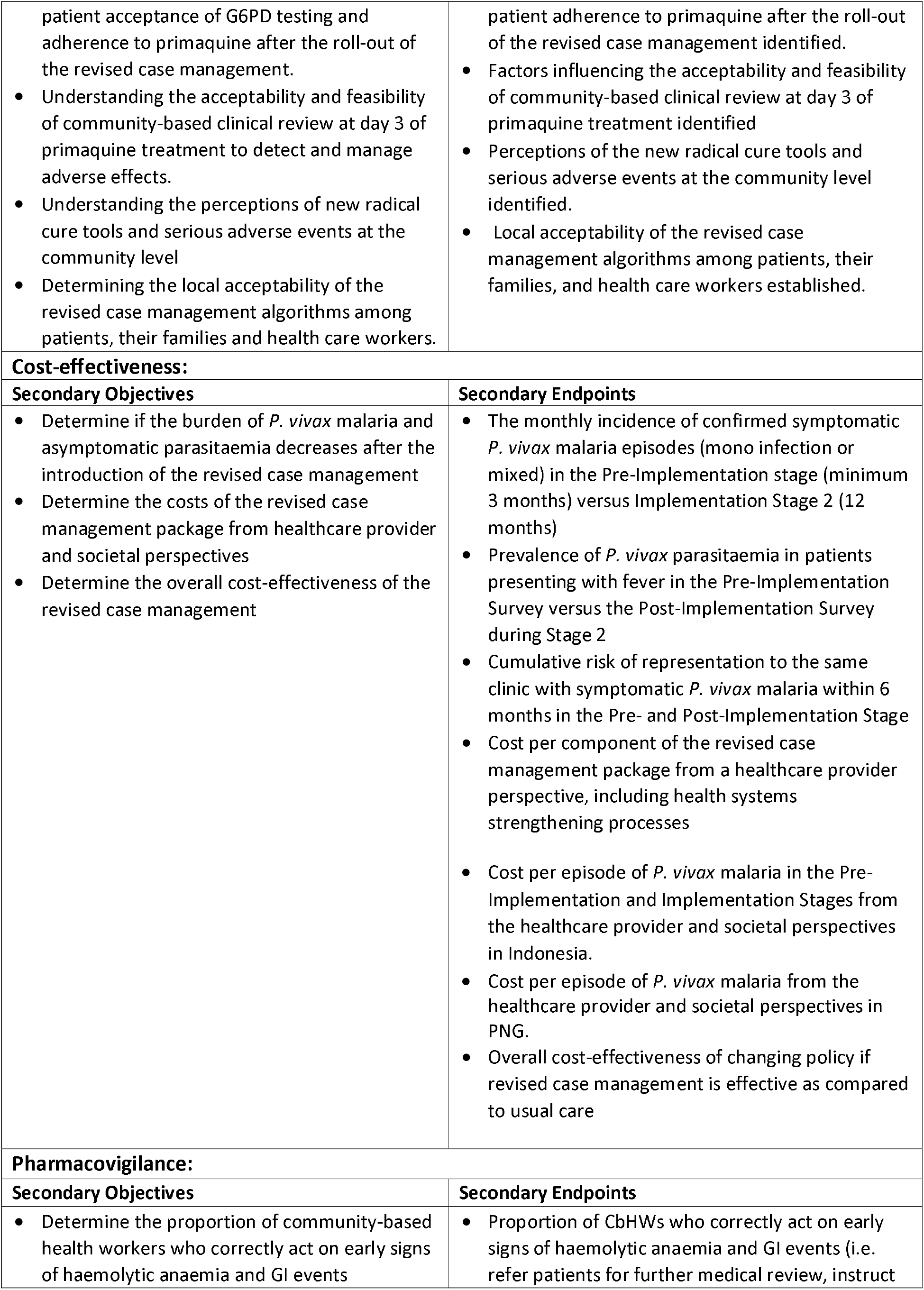
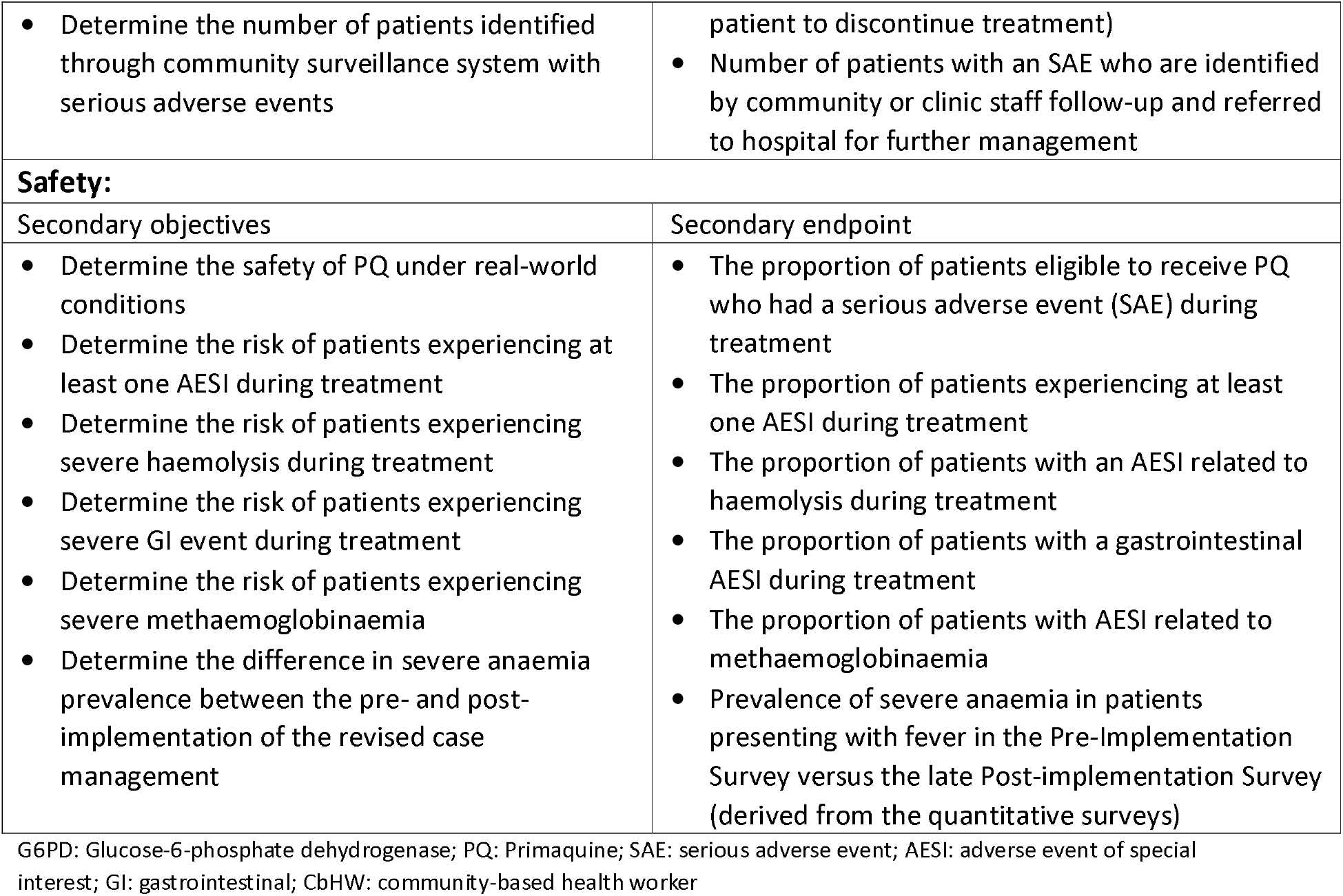
Study objectives and endpoints.

### Study participant selection and recruitment

Criteria for participation in the SCOPE study differ between the stages of the study (Table 2). To be included in the pre or post-implementation cross-sectional prevalence surveys, patients must have presented to a study clinic with fever or history of fever within the last 48 hours and must consent to the study procedures, including providing a finger-prick blood sample. For inclusion in Stage 1 or Stage 2, patients must be >6 months of age and >5kg in weight (Indonesia) and >1 year of age in PNG with microscopically or RDT-confirmed *P. vivax* mono- or mixed species infection. Patients will be excluded from the Stage 1 and Stage 2 studies (but not routine malariometric surveillance data collection) if they have a haemoglobin <8g/dL, are pregnant or breastfeeding an infant <6 months of age in Indonesia or <12 months of age in PNG, have had a previous adverse reaction to primaquine, have evidence of severe malaria or need hospital admission or referral. Participants in the various qualitative components of the study will be selected purposively to ensure adequate demographic distribution and representation from key stakeholders (Table 8). Patients with vivax malaria who participate in the SCOPE study will receive a unique identification number along with a brief malaria-related medical summary. Patients will be asked to bring their unique number with them for any future appointments to enable linkage of repeated episodes.

**Table 8.**
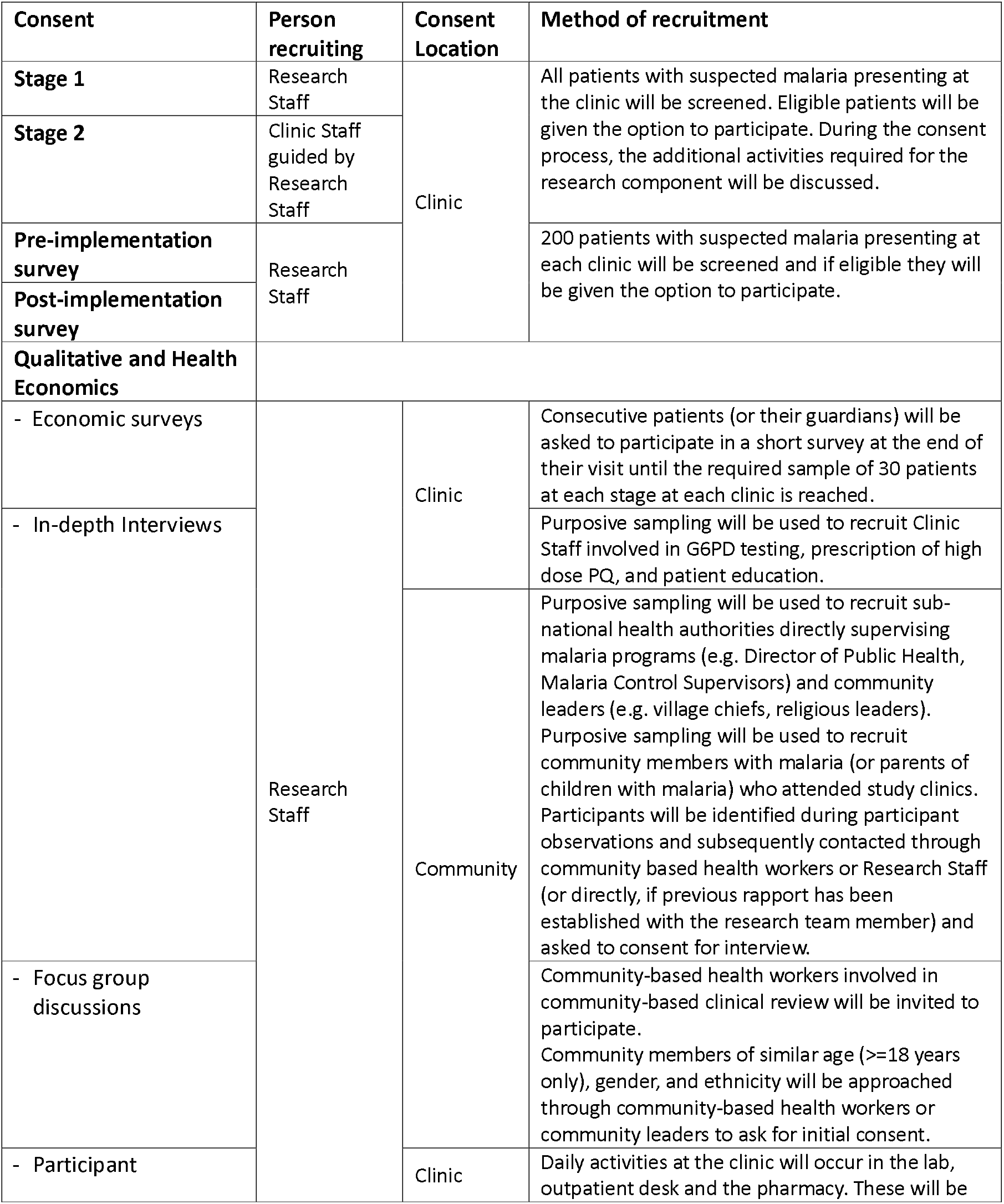
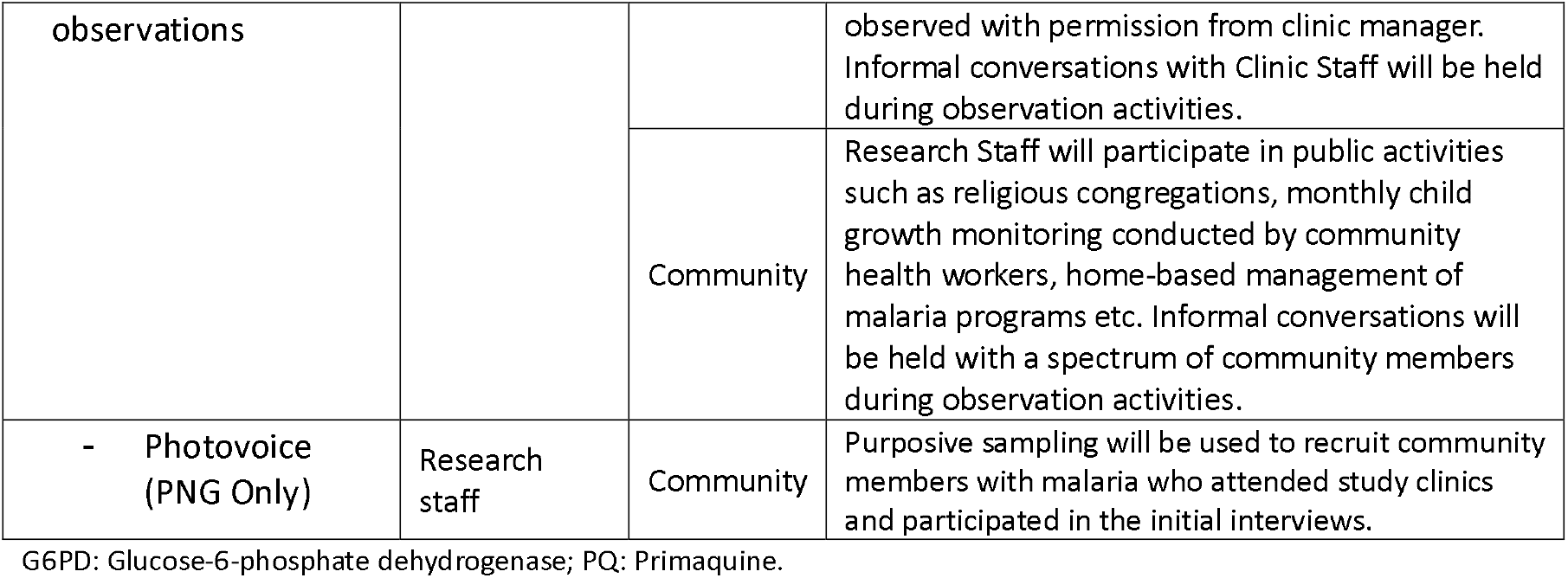
Participant recruitment and staff involved.

### Participant Consent

Individual written informed consent collected by trained researchers and clinic healthcare workers will be obtained from participants (or their guardians in the case of minors) in the pre-and post-implementation cross-sectional surveys, Stages 1 and 2, economic surveys, in-depth interviews, focus group discussions and photovoice. In Indonesia, minors over the age of 11 years will be asked to sign an assent form in addition to full written consent provided by their legal guardian. All patient information and the consent documents will be provided in the relevant local language. Provisions have been made for participants who are illiterate to have the consent documents read to them and their written consent recorded via a “mark” witnessed by a third party independent of the study team. Patients declining to participate in Stages 1 or 2 will receive malaria care as per national guidelines. Participants will be told that they can withdraw consent at any time without risk of prejudice, and only their data up to the time of withdrawal will be retained. Participants will remain in the study for the duration of their treatment, and their completion of the study will be facilitated by the community-based reviews. For qualitative observations of health workers, a waiver of consent was granted by all ethics committees, and health centres will be made aware that observations on staff will be carried out.

### Patient and Public Involvement

Before the study commences, in-depth stakeholder engagement sessions will be held with clinic staff, community leaders, religious and secular leaders, and the Ministries of Health. Engaged participants will have the opportunity to provide feedback on the operating procedures and patient educational materials and where possible, their feedback will be incorporated into the study. Community awareness and engagement sessions in villages within the health facility catchment area will also be conducted with the support of community leaders and clinic staff.

After the study is completed, there will be a process of community engagement with clinic staff, community leaders and community members to describe the results of the study and to gather feedback on possible refinement of future vivax malaria management and mechanisms for expansion into routine practice. Findings will be discussed with representatives from the respective National Malaria Control Programmes with respect to the viability of the intervention in terms of safety, feasibility, and cost-effectiveness.

### Power and Sample Size

In Stage 1, 4 clinics will enrol 800 patients (500 in Indonesia and 300 in Papua New Guinea). Assuming 764 of these patients will have a G6PD activity >70% and be eligible for PQ7 and considering that the proportion of patients receiving short-course primaquine who have to stop therapy within 7 days is 5%, this sample size will produce a 95% confidence interval for the true proportion needing to stop treatment with a margin of error +/- 1.55%.

In Stage 2, the implementation study will be conducted at ten health facilities across Indonesia and Papua New Guinea. Based on recruitment during Stage 1 and prior surveillance, we estimate a total of 11,410 patients with vivax malaria will be eligible and recruited into Stage 2 in Indonesia and Papua New Guinea. Across the 10 clinics participating in Stage 2, a mean of 136 patients with vivax malaria are expected to be seen at each clinic per month, 60% of which (n=82) are predicted to be relapses and therefore preventable with improved primaquine therapy. Assuming the implementation package in Stage 2 will reduce the individual risk of relapse by 30%, the monthly incidence of vivax malaria is expected to fall by 17 cases (12.6%) per month. With a two-sided alpha of 0.05 and a predicted standard deviation of the mean variation in monthly numbers of vivax malaria cases at each clinic of 15, our sample size would achieve 89% power to detect this difference in incidence.

The risk of recurrence will also be assessed at an individual level using comparisons with patients recruited during the pre-implementation phase. Based on a presumptive sample size of 11,410 patients matched 1:1 with 11,410 patients with vivax malaria in the pre-implementation phase, our sample size would have 100% power to detect a 30% reduction in individual risk of recurrence at 6 months and 92% power to detect a 10% reduction, assuming 10% loss to follow-up in each group and a baseline risk of recurrence at 6 months of 20%.

If 70% of patients recruited into Stage 2 receive all components of the implementation package correctly, a sample size of 11,410 will give a 95% confidence interval for the true proportion of +/- 0.84%. If 1% of patients are predicted to have a treatment-limiting haemolytic event, this sample size will produce an estimate with a 95% confidence interval of +/-0.18%.

### Analysis

Quantitative analyses will be done according to an *a priori* statistical analysis plan (Supplementary Data 5). Briefly, numbers (with denominators) and proportions receiving the components of the intervention package and experiencing AESIs, or SAEs will be presented in graphical and tabular format with associated 95% confidence intervals. The change in incidence of vivax malaria between the pre-implementation Stage and Stage 2 will be shown graphically and the statistical significance of observed changes will be assessed by fitting a segmented regression model accounting for autocorrelation with adjustment for time-varying cofactors. For before versus after comparisons of count data, such as number of vivax malaria cases per month, incidence rate ratios will be calculated and negative binomial regression models fitted. Individual-level risk of representation with vivax malaria will be calculated using survival techniques including Kaplan-Meier curves and Cox proportional hazards regression.

The costs to patients and their families of *P. vivax* infection and the total healthcare provider costs (including costs of community engagement meetings, the patient education package, clinical reviews, pharmacovigilance, staff training, severe adverse events (where relevant), and malaria surveillance), will be used in combination with before-and-after case numbers to determine the overall cost-effectiveness of the package of revised case management (cost per infection or disability-adjust life-year averted). Pending discussions with decision makers, this may include a budget impact analysis and/or a distributional cost-effectiveness analysis.

Qualitative data will be set-up in a preliminary coding tree in NVivo based on operational conceptualisation of relevant outcomes of the study, i.e., feasibility, acceptability and risk appraisal. All written data will subsequently be entered and coded into the same NVivo project. Deductive coding based on the preliminary coding tree and inductive coding to yield new findings will be done. For Photovoice, analysis involves the collective interpretation of images by the community members as a way of co-creating knowledge and co-constructing meaning. To facilitate the group discussions of the photographs, the SHOWED guide is used with the following questions: What do we see here? What is really happening here? How does this relate to our lives? Why does this concern, situation or strength exist? How can we become empowered through our new understanding? What can we do?

## ETHICS AND DISSEMINATION

### Ethics approval

This study protocol (Master Protocol Version 4.0, 28 April 2023) was approved by the World Health Organization Ethics Committee (Indonesia: ERC 0003810, PNG ERC 0003892); Menzies School of Health Research (HREC: 2023-4524), Alfred Health (Burnet Institute, Project No: 18/23), University of Gadjah Mada (KE/FK/0079/EC/2023), University of Indonesia (KET 347/UN2.F1/ETIK/PPM.00.02/2023), University of Sumatra Utara (80/KEPK/USU/2023), Institute of Tropical Medicine (1655/23), PNG Institute of Medical Research (PNGIMR) (IRB: 22.02), and the PNG National Department of Health Medical Research Advisory Committee (MRAC: 22.66). The study was registered on clinicaltrials.gov for Indonesia: NCT05879224 and PNG: NCT05874271. Changes to protocols will be communicated to all involved ethics boards and relevant institutions and updated in the clinical trials registration. Stage 1 of the study began in October 2023 and was completed in September 2024.

### Participant Safety

Although pre-treatment G6PD testing will reduce the risk of severe drug-induced haemolysis, the risk cannot be eliminated completely. Severe haemolysis may still occur from *P. vivax* parasitaemia alone, misdiagnosis of G6PD status (due to an erroneous SD Biosensor result or human error by misinterpretation or incorrect prescription of PQ) or another concomitant, undiagnosed haemoglobin or red cell pathology such as the thalassaemia’s. Staff will receive refresher training sessions on the correct use of the SD Biosensor device and regular quality control will be undertaken to reduce measurement error. The day 3 (and in the case of Stage 1, the day 7) clinical review has been timed to maximise the likelihood of early detection of impending haemolysis or other severe adverse effects of primaquine. Conservative and standardised clinical escalation pathways are designed to adjust management strategies to mitigate the risk of adverse outcomes. (Figure 4).

**Figure 4.**
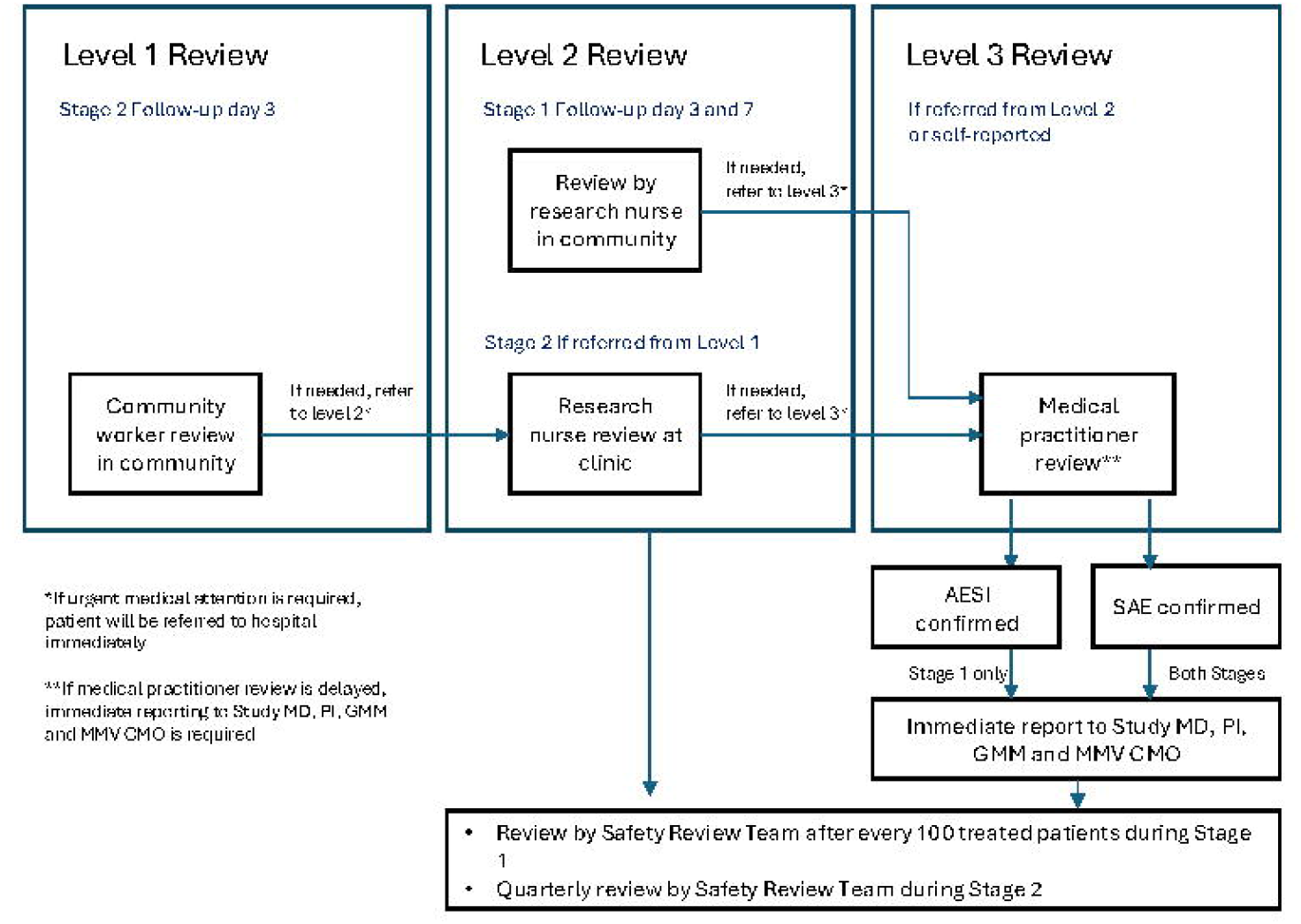
Patient Review (Level 1, 2 and 3) and Adverse Event Collection Process. Footnote: Study MD: Study medical doctor; PI: prinicipal investigator; GMM: global medical monitor; MMV CMO: Medicines for Malaria Venture Chief Medical Officer.

### Adverse Event Detection and Categorisation

The severity of adverse events will be graded according to the NCI Common Terminology Criteria for Adverse Events (CTCAE vs 5.0) (33). In addition, the Hillmen urine colour chart will be used during patient clinical reviews (34). Adverse Events of Special Interest (AESI) include haemolysis, gastrointestinal events and methaemoglobinaemia. Criteria for a haemolytic AESI include one or more of the following with onset after commencing PQ:

i. Grade 3 or 4: fatigue, breathlessness or dizziness
ii. Severe pallor or jaundice
iii. Dark urine (Hillmen score >7)
iv. Fall in haemoglobin from baseline >3g/dL
v. Fall in haemoglobin to <7g/dL

Gastrointestinal AESI criteria include Grade 3 or 4 abdominal pain, nausea, anorexia or vomiting, and methaemoglobinaemia AESI criteria include: methaemoglobin >10%, with Grade 3 or 4 breathlessness or dizziness (Supplementary Data 4).

Serious Adverse Events (SAE) will be defined as an untoward medical occurrence irrespective of cause that occurs after commencement of PQ that: results in death, is life threatening, requires inpatient hospitalisation or prolongs existing hospitalisation, results in persistent or significant disability/incapacity, is a congenital anomaly/birth defect or requires medical intervention to prevent permanent impairment or damage.

### Follow-up and Review of Adverse Events

Clinical referral pathways will be established for patients requiring advanced management of haemolysis or other adverse reactions. Community-based health care workers will actively engage communities during Stage 2 follow-up visits to identify individuals who died or required hospital admission, blood transfusion or dialysis after day 3. Prior to commencing the study, pharmacovigilance awareness will be raised in the community and at the emergency departments at local referral hospitals.

Immediately upon detection of an AESI or SAE, a cascade of adverse event reporting will be initiated including submission of the Level 3 safety review and SAE form (if applicable) to the Study Doctor, Global Medical Monitor (GMM), Principal Investigators, Study Sponsor and Chief Medical Officer from Medicines for Malaria Venture. Where possible follow-up details of each event will be provided to the GMM. Community-based health workers, research nurses and site medical practitioners will take all appropriate steps to protect the safety of participants and will ensure follow-up of the evolution of each adverse event until resolution or permanent stabilisation. The GMM will gather all safety data and report to the Safety Review Team (SRT).

### Data management

In Indonesia, hard copy clinic register, and case report forms will be stored in locked filing cabinets at the clinics. Data from these forms will be transcribed into an electronic format using REDCap (v14.6.11) data capture software. In PNG, all data will be collected directly into REDCap (v14.6.9) software (35, 36). Automatic checks of data validity will be built into the data entry forms. Verbal data from qualitative surveys will be recorded in MP3 or WAV format and subsequently transcribed into written form using standard word processing software. Patient identifiers will be stripped from datasets once data cleaning and preparation is complete. Deidentified research data will be stored for the long-term in the original electronic format, in a unified large database that contains all research data other than participant identifiable data.

### Data monitoring

#### Study coordination

The study will be coordinated by two sponsors 1) Menzies School of Health Research, Darwin, Australia (with study sites in Papua; Sumatera Utara; and Lampung, Indonesia, managed by YPKMP/ Universitas Gadjah Mada; Universitas Sumatra Utara; and University of Indonesia, respectively), and 2) Burnet Institute, Melbourne, Australia (with study sites in Baro, Mugil, Napapar and Wirui, managed by the Papua New Guinea Institute of Medical Research). The sponsors will provide study oversight via the principal investigators. Additionally, Medicines for Malaria Venture (MMV), Geneva, will provide overall study oversight to the study sponsors. An insurance policy from each Sponsor, will cover any potential medical costs if a patient becomes sick or injured because of the study. The investigators and sponsors will have access to the final dataset.

#### Safety Review Team (SRT) and Safety Monitoring Committee (SMC)

The SRT and SMC will be established to monitor safety and conduct of the study. The SRT will be managed by the GMM, and include members from the global study team, with an independent chair. The SMC will only include independent experts, and take place during Stage 1 only. The SRT will meet after every 100 patients to monitor and review individual participant safety data, or at an ad-hoc basis in the case of any safety events during Stage 1 (Figure 4). At each meeting, the SRT will vote on whether the study should be continued, modified or stopped. Interim analyses will be performed after 400 patients and after completion of Stage 1 (800 patients). The SRT will review the data package at these time points and forward a proposal to the SMC regarding the appropriateness of study continuation. The SRT will approve the progress to Stage 2 based on the SMC recommendations and endorsement from National Departments of Health in each country. The SRT will meet quarterly during Stage 2.

#### Monitoring and Auditing

A study monitoring plan with detailed monitoring templates has been prepared separately for Indonesia and PNG. Each study site will have at least three monitoring visits throughout the study period. The frequency may be increased based on the monitor’s recommendations at each site. Auditing will take place as per either the sponsor, MMV or the national health authority’s request.

### Dissemination

Dissemination plans include presentations at scientific conferences, peer-reviewed publications, and reporting at the National Ministries of Health in both countries as well as provincial and district health authorities in study locations.

## DISCUSSION

Progress towards global elimination of *P. vivax* malaria has been slow, largely because of healthcare providers’ inability to prevent relapses safely and effectively. Recommended primaquine regimens are prolonged and current doses are insufficient in many tropical areas. The risk of precipitating drug-induced haemolysis in patients with G6PD deficiency limits the use of this critically important drug in the many vivax-endemic areas where G6PD testing is unavailable. Short, high-dose primaquine regimens have potential to improve patient adherence and thus treatment effectiveness and rapid and reliable pre-treatment G6PD testing can facilitate individualised treatment for patients and reduce the risk of adverse events. Shortened courses of primaquine at a total dose of 7mg/kg are known to be efficacious for preventing relapse under trial conditions (23, 37) but the safety and effectiveness in real-world use is unknown.

The SCOPE implementation study is designed to provide robust data on the feasibility, safety, costs, and cost-effectiveness of having the typical primaquine treatment duration for patients with vivax malaria reduced to 7-days, by introducing pre-treatment point of care G6PD activity assessment and then providing double the usual primaquine total dose (7mg/kg), basic community pharmacovigilance and enhanced patient education. Following the Stage 1 phase, study conditions will emulate real-world practice as closely as possible.

This study has several important strengths. It will enrol a large number of patients with vivax malaria and provide precise estimates of the frequency of key safety outcomes. Patients will be enrolled at 10 sites, encompassing widely disparate *P. vivax* endemicities ensuring generalisability of the results to many endemic areas. The intensive first phase with rigorous community-based pharmacovigilance ensures confidence in the safety of the interventions prior to large-scale roll-out in Stage 2 while longstanding malariometric surveillance data collection at the study clinics will improve the robustness of before-versus-after comparisons of vivax malaria incidence.

The study has several limitations. Non-study malaria control activities will continue throughout the study period and will confound analyses of temporal trends. Collection of informed consent for study participation will impact clinic workflow and create a deviation from a true, real-world patient experience. Patient migration, absence of a robust national identification system, and diverse treatment seeking behaviours may all result in incomplete detection of vivax malaria recurrence resulting in an attrition bias affecting analyses of individual risk of recurrence.

The SCOPE study is endorsed by the Indonesian and PNG Ministries of Health and aligns with national malaria control priorities. Evidence provided by this study is expected to guide national vivax malaria management strategies in both countries and aim to contribute to reducing the burden of vivax malaria.

## LIST OF ABBREVIATIONS

ACT: Artemisinin-based Combination Therapy
ADL: Activity of Daily Living
AE: Adverse Event
AESI: Adverse Event of Special Interest
CbHW: Community-based Health Worker
CQI: Continuous Quality Improvement
FGD: Focus Group Discussion
G6PD: Glucose-6-Phosphate Dehydrogenase
G6PDd: G6PD deficiency
GST: Global Study Team
HCP: Healthcare Provider
MoH: Ministry of Health
NDA: National Drug Authority
NDoH: National Department of Health
NMCP: National Malaria Control Program
PCR: Polymerase Chain Reaction
PNG: Papua New Guinea PQ Primaquine
PQ7: Primaquine 1.0 mg/kg/day for 7 days
PQ14: Primaquine 0.5 mg/kg/day for 14 days
PQ8W: Primaquine 0.75mg/kg/week for 8 weeks
RDT: Rapid Diagnostic Test
SAE: Serious Adverse Event
SCOPE: Short COurse PrimaquinE for the radical cure of P. vivax
SMC: Safety Monitoring Committee
SRT: Safety Review Team
YPKMP: Yayasan Pengembangan Kesehatan dan Masyarakat Papua (Papua Health and Community Development Foundation)

## DECLARATIONS

### Consent for publication

All authors consent to publication

### Availability of data and materials

The datasets that will be generated during the current study will be made available from the corresponding authors on reasonable request.

### Patient and public involvement

Patients and/or the public were involved consulted the design, or conduct, or reporting, or dissemination plans of this research. Refer to the Methods section for further details.

### Provenance and peer review

Not commissioned; externally peer reviewed

## Acknowledgements

We thank Emilie Alirol for assistance developing the design of the study and Piero Olliaro and Arantxa Roca-Feltrer for reviewing advice on the study design and review of the protocol.

## Author contributions

ML, JRP, AP, IS, RNP, LJR, ND, SD conceived the study and led the protocol development; ER, VSS, EN, LF, MM, AR, RF, KM, PD, BL, PA, AD, GL, MT, AS, RN, SP, HH and EJ contributed to protocol development. ML, JRP, AP, IS, RNP, LJR, ND, SD, JA, LF, FA, SA, AS, RN, MM, MOK, VSS, ER, GL, KM, PD, TD, HH, HD, TW contributed to study materials development. JAS, ND and RNP developed the statistical analysis plan and GL the data management plan. All authors read and approved the final protocol manuscript.

## References

1. Price RN, Commons RJ, Battle KE, Thriemer K, Mendis K. Plasmodium vivax in the Era of the Shrinking P. falciparum Map. Trends Parasitol. 2020;36(6):560–70.

2. Commons RJ, Simpson JA, Watson J, White NJ, Price RN. Estimating the Proportion of Plasmodium vivax Recurrences Caused by Relapse: A Systematic Review and Meta-Analysis. Am J Trop Med Hyg. 2020;103(3):1094–9.

3. World Health Organisation. World Malaria Report 2023. 2023.

4. Rueangweerayut R, Bancone G, Harrell EJ, Beelen AP, Kongpatanakul S, Mohrle JJ, et al. Hemolytic Potential of Tafenoquine in Female Volunteers Heterozygous for Glucose-6-Phosphate Dehydrogenase (G6PD) Deficiency (G6PD Mahidol Variant) versus G6PD-Normal Volunteers. Am J Trop Med Hyg. 2017;97(3):702–11.

5. Thriemer K, Ley B, von Seidlein L. Towards the elimination of Plasmodium vivax malaria: Implementing the radical cure. PLoS Med. 2021;18(4):e1003494.

6. TGA. Australian Public Assessment Report: Tafenoquine succinate 2019 [Available from: https://www.tga.gov.au/resources/auspar/auspar-tafenoquine-succinate-0.

7. FDA. Krintafel (Tafenoquine) 2018 [Available from: https://www.accessdata.fda.gov/drugsatfda_docs/label/2018/210795s000lbl.pdf.

8. FDA. Arakoda (Tafenoquine) Tablets 2018 [Available from: https://www.accessdata.fda.gov/drugsatfda_docs/label/2018/210607lbl.pdf.

9. Recht J, Ashley EA, White NJ. Use of primaquine and glucose-6-phosphate dehydrogenase deficiency testing: Divergent policies and practices in malaria endemic countries. PLoS Negl Trop Dis. 2018;12(4):e0006230.

10. Mehdipour P, Rajasekhar M, Dini S, Zaloumis S, Abreha T, Adam I, et al. Effect of adherence to primaquine on the risk of Plasmodium vivax recurrence: a WorldWide Antimalarial Resistance Network systematic review and individual patient data meta-analysis. Malar J. 2023;22(1):306.

11. Thriemer K, Ley B, Bobogare A, Dysoley L, Alam MS, Pasaribu AP, et al. Challenges for achieving safe and effective radical cure of Plasmodium vivax: a round table discussion of the APMEN Vivax Working Group. Malar J. 2017;16(1):141.

12. World Health Organisation. WHO Guidelines for Malaria. Geneva: WHO; 2023.

13. Devine A, Battle KE, Meagher N, Howes RE, Dini S, Gething PW, et al. Global economic costs due to vivax malaria and the potential impact of its radical cure: A modelling study. PLoS Med. 2021;18(6):e1003614.

14. Paediatric Society of Papua New Guinea. Standard Treatment for Common Illnesses of Children in Papua New Guinea. 2016.

15. Ministry of Health Republic of Indonesia. Pocket book on Malaria Case Management. 2023.

16. Rahmalia A, Poespoprodjo JR, Landuwulang CUR, Ronse M, Kenangalem E, Burdam FH, et al. Adherence to 14-day radical cure for Plasmodium vivax malaria in Papua, Indonesia: a mixed-methods study. Malar J. 2023;22(1):162.

17. Douglas NM, Poespoprodjo JR, Patriani D, Malloy MJ, Kenangalem E, Sugiarto P, et al. Unsupervised primaquine for the treatment of Plasmodium vivax malaria relapses in southern Papua: A hospital-based cohort study. PLoS Med. 2017;14(8):e1002379.

18. Poespoprodjo JR, Burdam FH, Candrawati F, Ley B, Meagher N, Kenangalem E, et al. Supervised versus unsupervised primaquine radical cure for the treatment of falciparum and vivax malaria in Papua, Indonesia: a cluster-randomised, controlled, open-label superiority trial. Lancet Infect Dis. 2021.

19. Abreha T, Hwang J, Thriemer K, Tadesse Y, Girma S, Melaku Z, et al. Comparison of artemether-lumefantrine and chloroquine with and without primaquine for the treatment of Plasmodium vivax infection in Ethiopia: A randomized controlled trial. PLoS Med. 2017;14(5):e1002299.

20. Baird JK, Hoffman SL. Primaquine Therapy for Malaria. Clinical Infectious Diseases. 2004;39(9):1336–45.

21. Commons RJ, Rajasekhar M, Edler P, Abreha T, Awab GR, Baird JK, et al. Effect of primaquine dose on the risk of recurrence in patients with uncomplicated Plasmodium vivax: a systematic review and individual patient data meta-analysis. Lancet Infect Dis. 2023.

22. Chu CS, Phyo AP, Turner C, Win HH, Poe NP, Yotyingaphiram W, et al. Chloroquine Versus Dihydroartemisinin-Piperaquine With Standard High-dose Primaquine Given Either for 7 Days or 14 Days in Plasmodium vivax Malaria. Clin Infect Dis. 2019;68(8):1311–9.

23. Taylor WRJ, Thriemer K, von Seidlein L, Yuentrakul P, Assawariyathipat T, Assefa A, et al. Short-course primaquine for the radical cure of Plasmodium vivax malaria: a multicentre, randomised, placebo-controlled non-inferiority trial. Lancet. 2019;394(10202):929–38.

24. Luzzatto L. Glucose 6-phosphate dehydrogenase deficiency: from genotype to phenotype. Haematologica. 2006;91(10):1303–6.

25. Rajasekhar M, Simpson JA, Ley B, Edler P, Chu CS, Abreha T, et al. Primaquine dose and the risk of haemolysis in patients with uncomplicated Plasmodium vivax malaria: a systematic review and individual patient data meta-analysis. Lancet Infect Dis. 2023.

26. Chu CS, Bancone G, Moore KA, Win HH, Thitipanawan N, Po C, et al. Haemolysis in G6PD Heterozygous Females Treated with Primaquine for Plasmodium vivax Malaria: A Nested Cohort in a Trial of Radical Curative Regimens. PLoS Med. 2017;14(2):e1002224.

27. Yilma D, Groves ES, Brito-Sousa JD, Monteiro WM, Chu C, Thriemer K, et al. Severe Hemolysis during Primaquine Radical Cure of Plasmodium vivax Malaria: Two Systematic Reviews and Individual Patient Data Descriptive Analyses. Am J Trop Med Hyg. 2023;109(4):761–9.

28. Ley B, Winasti Satyagraha A, Rahmat H, von Fricken ME, Douglas NM, Pfeffer DA, et al. Performance of the Access Bio/CareStart rapid diagnostic test for the detection of glucose-6-phosphate dehydrogenase deficiency: A systematic review and meta-analysis. PLoS Med. 2019;16(12):e1002992.

29. Pal S, Bansil P, Bancone G, Hrutkay S, Kahn M, Gornsawun G, et al. Evaluation of a Novel Quantitative Test for Glucose-6-Phosphate Dehydrogenase Deficiency: Bringing Quantitative Testing for Glucose-6-Phosphate Dehydrogenase Deficiency Closer to the Patient. Am J Trop Med Hyg. 2019;100(1):213–21.

30. Alam MS, Kibria MG, Jahan N, Thriemer K, Hossain MS, Douglas NM, et al. Field evaluation of quantitative point of care diagnostics to measure glucose-6-phosphate dehydrogenase activity. PLoS One. 2018;13(11):e0206331.

31. Eldridge SM, Chan CL, Campbell MJ, Bond CM, Hopewell S, Thabane L, et al. CONSORT 2010 statement: extension to randomised pilot and feasibility trials. Pilot and Feasibility Studies. 2016;2(1).

32. SPIRIT 2013 Statement: Defining Standard Protocol Items for Clinical Trials. Annals of Internal Medicine. 2013;158(3):200–7.

33. Institute NC. Common Terminology Criteria for Adverse Events (CTCAE) 2020 [Available from: https://ctep.cancer.gov/protocoldevelopment/electronic_applications/ctc.htm.

34. Hillmen P, Hall C, Marsh JCW, Elebute M, Bombara MP, Petro BE, et al. Effect of Eculizumab on Hemolysis and Transfusion Requirements in Patients with Paroxysmal Nocturnal Hemoglobinuria. New England Journal of Medicine. 2004;350(6):552–9.

35. Harris PA, Taylor R, Thielke R, Payne J, Gonzalez N, Conde JG. Research electronic data capture (REDCap)--a metadata-driven methodology and workflow process for providing translational research informatics support. J Biomed Inform. 2009;42(2):377–81.

36. Harris PA, Taylor R, Minor BL, Elliott V, Fernandez M, O’Neal L, et al. The REDCap consortium: Building an international community of software platform partners. J Biomed Inform. 2019;95:103208.

37. Thriemer K, Degaga TS, Christian M, Alam MS, Rajasekhar M, Ley B, et al. Primaquine radical cure in patients with Plasmodium falciparum malaria in areas co-endemic for P falciparum and Plasmodium vivax (PRIMA): a multicentre, open-label, superiority randomised controlled trial. The Lancet. 2023.

