## Supplementary data 2 for "High daily dose Short COurse PrimaquinE after G6PD testing for the radical cure of *Plasmodium vivax* malaria in Indonesia and Papua New Guinea: The SCOPE implementation study protocol"

Supplementary File 2: SCOPE Study Clinics Primaquine Dosing

**Table 1. Primaquine regimen based on G6PD activity for Indonesia and PNG**

| **Classification** | **G6PD activity level** | | **Treatment arm** | **Target dose of Primaquine** |
| --- | --- | --- | --- | --- |
|  | % | U/g Hb |  |  |
| **Normal** | > 70% | ≥ 6.1 U/g Hb | PQ7 | Daily dose : 1 mg/kg/day  Duration : 7 days  Total dose : 7 mg/kg |
| **Intermediate** | 30-70% | 4.1 – 6.0 U/g Hb | PQ14 | Daily dose : 0.5mg/kg/day  Duration : 14 days  Total dose : 7 mg/kg |
| **Deficient** | < 30% | ≤ 4.0 U/g Hb | PQ8w | Weekly dose : 0.75 mg/kg/week  Duration : 8 weeks  Total dose : 6 mg/kg |

**Table 2 SCOPE Primaquine Dosing in Indonesia using 15mg PQ tablets**

| **G6PD activity** | **Treatment**  **Arm** | **Frequency and duration** | **Dose- Number of tablets per day according to body weight (kg)** | | | | | | |
| --- | --- | --- | --- | --- | --- | --- | --- | --- | --- |
|  |  |  | **< 6** | **6-11** | **12-20** | **21-25** | **26-35** | **36-50** | **≥ 51** |
| **Normal:**  ≥ 6.1 U/g Hb | **PQ7** | Once daily, for 7 days | - | ½ | 1 | 1½ | 2 | 3 | 4 |
| **Intermediate:**  4.1 – 6.0 U/g Hb | **PQ14** | Once daily, for 14 days | - | ¼ | ½ | ¾ | 1 | 1½ | 2 |
| **Deficient:**  ≤ 4.0 U/g Hb | **PQ8w** | Once weekly, for 8 weeks | - | ½ | 1 | 1½ | 1½ | 2 | 3 |

**Table 3 SCOPE Primaquine Dosing in PNG using 7.5mg PQ tablets**

| **G6PD activity** | **Treatment**  **Arm** | **Frequency and duration** | **Dose- Number of tablets per day according to body weight (kg)** | | | | | | |
| --- | --- | --- | --- | --- | --- | --- | --- | --- | --- |
|  |  |  | **< 6** | **6-11** | **12-20** | **21-25** | **26-35** | **36-50** | **≥ 51** |
| **Normal:**  ≥ 6.1 U/g Hb | **PQ7** | Once daily, for 7 days | - | 1 | 2 | 3 | 4 | 6 | 8 |
| **Intermediate:**  4.1 – 6.0 U/g Hb | **PQ14** | Once daily, for 14 days | - | ½ | 1 | 1½ | 2 | 3 | 4 |
| **Deficient:**  ≤ 4.0 U/g Hb | **PQ8w** | Once weekly, for 8 weeks | - | 1 | 2 | 3 | 3 | 4 | 6 |
