## Supplementary data 3 for "High daily dose Short COurse PrimaquinE after G6PD testing for the radical cure of *Plasmodium vivax* malaria in Indonesia and Papua New Guinea: The SCOPE implementation study protocol"

**Supplementary File 3:** Master Review forms: 1) Pre-implementation form, 2) Baseline form, 3) Level 1, 4) Level 2, 5) Level 3 Review Form, and 6) SAE Form.

| Presentation Details | | | | | | |
| --- | --- | --- | --- | --- | --- | --- |
| Visit Date: __ / __ / 202__ | | | **SCOPE Study ID:** | | | |
| Participant Details | | | | | | |
| Sex: □ M □ F | | **Age:** ____ yrs ____ mths | | | **Weight:** _____ . ___kg | **Pregnant: □** Y **□** N **□** Unknown |
| Clinic Malaria Diagnosis | | | | | | |
| Fever or History of Fever in Last 48 hours: □ Yes □ No □ Unknown | | | | | | |
| Diagnostic: | □ Microscopy □ RDT | | | **Ringform:** + -- □ Unknown | | **Gametocyte:** + -- □ Unknown |
| Diagnosis: | *□ P. vivax □ P. falciparum □ P. malariae □ P. ovale □ Pv/Pan (RDT) □* Negative | | | | | |
| G6PD and Hb | | | | | | |
| G6PD Activity: ______ . ___ U/g Hb | | | | | **Hb:** ______ . ___ g/dL | |
| Sample Type: □ Capillary □ Venous | | | | | | |

**Participant Details**

**SCOPE Study ID:**

| **Microscopy Malaria Diagnosis** | | | | | | | | | | | | | |
| --- | --- | --- | --- | --- | --- | --- | --- | --- | --- | --- | --- | --- | --- |
| **Collection Date:** \|__\|__\| / \|__\|__\| / \| 2 \| 0 \| 2 \|__\|  D D M M Y Y Y Y | | | | | | | **Collection Time (24hr):** \|__\|__\|:\|__\|__\|  H H M M | | **Microscopist:** | | | **Read Number:** □ 1st □ 2nd □ 3rd | |
| **#** | **Smear type**  Select one only | **Slide quality**  Select one only | | | **Parasite type**  Select one only | | **Parasite count** | **Parasite count units** | | | | | **Malaria species**  Select one only |
| 1 | □ Thick Smear  □ Thin Smear | □ Good  □ Poor  □ Missing | | | □ Asexual  □ Sexual  □ Negative | | \|__\|__\|__\|__\|__\| | / \|__\|__\|__\|__\| WBC | | / \|__\|__\|__\|__\| HPF | / \|__\|__\|__\|__\| RBC | | □ Pv □ Pf □ Pm □ Po  □ None Seen |
| 2 | □ Thick Smear  □ Thin Smear | □ Good  □ Poor  □ Missing | | | □ Asexual  □ Sexual  □ Negative | | \|__\|__\|__\|__\|__\| | / \|__\|__\|__\|__\| WBC | | / \|__\|__\|__\|__\| HPF | / \|__\|__\|__\|__\| RBC | | □ Pv □ Pf □ Pm □ Po  □ None Seen |
| 3 | □ Thick Smear  □ Thin Smear | □ Good  □ Poor  □ Missing | | | □ Asexual  □ Sexual  □ Negative | | \|__\|__\|__\|__\|__\| | / \|__\|__\|__\|__\| WBC | | / \|__\|__\|__\|__\| HPF | / \|__\|__\|__\|__\| RBC | | □ Pv □ Pf □ Pm □ Po  □ None Seen |
| 4 | □ Thick Smear  □ Thin Smear | □ Good  □ Poor  □ Missing | | | □ Asexual  □ Sexual  □ Negative | | \|__\|__\|__\|__\|__\| | / \|__\|__\|__\|__\| WBC | | / \|__\|__\|__\|__\| HPF | / \|__\|__\|__\|__\| RBC | | □ Pv □ Pf □ Pm □ Po  □ None Seen |
| 5 | □ Thick Smear  □ Thin Smear | □ Good  □ Poor  □ Missing | | | □ Asexual  □ Sexual  □ Negative | | \|__\|__\|__\|__\|__\| | / \|__\|__\|__\|__\| WBC | | / \|__\|__\|__\|__\| HPF | / \|__\|__\|__\|__\| RBC | | □ Pv □ Pf □ Pm □ Po  □ None Seen |
| *Note: Complete separate rows if both Asexual and Sexual parasites are found. Complete separate rows for each species if mixed infection.*  *Note: Complete a separate page for each read (1^st^, 2^nd^, 3^rd^)* | | | | | | | | | | | | | |

| SCOPE Participant Details | | | | | | |
| --- | --- | --- | --- | --- | --- | --- |
| Visit Date: ___ / ___ / 202__ | | **Malaria Card #:** _____________ | | | **SCOPE Study ID:** _____________ | |
| Patient Initials: | | **Sex**: □ M □ F | | | **Age**^1^**:** ____ yrs ____ months | |
| Weight: ______ . ___kg | | **Pregnant: □** Y **□** N **□** Unknown | | | **Lactating <6m: □** Y **□** N **□** Unknown | |
| Ethnic Group: | | | | | | |
| Malaria Diagnosis | | | | | | |
| Diagnostic: □ Microscopy □ RDT | | | **Ringform**: □ + □ -- □ Unknown | | | **Gametocyte:** □ + □ -- □ Unknown |
| Diagnosis: □ *P. vivax □ P. falciparum □ P. malariae □ P. ovale* □ Pv/Pan (RDT) □ Negative | | | | | | |
| G6PD, Hb and Treatment Duration | | | | | | |
| Hemocue Hb ^2^: ______ . ___ g/dL | | | | | | |
| Biosensor | | | **G6PD Category** | | | **PQ Treatment Duration** |
| G6PD: ______ . ___ U/g Hb  Hb: ______ . ___ g/dL | | | **□** Normal (≥6.1 U/g Hb) | | | □ 7 days (PQ7) |
|  |  |  | **□** Intermediate (4.1-6.0 U/g Hb) | | | □ 14 days (PQ14) |
|  |  |  | **□** Deficient (≤4.0 U/g Hb) | | | □ 8 weeks (PQ8W) |
| Staff ID (G6PD Tester): | | | | | | |
| Treatment | | | | | | |
| PQ: | ______ total tablets | **1st dose PQ observed:** | | □ Yes □ No | | |
|  |  | **If Yes, taken with food?** | | □ Yes □ No | | |
|  |  | **If No, reason:**  **□** Has not eaten  **□** Prefers to take at home  **□** Refused | | **.**  **□** Cannot swallow tablets  **□** No water or other liquids available  **□** Other (specify) _____________________ | | |
| DHP: | ______ total tablets | **1st dose DHP observed:** | | □ Yes □ No | | |
|  |  | **If No, reason:**  **□** Has not eaten  **□** Prefers to take at home  **□** Refused | | **□** Cannot swallow tablets  **□** No water or other liquids available  **□** Other (specify) _____________________ | | |
| Other: | | | | | | |
| Staff ID (Prescriber): | | | | | | |

| Level 1 Review Form – COMMUNITY HEALTH WORKER EVALUATION | | | | | |
| --- | --- | --- | --- | --- | --- |
| Date of review: ___ / ___ / 202__ Day of Review: □ Day 3 □ Day _______ | | | | | |
| Format of review: □ Phone call □ Personal visit | | | | | |
| PATIENT DETAILS | | | | | |
| Patient Initials: ______________ Malaria card number: ________________________  Sex: □ M □ F Age: _____ yrs _____ mths | | | | | |
| YESTERDAY  Yesterday did you take your malaria tablets with food? □ Yes □ No □ I didn’t take any | | | | | |
| Do your tablets make you feel sick? □ Yes □ No | | | | | |
| SYMPTOM CHECKLIST | | | | | |
| Mild/ Moderate | Mild/Moderate abdominal pain | | | Yes / No | ***If any…***  ***remind patient to take tablets with food or split PQ tablets morning and evening*** |
|  | Vomiting (only once) | | | Yes / No |  |
|  | Feeling sick | | | Yes / No |  |
| Severe | Dark (red or black) urine | | | Yes / No | ***If any…***  ***withhold PQ and refer patient to clinic for review*** |
|  | Severe abdominal / back pain | | | Yes / No |  |
|  | Unable to eat food | | | Yes / No |  |
|  | Vomiting (>2x in a day) | | | Yes / No |  |
|  | Significant breathlessness on exertion | | | Yes / No |  |
|  | Dizziness / unable to stand up | | | Yes / No |  |
| Other Significant Symptoms (specify):__________________________________________________________ | | | | | |
| ACTION: | | | | | |
| Tick all that apply | | □ | No change to primaquine administration | | |
|  |  | □ | Education on completing treatment (if no grey flags) | | |
|  |  | □ | Primaquine withheld | | |
|  |  | □ | Referred for Level 2 Review | | |

| ­­Level 2 Review Form - NURSE EVALUATION | | | |
| --- | --- | --- | --- |
| Date of review: ___ / ___ / 202__ Day of Review: □ Day3 □ Day7 □ Day _____ | | | |
| Location of Assessment: □ Home □ Clinic □ Other ________________ | | | |
| PATIENT DETAILS | | | |
| SCOPE ID: ____________ Patient Initials: ___________ Malaria card number: ______________ Sex: □ M □ F Age: _____ yrs ____ mths Initial Treatment: □ PQ7 □ PQ14 □ PQ8W | | | |
| Yesterday did you take your malaria tablets? | | □ Yes □ No □ Already completed tablets | |
| If Yes, did you take them with food? | | □ Yes □ No | |
| REPORTED REACTION(S) | | | |
| Temperature: ____.___ C | **Pulse:** _____ per min | | **Resp Rate:** ____per min |
| Symptom | **Severity (see criteria table)** | | **If present, date of onset** |
| Abdominal pain | None 1 2 3 4 | | ___/___/202__ |
| Nausea | None 1 2 3 4 | | ___/___/202__ |
| Unable to eat | None 1 2 3 4 | | ___/___/202__ |
| Vomiting | None 1 2 3 4 | | ___/___/202__ |
| Back pain | None 1 2 3 4 | | ___/___/202__ |
| Breathlessness | None 1 2 3 4 | | ___/___/202__ |
| Dizziness | None 1 2 3 4 | | ___/___/202__ |
| Fatigue | None 1 2 3 4 | | ___/___/202__ |
| Vomiting >2x per day | Yes / No | | ___/___/202__ |
| Jaundice | Yes / No | | ___/___/202__ |
| Blue lips | Yes / No | | ___/___/202__ |
| Dark urine | Yes / No | | ___/___/202__ |
| Other : | ________________________ | | ___/___/202__ |

| INVESTIGATIONS | | | | | | | | |
| --- | --- | --- | --- | --- | --- | --- | --- | --- |
| Hemocue | **Date** | | |  | | **Hemocue Hb** | | Change in Hb (B – A)^[1]^ |
| Baseline (A) | ___/___/202__ | | |  | | ____.____ g/dL | |  |
| Result today (B) | ___/___/202__ | | |  | | ____.____ g/dL | | ____.____ g/dL |
| Biosensor | **Date** | | | **G6PD Activity** | | **Biosensor Hb** | | Change in Hb (D– C) |
| Baseline (C) | ___/___/202__ | | | ___.___U/g Hb | | ____.____ g/dL | |  |
| Result today (D) | ___/___/202__ | | | ___.___U/g Hb | | ____.____ g/dL | | ____.____ g/dL |
| MetHb (if measured) | ___/___/202__ | | | ______.____ % | |  | |  |
| MANAGEMENT | | | | | | | | |
| Need for Referral? | | □ Any Grey Flags □ Fall in Hb >3g/dL □ Hb <7 g/dL □ Referral Not Required | | | | | | |
| Primaquine | | □ Continued □ Withheld □ Finished □ Modified | | | | | | |
|  | | Dose: ___.__ mg □ 1x day □ 2x day Duration: ___ days | | | | | | |
| Referred for Level 3 Review | | | **□** Yes □ No | | If Yes | | Date: __ /__ / 202__ Time: ___:___ | |
| Level 3 Confirmation | | | **□** Yes □ No | | If Yes | | Date: __ /__ / 202__ Time: ___:___ | |
| Notification: If patient won’t be reviewed by Level 3 within 3 hrs | | | | | | | | |
| WhatsApp Group | | | □ Yes □ No | | If Yes | | Date: __ /__ / 202__ Time: ___:___ | |

| Education Given: | □ Yes □ No |
| --- | --- |
| Decision Tree | |

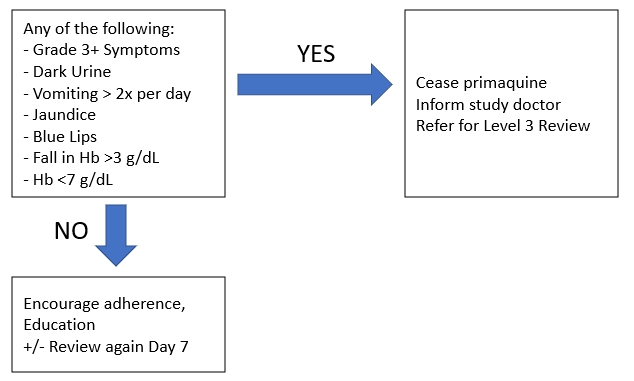

| **Term** | **Grade 1** | **Grade 2** | **Grade 3** | **Grade 4** |
| --- | --- | --- | --- | --- |
| **Abdominal pain** | Mild pain | Moderate pain; limiting instrumental Activity of Daily Living (ADL) | Severe pain; limiting self-care ADL |  |
| **Nausea** | Loss of appetite without alteration in eating habits | Oral intake decreased without significant weight loss, dehydration or malnutrition | Inadequate oral caloric or fluid intake; tube feeding, or hospitalisation indicated |  |
| **Anorexia** | Loss of appetite without alteration in eating habits | Oral intake altered without significant weight loss or malnutrition; oral nutritional supplements indicated | Associated with significant weight loss or malnutrition | Life-threatening consequences; urgent intervention indicated |
| **Vomiting** | Intervention not indicated | Outpatient IV hydration; medical intervention indicated | Tube feeding, or hospitalisation indicated | Life-threatening consequences |
| **Back Pain** | Mild pain | Moderate pain; limiting instrumental ADL | Severe pain; limiting self-care ADL |  |
| **Breathlessness** | Shortness of breath with moderate exertion | Shortness of breath with minimal exertion; limiting instrumental ADL | Shortness of breath at rest; limiting self-care ADL | Life-threatening consequences; urgent intervention indicated |
| **Dizziness** | Mild unsteadiness or sensation of movement | Moderate unsteadiness or sensation of movement; limiting instrumental ADL | Severe unsteadiness or sensation of movement; limiting self-care ADL |  |
| **Fatigue** | Fatigue relieved by rest | Fatigue not relieved by rest; limiting instrumental ADLs | Fatigue not relieved by rest; limiting self care ADLs |  |

| **Level 3 Review Form - Medical Evaluation** | | |
| --- | --- | --- |
| **To be completed for all patients referred after Level 2 review with suspected SAE / AESI**  **Location: □ Clinic □ Hospital □ Other: _____________**  **Report type: □ Initial □ Follow-up FU #: ____**  **Date of Level 3 Review: __/ __/ 202___ Time: ____:____** | | |
| **PATIENT DETAILS** | | |
| **SCOPE ID: ___________ Patient Initials: _____ Malaria card number: ______________**  **Sex: □ M □ F Age: ___ yrs ___mths Initial Treatment: □ PQ7 □ PQ14 □ PQ8W** | | |
| **CLINICAL PRESENTATION** | | |
| **Date referred** | **___ / ___ / 202__** | |
| **Temperature:____.___ C** | **Pulse: _____ per min** | **Resp rate: ____ per min** |
| **Symptom** | **Severity (criteria table)** | **If present, date of onset** |
| **Abdominal pain** | **None 1 2 3 4** | **___/___/202__** |
| **Nausea** | **None 1 2 3 4** | **___/___/202__** |
| **Unable to eat** | **None 1 2 3 4** | **___/___/202__** |
| **Vomiting** | **None 1 2 3 4** | **___/___/202__** |
| **Back pain** | **None 1 2 3 4** | **___/___/202__** |
| **Breathlessness** | **None 1 2 3 4** | **___/___/202__** |
| **Dizziness** | **None 1 2 3 4** | **___/___/202__** |
| **Fatigue** | **None 1 2 3 4** | **___/___/202__** |
| **Fever** | **Yes / No** | **___/___/202__** |
| **Severe pallor** | **Yes / No** | **___/___/202__** |
| **Jaundice** | **Yes / No** | **___/___/202__** |
| **Cyanosis (Blue lips)** | **Yes / No** | **___/___/202__** |
| **Dark (red or black) urine** | **Yes / No**  **Colour #:______** | **___/___/202__** |
| **Other: _______________** |  | **___/___/202__** |
| **NARRATIVE:**  **______________________________________________________________________________________________________________________________________________________________________________________________________________________________________________________________________________________________________________________________________________________________________________________________________________________________________________________** | | |

| **INVESTIGATIONS** | | | | |
| --- | --- | --- | --- | --- |
| **Haemoglobin** | **Date** | | **Result** | **Method** |
| **Baseline (A)** | **__ / __ / 202__** | | **Hb: ___ . __ g/dL** | **□ Hemocue □ CBC** |
| **Today (B)** | **__ / __ / 202__** | | **Hb: ___ . __ g/dL** | **□ Hemocue □ CBC** |
| **Nadir (C)** | **__ / __ / 202__** | | **Hb: ___ . __ g/dL** | **□ Hemocue □ CBC** |
| **Max Fall Hb: ____ . __ g/dL (C-A)** | | | **Max % Fall Hb: ____.__ %** | |
| **Biosensor Hb** | **Date** | | **Result** |  |
| **Baseline (D)** | **__ / __ / 202__** | | **Hb: ___ . __ g/dL** |  |
| **Today (E)** | **__ / __ / 202__** | | **Hb: ___ . __ g/dL** |  |
| **Nadir (F)** | **__ / __ / 202__** | | **Hb: ___ . __ g/dL** |  |
| **Max Fall Hb: ____ . __ g/dL (F-D)** | | | **Max % Fall Hb: ____.__ %** | |
| **G6PD: Baseline** | **__ / __ / 202__** | | **___. ___ U/g Hb** |  |
| **______________** | **__ / __ / 202__** | | **___. ___ U/g Hb** |  |
| **______________** | **__ / __ / 202__** | | **___. ___ U/g Hb** |  |
| **Met Hb:** | **__ / __ / 202__** | | **___. ___ %** | **For all patients at Level 3** |
| **ADVERSE EVENT CLASSIFICATION** | | | | |
| **Severity** | | **Max Graded Symptom: Date of onset:___/___/202__**  **□ Grade 1 □ Grade 2 □ Grade 3 □ Grade 4 □ Grade 5** | | |
| **For Follow Up Visits** | | **□ Recovered/resolved □ Recovering/resolving □ Worsening** | | |
| **AESI - Haemolysis**  **□ Yes □ No** | | **Date of onset: ___/___/202__**  **Any of the following:**  **□ Grade 3 or 4: Fatigue, Dizziness, Breathlessness (onset after starting PQ)**  **□ Severe Pallor or Jaundice**  **□ Dark Urine: Hillmen >7**  **□ Fall in Hb > 3 g/dL**  **□ Hb <7g/dl** | | |
| **AESI - Gastrointestinal**  **□ Yes □ No** | | **Date of onset: ___/___/202__**  **□ Grade 3 / 4: Abdominal Pain, Nausea, Anorexia or Vomiting** | | |
| **AESI - MetHb**  **□ Yes □ No** | | **□ MetHb: >10% Date of onset: ___/___/202__**  **AND Grade 3 or 4: Breathlessness or Dizziness** | | |
| **SAE**  **□ Yes □ No** | | **□ Death** | | |
|  |  | **□ Life threatening** | | |
|  |  | **□ Hospitalisation or prolongation of hospitalisation** | | |
|  |  | **□ Persistent or significant disability** | | |
|  |  | **□ Congenital abnormality/birth defect** | | |
|  |  | **□ Other________________________** | | |

| **CLINICAL MANAGEMENT** | | |
| --- | --- | --- |
| **Changes to PQ** | **□ Continued □ Withheld □ Finished □ Ceased □ Restarted □ Modified** | |
|  | **Dose: ___.__ mg □ 1x day □ 2x day Duration: ___ days** | |
| **NARRATIVE for AESI: If it meets criteria for SAE, complete SAE Form □**  **_________________________________________________________________________**  **_________________________________________________________________________**  **_________________________________________________________________________**  **_________________________________________________________________________**  **____________________________________________________________________________________________________________________________________________________________________________________________________________________________________________________________________________________________________** | | |
| **Planned Review** | | **□ No further follow-up required**  **□ Routine Follow Up: □ Day 3 or □ Day 7**  **□ Review again on Day ____ Date: __ / __ / 202__**  **□ Referred to Hospital**  **If Referred: Date: __ / __ / 202__ Time: __ : __** |
| **NOTIFICATION □ Yes □ No**  **Any confirmed AESI must be notified to the PIs and GMM as soon as possible**  **If an SAE, complete an SAE form and send report to PIs, GMM and MMV within 24 hrs** | | |
| **Time of Notification** | | **Date: __ / __ / 202____ Time: ___ : ___** |
| **Completion of SAE Form** | | **□ Yes □ Not Applicable** |
| **Process** | | **□ WhatsApp**  **□ Email scanned Level 3 Form +/- SAE Form** |
| **People Notified** | | **□ Local PI**  **□ Study MD**  **□ Menzies / Burnet PI**  **□ Global Medical Monitor**  **□ MMV Chief Medical Officer** |

| **Term** | **Grade 1** | **Grade 2** | **Grade 3** | **Grade 4** |
| --- | --- | --- | --- | --- |
| **Abdominal pain** | **Mild pain** | **Moderate pain; limiting instrumental ADL** | **Severe pain; limiting self-care ADL** |  |
| **Nausea** | **Loss of appetite without alteration in eating habits** | **Oral intake decreased without significant weight loss, dehydration or malnutrition** | **Inadequate oral caloric or fluid intake; tube feeding, or hospitalisation indicated** |  |
| **Anorexia** | **Loss of appetite without alteration in eating habits** | **Oral intake altered without significant weight loss or malnutrition; oral nutritional supplements indicated** | **Associated with significant weight loss or malnutrition** | **Life-threatening consequences; urgent intervention indicated** |
| **Vomiting** | **Intervention not indicated** | **Outpatient IV hydration; medical intervention indicated** | **Tube feeding, or hospitalisation indicated** | **Life-threatening consequences** |
| **Back Pain** | **Mild pain** | **Moderate pain; limiting instrumental ADL** | **Severe pain; limiting self-care ADL** |  |
| **Breathlessness** | **Shortness of breath with moderate exertion** | **Shortness of breath with minimal exertion; limiting instrumental ADL** | **Shortness of breath at rest; limiting self-care ADL** | **Life-threatening consequences; urgent intervention indicated** |
| **Dizziness** | **Mild unsteadiness or sensation of movement** | **Moderate unsteadiness or sensation of movement; limiting instrumental ADL** | **Severe unsteadiness or sensation of movement; limiting self-care ADL** |  |
| **Fatigue** | **Fatigue relieved by rest** | **Fatigue not relieved by rest; limiting instrumental ADLs** | **Fatigue not relieved by rest; limiting self care ADLs** |  |

**HILLMEN URINE CHART**

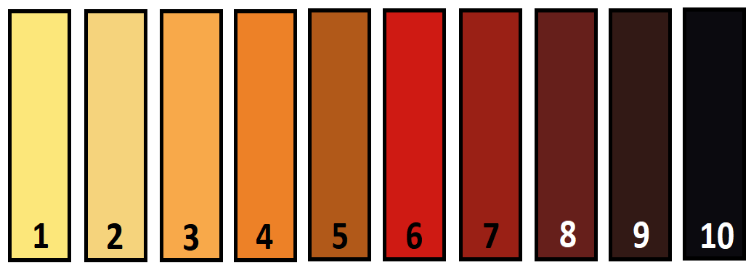

| **REPORT DETAILS** | |
| --- | --- |
| **Date of Assessment**: __ / __ / 202__ | **Time**: ___ : ___ |
| **Report type:** □ Initial □ Follow-up FU #: ____ | |
| **Location of Assessment (hospital/clinic name):** | |
| **PATIENT DETAILS** | |
| **Patient Initials**: _______________ **Medical ID number:** ________________________  **Sex**: □ M □ F **Age:** _______yrs _____ mths **Date of Birth:** __ /__ /202___  **Weight**: _______ . __ kg  **Country**: ____________________ | |
| **MALARIA DETAILS** - Only complete for the initial SAE report | |
| **Date of Malaria Diagnosis**: __ / __ / 202__ **Method:** □ Microscopy □ RDT □ Other___ | |
| **Malaria Species**: □ Pf □ Pv □ Pm □ Po □ Unknown | |
| **Baseline Parasitaemia**: ______________ per uL □++++ □+++ □ ++ □+ □ Unknown | |
| **TREATMENT DETAILS** - Only complete for the initial SAE report | |
| **Schizontocidal Treatment**: □ CQ □ AL □ DP □ Quinine □ Other:___________  **Date PQ Commenced**: ___ /___ /202____  **Daily Dose of PQ**: _______.__ mg **Calculated mg/kg Dose:**__________  **PQ Dosing schedule**: □ 1x day □ 2x day □ 1x week □ No Tablets taken  **Planned duration of PQ**: □ 7 days □ 14 days □ 8 Weeks | |
| **Number of PQ doses taken before adverse event detected**: ________ | |
| **Did the patient take their last PQ tablets with food**? □ Yes □ No □ Unsure | |

| **CONCOMITANT MEDICATION** | | | | |
| --- | --- | --- | --- | --- |
| **Medication** | **Start Date**  dd/mm/yyyy | **Stop Date**  dd/mm/yyyy | **Dose** | **Frequency** |
|  | __ / __ / 202__ | __ / __ / 202__ |  |  |
|  | __ / __ / 202__ | __ / __ / 202__ |  |  |
|  | __ / __ / 202__ | __ / __ / 202__ |  |  |
|  | __ / __ / 202__ | __ / __ / 202__ |  |  |
|  | __ / __ / 202__ | __ / __ / 202__ |  |  |
|  | __ / __ / 202__ | __ / __ / 202__ |  |  |

| **CLINICAL PRESENTATION, TIMING AND PROGRESS** | | |
| --- | --- | --- |
| **ADVERSE EVENT (AE)**: Main Diagnosis:____________________________________ | | |
| Date AE Started: | __ / __ / 202__ |  |
| Date AE Met Serious Criteria: | __ / __ / 202__ |  |
| Date SAE Detected: | __ / __ / 202__ |  |
| Date of Hospitalisation: | __ / __ / 202__ | □ Not applicable |
| Date of Discharge: | __ / __ / 202__ | □ Still in hospital |
| **NARRATIVE**  ________________________________________________________________________  ________________________________________________________________________  ________________________________________________________________________  ________________________________________________________________________  ________________________________________________________________________  ________________________________________________________________________  ________________________________________________________________________  ________________________________________________________________________ | | |

| **RELEVANT MEDICAL HISTORY** - Only complete for the initial SAE report | | | | | | |
| --- | --- | --- | --- | --- | --- | --- |
| **Medical condition** | | Start Date  dd/mmm/yyyy | | Stop Date  dd/mmm/yyyy | | Ongoing |
|  | | __ / __ / 202__ | | __ / __ / 202__ | | □ |
|  | | __ / __ / 202__ | | __ / __ / 202__ | | □ |
|  | | __ / __ / 202__ | | __ / __ / 202__ | | □ |
|  | | __ / __ / 202__ | | __ / __ / 202__ | | □ |
| **INVESTIGATIONS** | | | | | | |
| **Haemoglobin** | **Date** | | **Result** | | **Method** | |
| Pre-treatment | __ / __ / 202__ | | Hb: ___ . __ g/dL | | □CBC □Hemocue □Biosensor | |
| At time of Event Detection | __ / __ / 202__ | | Hb: ___ . __ g/dL | | □CBC □HemoCue □Biosensor | |
| Current | __ / __ / 202__ | | Hb: ___ . __ g/dL | | □CBC □HemoCue □Biosensor | |
| Nadir | __ / __ / 202__ | | Hb: ___ . __ g/dL | | □CBC □HemoCue □Biosensor | |
| **Max Fall in Hb:** | | | | | Hb: __ . __ g/dL | |
| **Maximum Fractional Fall in Hb** | | | | | ____.__ % | |

| **OTHER RELEVENT INVESTIGATIONS** | | | | | |
| --- | --- | --- | --- | --- | --- |
| **G6PD STATUS** - Only complete for the initial SAE report | | | | | |
| **Date of Testing**: __ / __ / 202__ | | | | | |
| **Quantitative Result**: ____ . __ U/g Hb □ Deficient □ Intermediate □ Normal | | | | | |
| **Genotyping**: □ Not done □ Normal □ Variant ______________ | | | | | |
| **LABORATORY TESTS** – If follow up report just add relevant updates | | | | | |
| **Test** | **Date** | | **Result** | | **Other:** |
| **WBC:** | ___ / ___ / 202__ | | _________ x10^9^ | |  |
| **Plt:** | ___ / ___ / 202__ | | _________ | |  |
| **Met Hb:** | ___ / ___ / 202__ | | ____.____ % | |  |
| **Na:** | ___ / ___ / 202__ | | ______ µmol/L | |  |
| **K:** | ___ / ___ / 202__ | | ______ µmol/L | |  |
| **Urea:** | ___ / ___ / 202__ | | ______ µmol/L | |  |
| **Total Bili:** | ___ / ___ / 202__ | | ______ µmol/L | |  |
| **Unconj Bili:** | ___ / ___ / 202__ | | ______ µmol/L | |  |
| **ALP** | ___ / ___ / 202__ | | ______ µmol/L | |  |
| **ALT** | ___ / ___ / 202__ | | ______ µmol/L | |  |
| **LDH:** | ___ / ___ / 202__ | | ______ µmol/L | |  |
| **CLASSIFICATION** | | | | | |
| **SAE**  The reason why classified as Serious | | □ Death: | |  | |
|  |  | □ Life threatening: | | | |
|  |  | □ Hospitalisation or prolongation of hospitalisation: | | | |
|  |  | □ Persistent or significant disability | | | |
|  |  | □ Is a congenital abnormality / birth defect | | | |
|  |  | □ Is an important and significant medical event | | | |
| **Organ Systems Involved** | | □ Haematological □ Gastrointestinal □ Respiratory  □ Cardiological □ Renal □ Liver □ Neurological | | | |
| **Relationship (Causality) to PQ** | | □ Not related □ Unlikely related □ Possibly related  □ Probably related □ Definitely related | | | |

| **CLINICAL MANAGEMENT** | | |
| --- | --- | --- |
| **IV Fluids** | □ Yes □ No |  |
| **Blood transfusion** | □ Yes □ No | Number of units: _____ Date: __/__/202_ |
| **Dialysis** | □ No | □ Peritoneal dialysis □ Haemodialysis |
| **Changes to PQ** | □ No change □ Withheld □ Cease □ Finished □ Restart | |
|  | If Restarted: Date Restarted: __/__/202_  □ Same Dose □ Modified Dose  Dose: ___.__ mg □ 1x Day □ 2x day Duration: ___ Days | |
| **NARRATIVE:**  ________________________________________________________________________  ________________________________________________________________________  ________________________________________________________________________  ________________________________________________________________________  ________________________________________________________________________  ________________________________________________________________________  ________________________________________________________________________  ________________________________________________________________________  ________________________________________________________________________  ________________________________________________________________________  ________________________________________________________________________  ________________________________________________________________________  ________________________________________________________________________  ________________________________________________________________________  ________________________________________________________________________ | | |

| **OUTCOME** | | | | | | |
| --- | --- | --- | --- | --- | --- | --- |
| □ Recovered / Resolved | | | | |  | |
| □ Recovering / Resolving | | | | |  | |
| □ Not recovered / Not resolved | | | | |  | |
| □ Recovered / Resolved with sequelae | | | | | Specify: | |
| □ Fatal: | | Date of death: ___ / ___ / 202__ | | | | |
|  | | Cause of death: | | | | |
| □ Unknown | | | |  | | |
| AE Stopped (Last date AE was present) | | | | ___ / ___ / 202__ | | |
| **NOTIFICATION**  Inform PIs, GMM and MMV within 24 hrs | | | | | | |
| **Time of Notification** | | | Date: __ / __ / 202__ Time: ___ : ___ | | | |
| **Process** | | | □ WhatsApp  □ Email scanned Level 3 Form +/- SAE Form | | | |
| **People Notified** | | | □ Local PI  □ Study MD  □ Menzies / Burnet PI  □ Global Medical Monitor  □ MMV Chief Medical Officer | | | |
| **CLINICIAN RESPONSIBLE FOR THE REVIEW** | | | | | | |
| **Name :** |  | | | | | Date: __ / __ / 202__ |
| **Role :** |  | | | | |  |
| **Address :** |  | | | | |  |
| **Mobile/Whatsapp** : | | | | | | |
| **Email** : | | | | | | |
| **Signature:** | | | | | | |
