## Supplementary data 4 for "High daily dose Short COurse PrimaquinE after G6PD testing for the radical cure of *Plasmodium vivax* malaria in Indonesia and Papua New Guinea: The SCOPE implementation study protocol"

**Supplementary File 4- Adverse Event Gradings**

**Grading for Adverse Events**

Adverse Event Grading the NCI Common Terminology Criteria for Adverse Events (CTCAE vs 5.0) [8].

Grade 1 - Mild: asymptomatic or mild symptoms; clinical or diagnostic observations only; intervention not indicated.

Grade 2 - Moderate: minimal, local or non-invasive intervention indicated; limiting age-appropriate instrumental ADL.

Grade 3 - Severe or medically significant but not immediately life-threatening; hospitalization or prolongation of hospitalization indicated; disabling; limiting self-care ADL.

Grade 4 - Life-threatening: urgent intervention indicated.

Grade 5 - Death related to AE

In addition, the Hillmen Urine colour Chart will be used during Level 3 Review (see table 16 and Level 3 Review Form).

**Adverse Events Grades**

| **Term** | **Grade 1** | **Grade 2** | **Grade 3** | **Grade 4** |
| --- | --- | --- | --- | --- |
| **Abdominal pain** | Mild pain | Moderate pain; limiting instrumental ADL | Severe pain; limiting self-care ADL |  |
| **Nausea** | Loss of appetite without alteration in eating habits | Oral intake decreased without significant weight loss, dehydration or malnutrition | Inadequate oral caloric or fluid intake; tube feeding, or hospitalisation indicated |  |
| **Anorexia** | Loss of appetite without alteration in eating habits | Oral intake altered without significant weight loss or malnutrition; oral nutritional supplements indicated | Associated with significant weight loss or malnutrition | Life-threatening consequences; urgent intervention indicated |
| **Vomiting** | Intervention not indicated | Outpatient IV hydration; medical intervention indicated | Tube feeding, or hospitalisation indicated | Life-threatening consequences |
| **Back Pain** | Mild pain | Moderate pain; limiting instrumental ADL | Severe pain; limiting self-care ADL |  |
| **Breathlessness** | Shortness of breath with moderate exertion | Shortness of breath with minimal exertion; limiting instrumental ADL | Shortness of breath at rest; limiting self-care ADL | Life-threatening consequences; urgent intervention indicated |
| **Dizziness** | Mild unsteadiness or sensation of movement | Moderate unsteadiness or sensation of movement; limiting instrumental ADL | Severe unsteadiness or sensation of movement; limiting self-care ADL |  |
| **Fatigue** | Fatigue relieved by rest | Fatigue not relieved by rest; limiting instrumental ADLs | Fatigue not relieved by rest; limiting self care ADLs |  |
| **Dark Urine** | Hillmen Urine Colour Chart  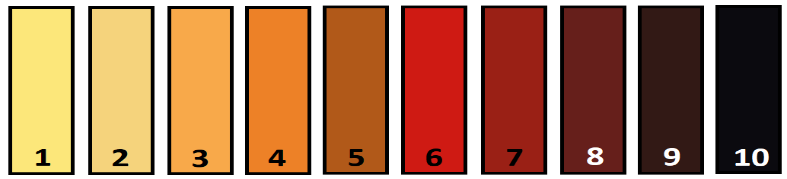 | | | |
