## Supplementary data 5 for "High daily dose Short COurse PrimaquinE after G6PD testing for the radical cure of *Plasmodium vivax* malaria in Indonesia and Papua New Guinea: The SCOPE implementation study protocol"

**Supplementary File 5:**

**SCOPE: Statistical Analysis Plan**

Protocol title: Feasibility of high daily dose short course primaquine after G6PD testing for the radical cure of *Plasmodium vivax* malaria

Short title: **S**hort **CO**urse **P**rimaquin**E** for the radical cure of P. vivax (SCOPE)

Version:	6.0

Authors:	Professor Julie A Simpson

Assoc/Professor Amalia Karahalios

Dr Alistair McLean

Centre for Epidemiology and Biostatistics, and Methods & Implementation Support for Clinical and Health (MISCH) research Hub

University of Melbourne, Melbourne, Australia

Dr Nicholas Douglas

Menzies School of Health Research, Darwin, Australia

Date:	23/10/2024

*This document has been written based on information contained in the SCOPE_PQFS_Master Protocol (version 4.0, 28/04/2023, Protocol Identifying Number - MMV_PQ_MASTER_21_01).*

| **LIST OF ACRONYMS**  **Acronym** | **Definition** |
| --- | --- |
| ACT | Artemisinin-based Combination Therapy |
| ADL | Activity of Daily Living |
| AE | Adverse Event |
| AESI  AMM | Adverse Event of Special Interest  Adjusted Male Median |
| ANZCTR  BRIN  CHW | Australian New Zealand Clinical Trials Registry  *Badan Riset dan Inovasi Nasional* (Indonesia)  Community Health Worker |
| CI | Confidence Interval |
| CQI | Continuous Quality Improvement |
| DFAT | Department of Foreign Affairs and Trade (Australia) |
| FDA | Food and Drug Administration |
| FGD | Focus Group Discussion |
| G6PD | Glucose-6-Phosphate Dehydrogenase |
| G6PDd | G6PD deficiency |
| GI | Gastrointestinal Intolerance |
| GST | Global Study Team |
| HCP | Healthcare Provider |
| MoH | Ministry of Health |
| NDA | National Drug Authority |
| NDoH | National Department of Health |
| NMCP | National Malaria Control Program |
| PCR | Polymerase Chain Reaction |
| PNG | Papua New Guinea |
| PQ | Primaquine |
| PQ7 | Primaquine 1.0 mg/kg/day for 7 days |
| PQ14 | Primaquine 0.5 mg/kg/day for 14 days |
| PQ8W | Primaquine 0.75mg/kg/week for 8 weeks |
| RDT | Rapid Diagnostic Test |
| SAE | Serious Adverse Event |
| SCOPE | **S**hort **CO**urse **P**rimaquin**E** for the radical cure of *P. vivax* |
| SMC | Safety Monitoring Committee |
| SRT  YPKMP | Safety Review Team  *Yayasan Pengembangan Kesehatan dan Masyarakat Papua* (Papua Health and Community Development Foundation) |

### 1 INTRODUCTION

#### 1.1 Preface

Outside of subsaharan Africa, *Plasmodium vivax* is becoming the predominant cause of malaria. Although prevention of relapses is crucial for reducing transmission of this important disease, only two drugs (primaquine and tafenoquine) are licensed for treatment of hypnozoites (the latent liver stages) of *P. vivax* and thus have potential to prevent relapses. Primaquine is the anti-relapse drug available in most endemic regions, but its widespread impact on health outcomes is limited by poor adherence to the standard 14-day course and concerns over the risk of drug-induced haemolysis in patients with glucose-6-phosphate dehydrogenase (G6PD) deficiency. Over the last few years, new tools and treatment strategies have been developed for better tolerated and more effective radical cure regimens. Short-course (7 days), high daily-dose primaquine has been shown to have similar efficacy compared with the same WHO-recommended total dose of 7mg/kg body weight administered over 14 days (Taylor W.J. et al. 2019) in areas with high risk of relapse. However, the higher daily dose in the 7 day regimen (1mg/kg) is associated with more adverse events, particularly an increased risk of haemolysis and gastrointestinal events. A semi-quantitative point-of-care G6PD test (SD Biosensor, ROK) has been developed and registered in Indonesia and the process of registration is underway in Papua New Guinea (PNG).

#### 1.2 Purpose of the analyses

The aim of the SCOPE study is to determine the effectiveness and safety of introducing a revised case management package to facilitate well-tolerated and effective radical cure of patients with vivax malaria at 10 public health care facilities in Indonesia and Papua New Guinea.

The revised case management package has been prioritised and endorsed by the respective Ministries of Health and will include: 1) semi-quantitative point-of-care glucose-6-phosphate dehydrogenase (G6PD) activity assessment prior to treatment with primaquine, 2) prescription of high dose (7mg/kg body weight total) primaquine over 7 (**PQ7**), 14 (**PQ14**) days or 8 weeks (**PQ8W**), to all eligible patients presenting with *P. vivax* malaria, 3) patient education and counselling prior to treatment and supervision of the first dose of primaquine at the health care facility, 4) community-based clinical review at day 3 of primaquine treatment to encourage treatment adherence and facilitate detection and management of adverse events to treatment and 5) improved malariometric surveillance and pharmacovigilance.

### 2 Study Objectives and Endpoints

#### 2.1 Study objectives

The overall objective of the SCOPE study is to determine the operational feasibility and cost-effectiveness of introducing the revised case management package for patients with vivax malaria to inform national antimalarial policy and practice.

This SAP is focused solely on the quantitative measures of operational feasibility, pharmacovigilance, safety and effectiveness in the Stage 1 and Stage 2 studies. A separate document will be prepared for the qualitative measures of operational feasibility and a health economics analysis plan (HEAP) will be prepared for the cost-effectiveness analyses.

##### 2.1.1. Stage 1 – Preliminary safety and feasibility study

**Primary objective:**

S1.PO1	To determine the safety of PQ in patients prescribed different treatment regimens

S1.PO2	Determine the risk of patients experiencing any AESI during treatment

**Secondary objectives:**

S1.SO1	Determine the risk of patients experiencing severe GI AESI during treatment

S1.SO2	Determine the risk of patients experiencing severe haemolysis AESI during treatment

S1.SO3	Determine the risk of patients experiencing severe methaemoglobinaemia AESI during treatment

S1.SO4	Determine the tolerability of PQ in patients prescribed different treatment regimens

S1.SO5	Determine primaquine dosing accuracy by G6PD activity category

S1.SO6	Determine the proportion of patients receiving a clinical review on day 3 and day 7 of PQ treatment

##### 2.1.2. Stage 2 – Large-scale implementation and feasibility study

**Primary objective:**

S2.PO1	To determine the operational feasibility of implementing revised case management for patients with *P. vivax* malaria.

**Secondary objectives:**

*Quantitative measures of operational feasibility*

S2.SO1	Determine the proportion of health care practitioners who comply with the revised radical cure treatment algorithm

S2.SO2	Determine the proportion of patients receiving a SD Biosensor G6PD test

S2.SO3	Determine primaquine dosing accuracy by G6PD activity category

S2.SO4	Determine the proportion of vivax malaria patients who are ineligible for daily primaquine and are incorrectly given primaquine (including infants, pregnant females and G6PD deficient patients)

S2.SO5	Determine the proportion of vivax malaria patients that are reviewed on day 3

S2.SO6	Determine the proportion of vivax malaria patients that adhere to their prescribed primaquine regimen

*Pharmacovigilance*

S2.SO7	Determine the proportion of community-based health workers who correctly act on early signs of haemolytic anaemia and GI events

S2.SO8	Determine the number of patients identified through the community surveillance system with serious adverse events

*Safety*

S2.SO9	Determine the safety of PQ under real-world conditions

S2.SO10	Determine the risk of patients experiencing at least one AESI during treatment

S2.SO11	Determine the risk of patients experiencing severe haemolysis AESI during treatment

S2.SO12	Determine the risk of patients experiencing severe GI AESI during treatment

S2.SO13	Determine the risk of patients experiencing severe methaemoglobinaemia AWSI during treatment

S2.SO14	Determine the difference in severe anaemia prevalence between the pre- and post-implementation periods of the revised case management

*Effectiveness*

S2.SO15	Determine if the incidence of symptomatic *P. vivax* malaria decreases after the introduction of the revised case management

S2.SO16	Determine the difference in prevalence of *P. vivax* parasitaemia in patients presenting with fever between the pre- and post-implementation periods of the revised case management

S2.SO17	Determine if the cumulative risk of representation to the same health facility with symptomatic *P. vivax* malaria within 6 months differs between the pre- and post-implementation periods of the revised case management

#### 2.2 Endpoints

##### 2.2.1. Stage 1 – Preliminary safety and feasibility study

All endpoints refer to patients enrolled into Stage 1 treated with daily or weekly PQ.

**Primary endpoints:**

S1.PE1	Proportion of patients experiencing at least one SAE during treatment.

S1.PE2	Proportion of patients experiencing at least one AESI, SAE or other AE of at least grade 3 during treatment.

**Secondary endpoints:**

S1.SE1	Proportion of patients with a gastrointestinal AESI during treatment.

S1.SE2	Proportion of patients with an AESI related to haemolysis during treatment.

S1.SE3	Proportion of patients with an AESI related to methaemoglobinaemia during treatment.

S1.SE4	Proportion of patients permanently stopping PQ before the end of treatment.

- Defined as the proportion of all patients enrolled who are given documented advice to permanently stop PQ during scheduled or ad hoc clinical review by a health professional

S1.SE5	Proportion of patients receiving correct treatment based on G6PD activity.

S1.SE6	Proportion of patients who were reviewed on day 3 and day 7.

**Exploratory endpoints:**

S1.EE1	Maximum absolute fall in haemoglobin within the first 7 days

S1.EE2	Maximum proportional fall in haemoglobin within the first 7 days

S1.EE3	Reduction of >25% in haemoglobin to a concentration of < 7 g/dl

S1.EE4	*POST HOC:* Level of agreement between SD Biosensor and HemoCue for haemoglobin measurement at study clinics and in the community

S1.EE5	*POST HOC:* Methaemoglobin concentrations on days 3 and 7

##### 2.2.2. Stage 2 – Large-scale implementation and feasibility study

**Primary endpoint:**

S2.PE1	Proportion of patients with *P. vivax* malaria who correctly receive all components of revised case management (including G6PD testing, correct treatment of PQ according to revised case management and G6PD activity, patient education, supervision of the first dose and community review on day 3)

**Secondary endpoints:**

*Quantitative measures of operational feasibility*

S2.SE1	Proportion of health care practitioners who comply with the revised radical cure algorithm as defined below:

- Proportion of health care workers at the SCOPE health facility sites who correctly prescribe PQ (i.e. PQ7, PQ14 or PQ8W) according to the implementation package ≥95% of the time

S2.SE2	Proportion of patients eligible for PQ who are tested for G6PD activity using the SD Biosensor at the time of their diagnosis of malaria.

S2.SE3	Proportion of eligible vivax malaria patients receiving the correct PQ regimen based on the result of the SD Biosensor test.

S2.SE4	Proportion of vivax malaria patients who are ineligible for daily primaquine and are incorrectly prescribed an inappropriate primaquine regimen.

Ineligible being defined as:

- Infant <6 months or <5kg weight (Indonesia) or <1 year (PNG), pregnant women or women breastfeeding infants <6 months of age (Indonesia) or <1 year of age in PNG. Should not receive any primaquine
- G6PD deficient (<30% enzyme activity). Should not receive daily primaquine.

S2.SE5	Proportion of vivax malaria patients that are reviewed on day 3.

S2.SE6	Proportion of vivax malaria patients that adhere to their prescribed primaquine regimen defined as follows:

- The proportion of patients who self-report that they have taken all prescribed PQ doses between the time of the initial prescription and the follow-up visit.

*Pharmacovigilance*

S2.SE7	Proportion of CHWs who correctly act on early signs of haemolytic anaemia and GI events defined as follows:

- Proportion of CHWs who refer 100% of patients documented as having a potential severe symptom (‘flag’) at day 3 review for level 2 clinical review
- Proportion of patients with a severe symptom (‘flag’) at day 3 review who are referred by a CHW for level 2 review

S2.SE8	Number of patients with a SAE who are identified by community or health facility staff follow-up and referred to hospital for further management.

*Safety*

S2.SE9	The proportion of patients eligible to receive PQ who had a serious adverse event (SAE) during treatment.

S2.SE10	The proportion of patients treated with PQ experiencing at least one AESI during treatment.

S2.SE11	The proportion of patients treated with PQ with an AESI related to haemolysis during treatment.

S2.SE12	The proportion of patients treated with PQ with a gastrointestinal AESI during treatment.

S2.SE13	The proportion of patients treated with PQ with AESI related to methaemoglobinaemia during treatment.

S2.SE14	Prevalence of severe anaemia (haemoglobin < 7g/dl) in patients presenting with fever in the Pre-Implementation Survey versus the late Post-implementation Survey (derived from the quantitative surveys).

- Median and distribution of haemoglobin concentrations will also be compared between surveys

*Effectiveness*

S2.SE15	The monthly incidence of confirmed symptomatic *P. vivax* malaria episodes (monoinfection or mixed) in the Pre-Implementation (minimum 6 months) versus Post-Implementation Stage 2 ((a) change in 6 months post implementation (i.e. period between 6 and 12 months post-implementation), (b) change in trend of incidence immediately post-implementation (i.e. entire 12 month period post-implementation)).

S2.SE16	Prevalence of *P. vivax* parasitaemia in patients presenting with fever in the Pre-Implementation Survey versus the Post-Implementation Survey during Stage 2 (derived from the quantitative surveys).

S2.SE17	Cumulative risk of representation to the same health facility with symptomatic *P. vivax* malaria within 6 months in the Pre- and Post-Implementation Stages.

**Exploratory endpoints:**

S2.EE1	Maximum absolute fall in haemoglobin within the first 7 days

S2.EE2	Maximum proportional fall in haemoglobin within the first 7 days

S2.EE3	Reduction of >25% in haemoglobin to a concentration of < 7 g/dl

### 3 Study Methods

#### 3.1 Study design

This is a staged, multicentre, implementation, before-and-after study of a revised case management package for patients with vivax malaria presenting to community health facilities in Indonesia and Papua New Guinea.

The study will be conducted at 10 pre-existing health facilities, 4 in Papua New Guinea (two in stage 1) and 6 in Indonesia (two in stage 1), all of which have significant experience managing patients with vivax malaria.

All of the study health facilities will collect surveillance data that are reported as part of national malaria surveillance systems. These data include patient demographics, diagnosis and treatment. Prior to the start of the project, selected sites have undergone several rounds of continuous quality improvement (CQI) to improve the quality of data collection. Further enhancements to malariometric surveillance and pharmacovigilance will be made as part of the revised case management with a plan to roll these changes out on a wider scale if the changes prove feasible and cost-effective.

The study will be divided into multiple stages (Figure 1). Prior to commencement of Stage 1, enhanced continuous malariometric surveillance and pharmacovigilance will be established at all 10 health facilities. These systems have already been established in 4 sites in PNG and 4 in Papua with more than 12 months of data available. Similar surveillance systems will also be established in North Sumatra and Lampung.

Within the first three months of the SCOPE pre-implementation period, in-depth interviews with key stakeholders and community surveys will be conducted to determine the acceptability, operational feasibility, performance and costs of existing vivax malaria management strategies, as well as baseline *P. vivax* slide or RDT positivity rates. Qualitative, economic and clinical data collection will not necessarily happen simultaneously.

Midway through the Pre-Implementation Stage, consecutive patients (within the bounds of participant accrual limits) presenting to each health facility with fever will be asked to participate in the Pre-Implementation Survey aiming for a total of 200 patients at each facility. Informed consent and assent for participation will be requested. A repeat cross-sectional survey of 200 consecutive patients presenting to each health facility with fever will then be conducted in the final 6 months of the post-implementation period for comparative purposes.

##### 3.1.1. Stage 1 – Preliminary safety and feasibility study

In Stage 1, the feasibility, safety and tolerability of point-of-care G6PD testing and PQ7 in G6PD normal (>70%) patients will be determined in 4 selected health facilities with staff undertaking clinical review on day 3 and also day 7 to ensure patient safety. Stage 2 will only proceed after review of the Stage 1 data by SRT and SMC.

After 3 months, the revised case management will be rolled out at 4 selected sites (2 in Indonesia and 2 in PNG), initiating Stage 1. The remaining sites will continue the Pre-Implementation Stage. Stage 1 is anticipated to last 6 months, during which a total of 800 patients with *P. vivax* malaria (~500 from Indonesia and ~300 from PNG) will be treated (Figure 1).

All patients that are eligible for and consent to participate in the study, will be tested by quantitative G6PD activity assessment using the G6PD STANDARD (SD biosensor, ROK). Patients with G6PD activity >70% (≥6.1 U/gHb) will be prescribed primaquine at a total dose of 7 mg/kg over 7 days (PQ7) and those with 30-70% activity (4.1-6.0 U/gHb) will be prescribed the same total dose of primaquine over 14 days (PQ14). Patients with G6PD activity <30% (≤4.0 U/gHb) will be prescribed primaquine at a dose of 0.75 mg/kg/week for 8 weeks (PQ8w).

The safety review team (SRT) will review all AESI (adverse event of special interest) and SAE (severe adverse event) data after every 100 patients treated with primaquine and may request to review additional data if there are safety concerns. A formal interim analysis is planned after enrolment of 400 patients (~150 patients from PNG, ~250 patient from Indonesia). The SRT will review the interim analysis outputs and share with the Safety Monitoring Committee (SMC) who will conduct an interim review and issue a recommendation regarding continuation of Stage 1.

At the end of Stage 1, once 800 patients (~300 patients from PNG, ~500 patients from Indonesia) have been enrolled, a final analysis will be conducted and reviewed by the SRT and presented to the Safety Monitoring Committee. We anticipate that 700 patients will have been treated with PQ7 and 100 with PQ14. Prior to study start the SMC will be tasked with defining specific *a priori* criteria that will be used to decide whether the study progresses directly to the main feasibility study for wider roll out in Stage 2 without changes in the protocol, whether the implementation package needs to be revised or if the study needs to be discontinued. The final analysis of Stage 1 will be reviewed by the SRT and summarized to support the SMC final review. If the SMC recommends progression to Stage 2, the data summary, together with the SMC recommendations will be shared with the NDoH and NDA for approval to progress to Stage 2.

##### 3.1.2. Stage 2 – Large-scale implementation and feasibility study

Stage 2 of the study will be a feasibility study conducted at 10 community health-care facilities: 6 sites in Indonesia and 4 sites in Papua New Guinea. The primary purpose of Stage 2 is to determine the feasibility and cost-effectiveness of the complete revised case management initially studied in Stage 1. Robust Pre-Implementation malariometric surveillance for a minimum of 12 months along with quantitative surveys of baseline operational metrics and costs will be followed by coordinated introduction of the revised case management across all study health facilities and a minimum of a further 12 months of surveillance – creating an interrupted time series comparing the endpoints listed in section 2.2.2.

During Stage 2 study procedures will be the same as those for Stage 1, except that the procedures will be performed by routine healthcare staff. All patients will be reviewed by community health workers (CHW) at home 3 days after starting PQ. The review may also take place at the health facility, if the patient presents there. Exceptionnally, the follow-up may also take place via a phone call if the patient is not physically reachable at home. The CHW will complete a signs and symptom checklist to detect early indicators of haemolysis or other AESI. Patients with signs of haemolysis or other AESI will have their primaquine ceased and will be referred to the health facility for medical review. Patients tolerating their medication will be encouraged to adhere to a full (7 or 14 days) course of treatment. Safety outcomes occuring between day 3 and end of treatment will be detected passively by community-based health workers, study health facilities and regional hospitals.

Continuous surveillance and pharmacovigilance over the entire study period will enable interrupted time-series analyses of malariometric indices (see section 2.2.2).

Figure 1 presents the study schedule for the baseline data, pre-implementation stage, and stages 1 and 2.

**Figure 1. Study overview**

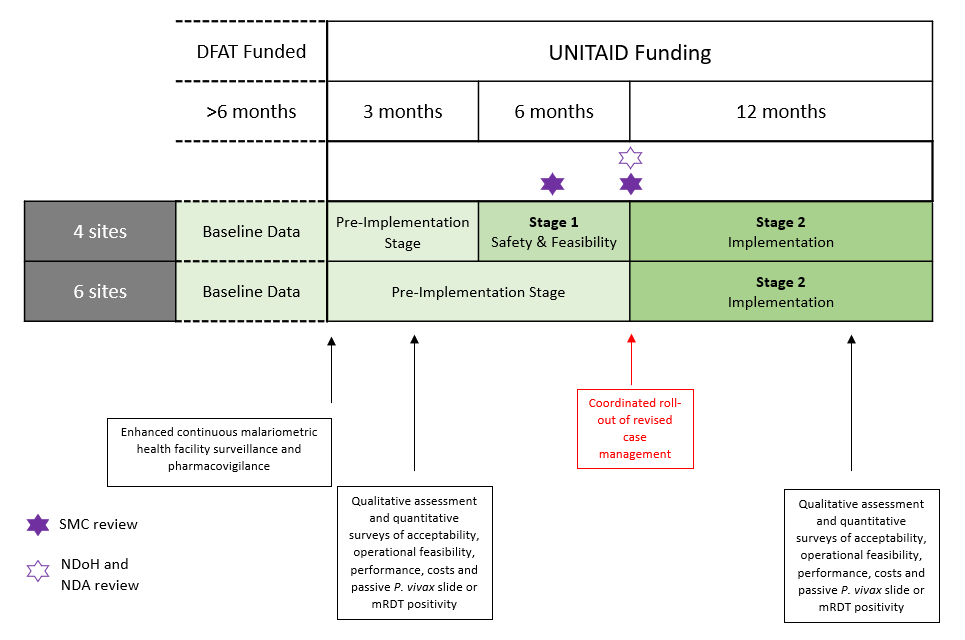

#### 3.2 Inclusion and exclusion criteria

**Inclusion criteria Stage 1 of the study (and thus receipt of either PQ7, PQ14 or PQ8w) are**: age >1 year (PNG) or ≥6 months (Indonesia), haemoglobin concentration ≥8 g/dl and signed agreement of patient or their carers to participate in the study and comply with all study-specific obligations, including attending follow-up clinical evaluations (at day 3 and day 7). Inclusion criteria for Stage 2 will be the same except that patients will only need to be able to attend a follow-up appointment (or receive a phone call) on day 3.

**Exclusion criteria**: pregnant and lactating females with infants <6 months in Indonesia and <1 year in PNG and patients with severe manifestations of malaria will not be prescribed primaquine and will not be enrolled in Stage 1. These patients will contribute incidence data to Stage 2 of the study but will not receive primaquine treatment. Patients’ ineligible for primaquine treatment will be treated according to current national guidelines.

### 4 Sample size

#### 4.1 Stage 1

In the Stage 1 preliminary study of the feasibility and safety of the implementation package, 4 health facilities will enrol 800 patients (~500 in Indonesia and ~300 in Papua New Guinea). Assuming 764 of these patients will have a G6PD activity >70% and be eligible for PQ7, and that the proportion of patients receiving short-course primaquine who have to stop therapy within 7 days is 5%, this sample size will produce a 95% confidence interval for the true proportion needing to stop treatment of +/- 1.55%.

#### 4.2 Stage 2

***NOTE:*** *Given slower than expected recruitment in Stage 1, the predicted patient recruitment in Stage 2 was revised down on 5 August 2024 to 70% of eligible patients (n=11,410) across Indonesia and Papua New Guinea. Additionally, an error was detected in the original power calculation for detection of a reduction in incidence of vivax malaria. In protocol vs 5.0, the total number of patients with vivax malaria recruited into the study was erroneously predicted to be 16,300 over 18 months (the initial timeframe for the study) when in reality this was the number of patients predicted over 12 months. This resulted in an underestimate of the monthly case incidence at each clinic and thus an underestimate of the power available for detection of an incidence reduction.*

*The revised and corrected power and precision calculations are provided below with the original erroneous calculations retained for reference. Power calculations for detection of an individual-level reduction in risk of vivax malaria recurrence are also provided.*

##### 4.2.1 Revised and corrected Stage 2 power and precision calculations

Assumptions underlying the following Stage 2 power calculations include the following:

1. 5% of patients are ineligible for the intervention package (pregnancy, lactation, severe malaria, age <6m (Indonesia) or <1 yr (PNG).
2. The impact on the risk and rate of infection will be the same across different endemic sites.
3. The mean baseline risk of *P. vivax* recurrence at 6 months is 20%
4. Loss to follow-up in both groups is 10%
5. Alpha = 0.05

During the 12-month pre-implementation period, a total of **17,160** patients with *P. vivax* (mono or mixed infections) presented to the ten study clinics (see Table 1). Assuming 5% of patients are ineligible for enrolment due to pregnancy, lactation or age under 6 months (Indonesia) or 12 months (PNG), this gives an estimated maximum number of patients eligible for recruitment into Stage 2 of **16,300**.

**Table 1 Number of patients with vivax malaria attending the ten study clinics in last 12 months**

| **Clinic name** | **Country** | **Locality** | **Estimated total number of *P. vivax* episodes per year** | **Estimated maximum recruitment in SCOPE Stage 2**  **(95%)** |
| --- | --- | --- | --- | --- |
| **Timika** | Indonesia | Timika, Southern Papua | 3173 | 3014 |
| **Bhintuka** | Indonesia | Timika,  Southern Papua | 1164 | 1106 |
| **Pasar Sentral** | Indonesia | Timika,  Southern Papua | 4937 | 4690 |
| **Wania** | Indonesia | Timika,  Southern Papua | 3681 | 3497 |
| **Tanjung Leidong** | Indonesia | North Sumatra | 500 | 475 |
| **Hanura** | Indonesia | Lampung | 450 | 428 |
| **Baro** | PNG | West Sepik | 417 | 396 |
| **Mugil** | PNG | Madang | 1044 | 992 |
| **Napapar** | PNG | East New Britain | 412 | 391 |
| **Wirui** | PNG | East Sepik | 1382 | 1313 |
|  |  | Total | 17,160 | **16,302** |

Footnotes: Estimates based on previous clinic records and number of patients with *P. vivax* recruited over last 12 months. Maximum enrolment assumes 5% of patients are ineligible because they are pregnant, lactating or under 6-12 months of age.

**S2.SE15 - Incidence Rate Reduction**

The proportion of patients recruited into SCOPE has been lower than expected and this will impact on the predicted reduction in incidence. The revised estimated reduction in incidence and predicted power to show this are provided in Table 2, with varying levels of recruitment (0.5-1).

**Table 2 – Power to detect a reduction in the incidence of *P. vivax* malaria**

| **Proportion of eligible *P. vivax* malaria patients recruited** | **Eligible patients over 12 months period** | **Predicted mean monthly incidence at each clinic preintervention** | **Predicted absolute incidence reduction** | **Predicted mean monthly incidence at each clinic post intervention** | **Power to detect predicted incidence reduction** |
| --- | --- | --- | --- | --- | --- |
| 1 | 16,300 | 135.8 | 24.44 | 111.36 | 99.5% |
| 0.9 | 14,670 | 135.8 | 22.00 | 113.80 | 98.4% |
| 0.8 | 13,040 | 135.8 | 19.56 | 116.24 | 95.5% |
| 0.7 | 11,410 | 135.8 | 17.11 | 118.69 | **89.3%** |
| 0.6 | 9,780 | 135.8 | 14.67 | 121.13 | 78.6% |
| 0.5 | 8,150 | 135.8 | 12.22 | 123.58 | 63.2% |

**Example STATA code**: power pairedm 135.8 111.36, sddiff(15) n(10) alpha(0.05).

Based on previous estimates, 60% of cases of vivax malaria seen at each clinic are due to relapse and therefore preventable with improved radical cure. We predict the intervention package will reduce the individual risk of relapse by 30%. With anticipated recruitment of 70% of eligible patients, the total predicted sample size in Stage 2 would fall to **11,410** patients receiving primaquine over 12 months of enrolment. Based on this lower predicted enrolment and using the assumptions as above, the predicted absolute reduction in incidence at each clinic would be 17.11 cases per month ((135.8*0.6)*0.3*0.7) and resultant power to detect this difference would be **89.3%.**

**S2.SE17 - Reduction in individual risk of recurrence**

The power estimates for detecting a reduction in the individual risk of recurrence of *P. vivax* within 6 months from an anticipated study enrolment of **11,410 patients** are presented in Table 3. The number of patients included in the pre-implementation and implementation stages are assumed to be equal (ie a total of 22,820 patients pre and post implementation).

**Table 3 – Power to detect reduction in the risk of recurrence**

| **Proportion of eligible *P. vivax* malaria patients recruited** | **Eligible patients over 12 months period** | **30% Risk Reduction**  **(HR 0.7)** | **20% Risk Reduction**  **(HR 0.8)** | **10% Risk Reduction**  **(HR 0.9)** |
| --- | --- | --- | --- | --- |
| 1 | 16,300 | 100% | 100% | 98% |
| 0.9 | 14,670 | 100% | 100% | 97% |
| 0.8 | 13,040 | 100% | 100% | 95% |
| 0.7 | 11,410 | 100% | 100% | **92%** |
| 0.6 | 9,780 | 100% | 100% | 88% |
| 0.5 | 8,150 | 100% | 100% | 82% |

**Example STATA code**: artsurv, edf0(0.2) hr (1,0.7) fp(1) n(22,820) alpha(0.05) nperiod(1) tunit(2) lg(1 2) ldf(0.1;0.1).

A sample size of 11,410 (70% eligible patients) would provide **92% power** to detect a minimum 10% reduction in risk of recurrence and 100% power to detect a 30% reduction in risk of recurrence.

**S2.PE1 Proportion of patients with *P. vivax* malaria who correctly receive all components of revised case management**

Assuming 70% of patients enrolled into Stage 2 receive all components of the implementation package correctly, the revised Stage 2 sample size of 11,410 patients will give a 95% confidence interval for the true proportion of +/- 0.84%.

**S2.SE9 Safety and protocol adherence outcomes**

Assuming 1% of patients have a treatment-limiting haemolytic event, the revised Stage 2 sample size of 11,410 patients will give a 95% confidence interval for the true proportion of +/- 0.18%.

##### 4.2.2 Original power and precision calculations – now superseded

**Indonesia and PNG combined study**

The Stage 2 implementation study will be conducted at ten health facilities across Indonesia and Papua New Guinea. Based on prior surveillance, we estimate that over 18 months, these health facilities will enrol approximately 16,300 patients with vivax malaria. Improved radical cure of *P. vivax* malaria is expected to reduce the risk of subsequent relapse and therefore reduce the overall incidence of vivax malaria. Across the 10 health facilities, a mean of 91 patients with vivax malaria are expected to be seen each month, 60% (n=54.3) of which are predicted to be relapses and therefore feasibly preventable. Assuming that the implementation package in Stage 2 will reduce relapses by 30%, the monthly incidence of *P. vivax* malaria is expected to fall by 16.3 (17.9%) to 74.7 cases per month. With a two-sided alpha of 0.05 and a predicted standard deviation of the mean variation in monthly numbers of vivax malaria of 15 episodes per month, our study would achieve 86% power to detect an 18% reduction in monthly *P. vivax* malaria incidence. Furthermore, assuming 70% of patients receive all components of the implementation package correctly, this sample size will give a 95% confidence interval for the true proportion of +/- 0.70%. If 1% of patients are predicted to have a treatment-limiting haemolytic event, this sample size will be powered to determine the true proportion within +/- 0.15%.

**Indonesia and PNG standalone studies**

In the unlikely scenario that either Indonesia or PNG will not proceed, the study will become a standalone country study. In this case the sample size calculations are as follows. In a standalone Indonesia study, Stage 1 will occur in the same two health facilities as outlined above. However, we will enrol 800 patients to retain the sample size of the combined study. Stage 1 will still produce a 95% confidence interval for the true proportion needing to stop treatment of +/- 1.55%. In Stage 2, the implementation study will be conducted at six health facilities in Indonesia. Based on prior surveillance, we estimate that over 12 months, these health facilities will enrol approximately 10,750 patients with vivax malaria. Across the 6 health facilities, a mean of 60 patients with vivax malaria are anticipated each month, 60% (n=36) of which are predicted to be relapses and are therefore preventable. Assuming that the implementation package in Stage 2 will reduce relapses by 30%, the monthly incidence of *P. vivax* malaria is expected to fall by 10.7 (17.9%) to 49.0 cases per month. The Indonesian study alone would retain 80% power to detect a 36% overall reduction in incidence following implementation of the interventions. Furthermore, assuming 70% of patients receive all components of the implementation package correctly, this sample size will give a 95% confidence interval for the true proportion of +/- 0.87%. If 1% of patients are predicted to have a treatment-limiting haemolytic event, this sample size will be powered to determine the true proportion within +/- 0.19%.

In PNG as a standalone study, Stage 1 will occur in two health facilities, as outlined above, however we will enrol 800 patients to retain the sample size of the combined study. Stage 1 will still produce a 95% confidence interval for the true proportion needing to stop treatment of +/- 1.55%. In Stage 2, the implementation study will be conducted at four health facilities in PNG. Based on prior surveillance, we estimate that over 12 months, these health facilities will enrol approximately 5,550 patients with vivax malaria. Improved radical cure of *P. vivax* malaria is expected to reduce the risk of subsequent relapse and therefore reduce the overall incidence of vivax malaria. Across the 4 health facilities, a mean of 77 patients with vivax malaria are anticipated each month, 60% (n=46.3) of which are predicted to be relapses and are therefore preventable. Assuming that the implementation package in Stage 2 will reduce relapses by 30%, the monthly incidence of *P. vivax* malaria is expected to fall by 13.8 (17.9%) to 63.2 cases per month. The PNG arm of the study would retain 80% power to detect a 42% overall reduction in incidence following implementation of the interventions. Furthermore assuming 70% of patients receive all components of the implementation package correctly, this sample size will give a 95% confidence interval for the true proportion of +/- 1.21%. If 1% of patients are predicted to have a treatment-limiting haemolytic event, this sample size will be powered to determine the true proportion within +/- 0.26%.

Results of the power calculations for the effectiveness endpoints provided above have been confirmed using numerical simulations. A total of 1000 simulated datasets from 10 health facilities, with 18 months of observations and 91 infections per month (in the pre-intervention phase) were generated. Between health-facility variation in the outcome was based on an intra-cluster correlation coefficient of 0.1, and within health-facility variation was assumed to have a standard deviation of the mean variation in monthly numbers of vivax malaria of 15 episodes per month.

### 5 Analysis Objectives

#### 5.1. Stage 1 – Preliminary safety and feasibility study

The analysis objectives for Stage 1 are to estimate the proportion of patients with the primary and secondary endpoints (i.e. safety, tolerability and clinical review endpoints) provided in section 2.2.1.

Following accrual of anecdotal evidence of some seemingly erratic haemoglobin results from the SD Biosensor during the early phase of the Stage 1 study, a decision was made to test haemoglobin in duplicate using the bedside HemoCue assay, where available, to ensure patient safety and to provide a means of assessing the reliability of the SD Biosensor. A relevant *post hoc* exploratory endpoint (S1.EE4) has been added to this Statistical Analysis Plan and details of analytical methods are provided below.

Methaemoglobin concentration on days 3 and 7 has recently been proposed as a surrogate marker of primaquine anti-hypnozoite activity (Fadilah I. et al. 20204). Following two reports of adverse events in Papua (low oxygen saturation and elevated methaemoglobin), methaemoglobin began to be measured routinely on days 3 and 7 in Papua midway through Stage 1. Methaemoglobin concentration has therefore been added as a relevant *post hoc* exploratory endpoint (S1.EE5).

#### 5.2. Stage 2 – Large-scale implementation and feasibility study

The primary analysis objective for Stage 2 is to estimate the proportion of patients with *P. vivax* malaria who correctly receive all components of revised case management (including G6PD testing, correct treatment with primaquine according to revised case management and G6PD activity, patient education, supervision of the first dose and community review at day 3), as stated in section 2.2.2. The secondary analysis objectives for Stage 2 include estimating the proportion of patients with secondary safety, tolerability and clinical review endpoints. The effectiveness and safety of implementing revised case management for patients with *P. vivax* malaria will also be estimated using an interrupted time series design for the incidence of symptomatic *P. vivax* malaria episodes, survival analysis of the cumulative risk of representation with *P. vivax* malaria and before versus after analyses of the prevalence of asexual *P. vivax* parasitaemia and severe anaemia in patients presenting to health facilities with fever.

All of the above estimates will be presented with 95% confidence intervals.

### 6 Analysis sets/Subgroups

#### 6.1. Stage 1 – Preliminary safety and feasibility study

The primary analyses of the endpoints in Stage 1 will include data from all patients enrolled across the four sites in PNG and Indonesia. Secondary analyses of patient-related study outcomes will be conducted overall and for the following prespecified subgroups:

- Patients from Indonesia, Papua New Guinea
- Study site
- Ethnic group
- Patients with G6PD activity >70%, ≥30% to ≤70% and <30%
- Patients prescribed PQ7, PQ14 and PQ8W
- Patients aged <5 years, 5-15 years and >15 years
- Males and females

#### 6.2. Stage 2 – Large-scale implementation and feasibility study

The primary analysis of the endpoints in Stage 2 will include all patients enrolled across the 10 health facilities in Indonesia and Papua New Guinea from the start of the pre-implementation period to the end of the post-implementation period (estimated to be about 16,300 patients). Before versus after analyses of the prevalence of asexual *P. vivax* parasitaemia and severe anaemia from the cross-sectional surveys will be done on the full aggregated dataset of all patients enrolled in the surveys across the 10 study sites (aiming for 200 patients each health facility for each survey = 4,000 patients total).

Secondary analyses of patient-related study outcomes will be conducted for the following pre-specified subgroups:

- Patients from Indonesia, Papua New Guinea, Timika and Sumatra
- Study site
- Ethnic group
- Patients with G6PD activity >70%, ≥30% to ≤70% and <30%
- Patients prescribed PQ7, PQ14 and PQ8W
- Patients aged <5 years, 5-15 years and >15 years
- Males and females

All of the health facilities in the study are already collecting *P. vivax* malaria incidence data. A sensitivity analysis of the change in incidence of symptomatic *P. vivax* malaria episodes will be done by including incidence data for up to three years prior to the start of the SCOPE study to capture a more-prolonged baseline.

### 7 Outcomes and Covariates

The primary, secondary and exploratory outcomes for Stage 1 and Stage 2 are defined in Section 2.2 (Endpoints).

Intervention and covariate variables for analyses of the impact of the implementation package on incidence of symptomatic *P. vivax* malaria (effectiveness outcome S2.SE15) will be defined as follows:

a) **Level change** intervention effect (i.e. change in median monthly incidence) 6 months after the start of Stage 2 (initiation of the implementation package); Figure 2A, Section 9.

- Intervention will take the value of 1 for all time points ≥6 months after the start of Stage 2 at each health facility site, and a value of 0 for all other timepoints.

b) **Slope change** intervention effect (i.e. change in the rate of change of monthly incidence) immediately after the start of Stage 2 (initiation of the implementation package); Figure 2B, Section 9.

- Intervention will take the value of 1 for all time points following the start of Stage 2 within each health facility site, and a value of 0 for all other timepoints.

The time month covariate will be defined as the number of months after the start of Stage 2, i.e. negative values for pre-implementation and positive values for post-implementation.

Seasonality will be captured using spline terms for the Indonesian health facility sites and the PNG health facility sites.

Any unexpected stock outages or major, discrete malaria control interventions external to the SCOPE project will also be captured and reported by health facility site and calendar time month.

The key covariates that will be included in the analyses of the safety and exploratory outcomes will be the haemoglobin concentration at enrolment, the mg/kg dose of primaquine and the G6PD enzyme activity.

### 8 Handling of Missing Values

We do not expect there to be any reason that calendar time month will be missing for any observations over the study period (pre-implementation, Stage 1 and Stage 2) at each of the 10 sites. This is based on our experience of routine data collection during the baseline data period (i.e. prior to pre-implementation stage). We will descriptively report the patterns of missingness for the outcome measures and other participant variables over time in each site. For all analyses, missing data will be handled using an available case analysis, i.e. missing data on any variable included in the analysis where the observation for an individual at that time point is missing is excluded from the analysis.

### 9 Statistical Methodology

Continuous surveillance and pharmacovigilance data will be summarised and presented to study staff in graphical and descriptive text format on a monthly basis to enable real-time feedback on case numbers and distributions, data integrity, clinical performance and revised case management adherence.

#### 9.1. Stage 1 – Preliminary safety and feasibility study

For Stage 1, the number and proportion of patients experiencing at least one SAE (S1.PE1) or at least one AESI (S1.PE2) will be computed by treat ment arm (PQ7, PQ14, PQ8w), and described in summary tables which will include breakdown by type of event, severity, relatedness to treatment, and timing of occurrence (see Appendix 4 for example Tables 1-4). The number and proportion of patients permanently discontinuing treatment (S1.SE4) will be computed by treatment arm and presented in a summary table, with breakdown of reason for discontinuation. Absolute numbers of AESIs with denominators (S1.PE1-3) and proportions with 95% confidence intervals (calculated using a binomial distribution) will be presented in tabular and graphical format. A full tabulation of all SAEs will also be presented.

For S1.EE3, the number and proportion (and 95% Confidence Interval) of patients observed to have a >25% reduction in haemoglobin to a concentration below 7 g/dl will be computed by treatment arm (PQ7, PQ14, PQ8w), and described in summary tables.

For S1.EE1 and S1.EE2, the unadjusted and adjusted mean absolute and proportional maximum change in haemoglobin within 7 days (95% CI) respectively will be presented by treatment arm (PQ7, PQ14, PQ8w). The adjusted means will be estimated from a linear regression model that adjusts for baseline haemoglobin level and G6PD enzyme activity level. The association between change in haemoglobin from baseline to day 2 and G6PD enzyme activity level will be estimated from a linear regression model that allows for a nonlinear association using fractional polynomials and adjusts for baseline haemoglobin level, age, sex and baseline parasitaemia. A scatterplot of the data will be presented with observations labelled by treatment arm (PQ7, PQ14, PQ8w).

For S1.EE4, the mean difference, standard deviation of the difference and 95% limits of agreement between Biosensor and HemoCue haemoglobin measurements will be computed using Bland-Altman plots (Bland JM & Altman DG 1995), stratified by day of measurement (day 0, 3, 7) and country. The data will be presented in tabular and graphical formats.

For S1.EE5, the distribution of methaemoglobin at baseline, day 3 and day 7 will be presented by treatment arm (PQ7, PQ14, Pq8w) using box and whisker plots. The distribution of methaemoglobin levels at day 7 will be compared between the PQ7 (daily PQ dose of 1mg/kg) and PQ14 (daily PQ dose of 0.5 mg/kg) treatment arms using linear regression after adjusting for baseline haemoglobin concentration, parasitaemia, age, sex, and G6PD enzyme activity level.

#### 9.2. Stage 2 – Large-scale implementation and feasibility study

For Stage 2, the proportion of eligible patients receiving all elements of the implementation package correctly, and other related outcomes, will be presented for all patients, as well as stratified by study country and health facility. Corresponding 95% confidence intervals of these outcomes will be calculated assuming a binomial distribution.

The change in the incidence of symptomatic *P. vivax* malaria episodes (S2.SE15 - effectiveness endpoint) between the Pre-Implementation Stage and post-implementation stage (Stage 2) will be shown graphically using an interrupted time series graph. Following the recommendations of Turner et al. (2020) the following components will be displayed: the health facility site-level (aggregated) data points versus time point of measurement (e.g. monthly for primary outcome) as a scatter plot; the interruption time will be displayed with a vertical dashed line; and the pre-interruption and post-interruption trend lines; and the counterfactual trend line (an example is given in Appendix 4, Figure 2).

The estimate of the effect of the implementation package will be determined by fitting a segmented regression model to the individual level data (Bernal J.L. et al. 2017). For the segmented regression model, we will estimate the impact of the revised case management as:

a) a level change comparing six months post-implementation to pre-implementation stage (Figure 2A) with a binary indicator variable that takes the value of 1 for all time points 6 months after the start of Stage 2 (initiation of the implementation package) within each health facility site, and a value of 0 for all other timepoints.

b) the difference in trends before and after introduction of the implementation package (i.e. slope change comparing post-implementation to pre-implementation stage) (Bernal J.L. et al. 2017) (Figure 2B) with a binary indicator variable that takes the value of 1 for all time points following the start of the implementation package within each health facility site, and a value of 0 for all other timepoints.

The model will be fitted using restricted maximum likelihood (REML) to account for autocorrelation and will include time (month) as a continuous variable, a binary variable for the implementation intervention (see (a) and (b) above), an interaction between elapsed time (since implementation) and the implementation, seasonal variation in malaria (captured using splines), adjusted for potential time-varying confounding variables including large scale malaria control activities outside of the SCOPE study and unexpected antimalarial stock outages, and using robust standard errors to account for clustering by health facility site (Turner SL et al. 2021).

For the effectiveness outcome S2.SE15, monthly incidence of symptomatic *P. vivax* malaria will be calculated using clinic catchment populations provided by health facilities (or if not available, the most recent census). Incidence rate ratios (IRRs) (95% CIs) for the intervention effect will be estimated from a negative binomial regression model fitted to the monthly aggregated data.

**Figure 2.** Example impact models for the implementation package on the effectiveness outcome measure S2.SE15 (incidence of symptomatic *P. vivax* malaria infections). (A) An impact model with a lagged change in level that occurs six months after the start of Stage 2 (‘Level Change’). (B) An impact model with an immediate change in slope after the implementation package (‘Slope Change’).

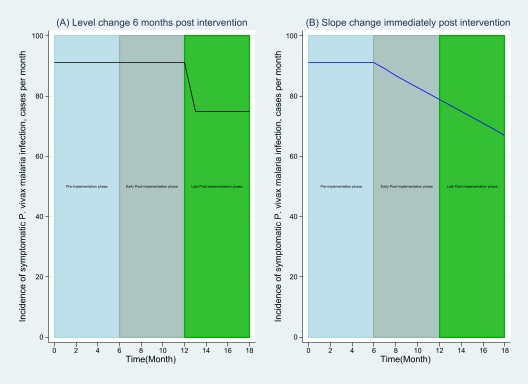

For comparison of outcomes S2.SE16 (the proportion of febrile patients with asexual *P. vivax* parasitaemia by microscopy) and S2.SE14 (the proportion of patients with severe anaemia (haemoglobin < 7 g/dl), determined from cross-sectional surveys in the pre- and post-implementation periods), prevalence ratios (95% CIs) for the intervention effect will be estimated from log-binomial regression models, and if there are convergence issues, Poisson regression with robust variance will be performed (Barros AJD & Hirakata VN 2003).

Individual patient data on the risk of representation with vivax malaria within 6 months to the same health facility site (S2.SE17) will be analysed using survival analysis and visualised with Kaplan-Meier curves. The risk of recurrence will be determined for all patients eligible for PQ treatment and compared between the pre- and post-implementation stages using log rank tests. Multivariable Cox proportional hazards regression will be used to compare these same groups, controlling for age, sex, health facility site, calendar time (month) and seasonality (captured using splines). The validity of the proportional hazards assumption will be assessed visually using Schoenfeld residuals and stratified analyses will be conducted where the assumption is violated. Both Kaplan-Meier and Cox proportional hazards regression analyses will be repeated for the subgroup of patients who are eligible for daily primaquine (i.e. excluding patients with G6PD activity <30%).

For S2.EE3, the number and proportion (and 95% Confidence Interval) of patients observed to have a >25% reduction in haemoglobin to a concentration below 7 g/dl will be computed by treatment arm (PQ7, PQ14, PQ8w), and described in summary tables.

For S2.EE1 and S2.EE2, the unadjusted and adjusted mean absolute and proportional maximum change in haemoglobin within 7 days (95% CI) respectively will be presented by treatment arm (PQ7, PQ14, PQ8w). The adjusted means will be estimated from a linear regression model that adjusts for baseline haemoglobin level and G6PD enzyme activity level.

**Clustering**

For all outcomes captured at the individual patient level the relevant hierarchy of observations is that observations over time are clustered within individuals and individuals are clustered within village. This clustering will be explored in analyses of the effectiveness outcomes using random effects with individuals nested within villages, in addition to the analyses described above that include robust standard errors for clustering by health facility.

#### 9.3 Measures to Adjust for Confounders, Heterogeneity and Autocorrelation

**Confounders**

Both pre- and post-implementation surveillance periods will be of sufficient duration to minimise the risk of outlier seasonal and social events on the incidence of vivax malaria. Multivariable models, controlling for known potential confounders such as sex, age, and seasonality will be used for comparisons of malaria incidence, slide positivity and other indices. For the exploratory outcomes, S1.EE1, S1.EE2, S2.EE1 and S2.EE2, multivariable linear regression models will be adjusted for known potential confounders, baseline haemoglobin levels and G6PD enzyme activity level.

**Heterogeneity**

The mixed effects models allow for some heterogeneity in slope and intercept at the individual and village level. A visual assessment of heterogeneity of intervention effect according to site characteristics will be performed.

**Autocorrelation**

Autocorrelation for the segmented mixed-effects regression analyses estimating the effectiveness of the implementation package will be assessed visually by plotting the residuals (difference between observed and model predicted outcome) over time for each model.

### 10 Sensitivity Analyses

The following sensitivity analyses will be performed for the analysis of the S2.SE15 effectiveness outcome (incidence symptomatic *P. vivax* malaria episodes);

- The analysis will include incidence data collected up to three years prior to the start of the pre-implementation phase.
- The analysis will exclude the incidence data from the 4 health facilities participating in Stage 1 from the pre-implementation period
- The analysis will be performed with alternative adjustments for seasonality
  - Adjustment with a categorical rainy/dry seasonality variable derived specifically for each site

### 11 Quality Control Plans

Details of data cleaning and verification, including consistency and range checks, will be specified in a Data Management Plan.

### 12 References

1. Taylor WRJ, Thriemer K, von Seidlein L, Yuentrakul P, Assawariyathipat T, Assefa A, et al (2019). Short-course primaquine for the radical cure of Plasmodium vivax malaria: a multicentre, randomised, placebo-controlled non-inferiority trial. The Lancet **394**(10202):929-38
2. Fadilah I, Commons RJ, Chau NH, Chu CS, Day NPJ, Koh G, et al (2024). Methaemoglobin as a surrogate marker of primaquine antihypnozoite activity in Plasmodium vivax malaria: A systematic review and individual patient data meta-analysis. PLoS Med **21**(9):e1004411
3. Barros AJD, Hirakata VN (2003). Alternatives for logistic regression in cross-sectional studies: an empirical comparison of models that directly estimate the prevalence ratio. BMC Med Res Methodol **3:** 21.
4. Bland JM, Altman DG (1995). Comparing methods of measurement: why plotting difference against standard method is misleading. The Lancet **346**(8982):1085-7.
5. Bernal, J.L., S. Cummins, and A. Gasparrini (2017). Interrupted time series regression for the evaluation of public health interventions: a tutorial. Int J Epidemiol **46**(1): p. 348-355.
6. Bazo-Alvarez, J. C., T. P. Morris, T. M. Pham, J. R. Carpenter and I. Petersen (2020). Handling Missing Values in Interrupted Time Series Analysis of Longitudinal Individual-Level Data. Clin Epidemiol **12**: 1045-1057.
7. Turner, S. L., A. Karahalios, A. B. Forbes, M. Taljaard, J. M. Grimshaw, E. Korevaar, A. C. Cheng, L. Bero and J. E. McKenzie (2021). Creating effective interrupted time series graphs: Review and recommendations. Research Synthesis Methods **12**(1): 106-117.
8. Turner S.L., et al. (2021). Evaluation of statistical methods used in the analysis of interrupted time series studies: a simulation study. BMC Med Res Methodol **21**(1): p. 181.

### Appendix 1

**VARIABLES IN THE DATA SET**

| **Variable in data set** | **Units** | **Variable label** |
| --- | --- | --- |
| **Baseline/Enrolment CRF** | | |
| scopeid | N/A | SCOPE Study ID |
| country | N/A | Country |
| staffid | N/A | Staff Identification Number |
| studysite | N/A | Study Site |
| visitdat | N/A | Visit Date |
| mcard | N/A | Indonesia: Malaria Card Number  PNG: Malaria ID |
| initials | N/A | Patient Initials |
| sex | N/A | Sex |
| age | years | Age (years) |
| agemonths | months | Age (months) |
| weight | kg | Weight |
| pregnant | N/A | Pregnant |
| lactating | N/A | Indonesia: Lactating < 6m  PNG: Lactating (infant younger than 12 months) |
| village01 | NA | Village of residence |
| village02 | NA | Village of residence |
| ethnic | N/A | Indonesia: Ethnic Group  PNG: Ethnicity |
| ethnichigh | N/A | Highland Papuan |
| ethniclow | N/A | Lowland Papuan |
| ethnicoth | N/A | Other Papuan (specify) |
| ethnicnon | N/A | Non-Papuan (specify) |
| diagnostic | N/A | Diagnostic |
| ringform | N/A | Ringform |
| gametocyte | N/A | Gametocyte |
| diagnosis_ms | N/A | Diagnosis (Microscopy) |
| diagnosis_rdt | N/A | Indonesia: Diagnosis (RDT)  PNG: RDT test result |
| g6pdtester | N/A | Indonesia: Staff ID (G6PD Tester)  PNG: ID of staff conducting G6PD test |
| g6pd | U/g Hb | G6PD |
| g6pdoor | N/A | G6PD Out of Measuring Range |
| hbhc | U/g Hb | Hb (Hemocue) |
| hbhcoor | N/A | Hb Hemocue Out of Measuring Range |
| hb | g/dL | Hb |
| hboor | N/A | Hb Out of Measuring Range |
| g6pdcat | N/A | G6PD Category |
| prescriber | N/A | Indonesia: Staff ID (Prescriber)  PNG: ID number of prescriber |
| pqduration | N/A | PQ Treatment Duration |
| pq | Tablets | PQ (total tablets) |
| pqobs | N/A | Indonesia: 1^st^ dose PQ observed?  PNG: First dose PQ taken at facility |
| pqfood | N/A | PQ taken with food? |
| pqreas | N/A | Indonesia: Reason 1^st^ dose PQ not observed  PNG: Reason 1^st^ dose PQ not taken |
| pqreasoth | N/A | Indonesia: Other reason 1^st^ dose PQ not observed  PNG: Other reason 1^st^ dose PQ not taken |
| dhp | DHP (total tablets) | DHP (total tablets) |
| dhpobs | 1^st^ dose DHP observed? | 1^st^ dose DHP observed? |
| dhpreas | Reason 1^st^ dose DHP not observed | Reason 1^st^ dose DHP not observed |
| dhpreasoth | Other reason 1^st^ dose DHP not observed | Other reason 1^st^ dose DHP not observed |
| al | AL (total tablets) | AL (total tablets) |
| txother | Other Treatment | Other Treatment |
| comments | Comments | Comments |
|  |  | **Field Label (en)** |
| **Level 2 Review CRF** | | |
| *scopeid* | *N/A* | *SCOPE Study ID* |
| *studysite* | *N/A* | *Study Site* |
| l2_reviewdat | N/A | Date of Review |
| l2_reviewday | N/A | Day of Review |
| l2_reviewdayoth | N/A | Day of Review (other) |
| l2_location | N/A | Location of Assessment |
| l2_locationoth | N/A | Location of Assessment (Other) |
| l2_reviewer | N/A | Indonesia: Staff Identification Number  PNG: Staff performing Level 2 Review |
| *initials* | *N/A* | *Patient Initials* |
| *mcard* | *N/A* | *Indonesia: Malaria Card Number*  *PNG: Malaria ID* |
| *sex* | *N/A* | *Sex* |
| *age* | *years* | *Age (years)* |
| *agemonths* | *months* | *Age (months)* |
| *pqduration* | *N/A* | *Initial Treatment* |
| l2_tabs | N/A | Yesterday, did you take your malaria tablets? |
| l2_tabsfood | N/A | If Yes, did you take them with food? |
| l2_temp | °C | Temperature |
| l2_pulse | beats per minute | Pulse |
| l2_resp | breaths per minute | Respiratory Rate |
| l2_abdosev | N/A | Abdominal pain - Severity |
| l2_abdodat | N/A | Abdominal pain - Date of Onset |
| l2_nauseasev | N/A | Nausea - Severity |
| l2_nauseadat | N/A | Nausea - Date of Onset |
| l2_eatsev | N/A | Unable to eat - Severity |
| l2_eatdat | N/A | Unable to eat - Date of Onset |
| l2_vomitsev | N/A | Vomiting - Severity |
| l2_vomitdat | N/A | Vomiting - Date of Onset |
| l2_backpainsev | N/A | Back pain - Severity |
| l2_backpaindat | N/A | Back pain - Date of Onset |
| l2_breathsev | N/A | Breathlessness - Severity |
| l2_breathdat | N/A | Breathlessness - Date of Onset |
| l2_dizzysev | N/A | Dizziness - Severity |
| l2_dizzydat | N/A | Dizziness - Date of Onset |
| l2_fatigue | N/A | Fatigue |
| l2_fatiguedat | N/A | Fatigue - Date of Onset |
| l2_vomit2 | N/A | Vomiting > 2 x per day |
| l2_vomit2dat | N/A | Vomiting > 2 x per day - Date of Onset |
| l2_jaund | N/A | Jaundice |
| l2_jaunddat | N/A | Jaundice - Date of Onset |
| l2_blue | N/A | Blue lips |
| l2_bluedat | N/A | Blue lips - Date of Onset |
| l2_dkurine | N/A | Dark urine |
| l2_dkurinedat | N/A | Dark urine - Date of Onset |
| l2_othsymp | N/A | Other symptom(s) |
| l2_othsympdat | N/A | Other symptom(s) - Date of Onset |
| *g6pd* | *U/g Hb* | *G6PD* |
| *g6pdoor* | *N/A* | *G6PD Out of Measuring Range* |
| *hb* | *g/dL* | *Hb* |
| *hboor* | *N/A* | *Hb Out of Measuring Range* |
| l2_g6pdtester | N/A | Indonesia: Staff ID (G6PD Tester)  PNG: ID of staff conducting G6PD test |
| l2_hbhcdat | N/A | Hemocue Date (today) |
| l2_hbhc | g/dL | Hemocue Hb (today) |
| l2_hbhcoor | N/A | Hemocue Hb Out of Measuring Range |
| l2_chghbhc | g/dL | Hemocue Change in Hb (Calculated) |
| l2_changehbhc | g/dL | Hemocue Change in Hb |
| l2_g6pddat | N/A | Biosensor Date (today) |
| l2_g6pd | U/g Hb | G6PD (today) |
| l2_g6pdoor | N/A | G6PD Out of Measuring Range |
| l2_hb | g/dL | Hb (today) |
| l2_hboor | N/A | Hb Out of Measuring Range |
| l2_changehb | g/dL | Change in Hb (Calculated) |
| l2_chghb | g/dL | Change in Hb |
| l2_methbdat | N/A | MetHb Date |
| l2_methb | % | MetHb % |
| l2_needref | N/A | Need for Referral? |
| l2_pq | N/A | Primaquine |
| l2_pqdose | mg | PQ Daily Dose |
| l2_pqday | N/A | Frequency of dosing |
| l2_pqduration | days | PQ Duration |
| l2_refl3 | N/A | Referred for Level 3 Review |
| l2_refl3dat | N/A | Level 3 Referral Date |
| l2_refl3tim | N/A | Level 3 Referral Time |
| l2_l3conf | N/A | Level 3 Confirmation |
| l2_l3confdat | N/A | Level 3 Confirmation Date |
| l2_l3conftim | N/A | Level 3 Confirmation Time |
| l2_whatsapp | N/A | Whatsapp Group |
| l2_whatsappdat | N/A | Whatsapp Group Date |
| l2_education | N/A | Education Given |
| l2_comments | N/A | Comments |
| **Level 3 Review CRF** | | |
| *scopeid* | *SCOPE Study ID* | *N/A* |
| *studysite* | *Study Site* | *N/A* |
| l3_location | Location | N/A |
| l3_locationoth | Location (Other) | N/A |
| l3_reptype | Report Type | N/A |
| l3_fupno | Follow-up Number | N/A |
| l3_reviewdat | Date of Level 3 Review | N/A |
| l3_reviewtim | Time of Level 3 Review | N/A |
| l3_reviewer | Indonesia: Staff Identification Number  PNG: Staff performing Level 3 Review | N/A |
| *initials* | *Patient Initials* | *N/A* |
| *mcard* | *Indonesia: Malaria Card Number*  *PNG: Malaria ID* | *N/A* |
| *sex* | *Sex* | *N/A* |
| *age* | *Age (years)* | *years* |
| *agemonths* | *Age (months)* | *months* |
| *pqduration* | *Initial Treatment* | *N/A* |
| l3_referdat | Date referred | N/A |
| l3_temp | Temperature | °C |
| l3_pulse | Pulse | beats per minute |
| l3_resp | Respiratory Rate | breaths per minute |
| l3_abdosev | Abdominal pain - Severity | N/A |
| l3_abdodat | Abdominal pain - Date of Onset | N/A |
| l3_nauseasev | Nausea - Severity | N/A |
| l3_nauseadat | Nausea - Date of Onset | N/A |
| l3_eatsev | Unable to eat - Severity | N/A |
| l3_eatdat | Unable to eat - Date of Onset | N/A |
| l3_vomitsev | Vomiting - Severity | N/A |
| l3_vomitdat | Vomiting - Date of Onset | N/A |
| l3_backpainsev | Back pain - Severity | N/A |
| l3_backpaindat | Back pain - Date of Onset | N/A |
| l3_breathsev | Breathlessness - Severity | N/A |
| l3_breathdat | Breathlessness - Date of Onset | N/A |
| l3_dizzysev | Dizziness - Severity | N/A |
| l3_dizzydat | Dizziness - Date of Onset | N/A |
| l3_fatiguesev | Fatigue - Severity | N/A |
| l3_fatiguedat | Fatigue - Date of Onset | N/A |
| l3_fever | Fever | N/A |
| l3_feverdat | Fever - Date of Onset | N/A |
| l3_pallor | Severe pallor | N/A |
| l3_pallordat | Severe pallor - Date of Onset | N/A |
| l3_jaund | Jaundice | N/A |
| l3_jaunddat | Jaundice - Date of Onset | N/A |
| l3_blue | Blue lips | N/A |
| l3_bluedat | Blue lips - Date of Onset | N/A |
| l3_dkurine | Dark urine | N/A |
| l3_dkurinedat | Dark urine - Date of Onset | N/A |
| l3_hillmen | Indonesia: Dark urine colour (Hillmen)  PNG: Hillmen Colour Chart #: | N/A |
| l3_othsymp | Other symptom(s) | N/A |
| l3_othsympdat | Other symptom(s) - Date of Onset | N/A |
| l3_narrativecp | Narrative | N/A |
| *hb* | *Hb* | *g/dL* |
| *hboor* | *Hb Out of Measuring Range* | *N/A* |
| l3_g6pdtester | Indonesia: Staff ID (G6PD Tester)  PNG: ID of staff conducting G6PD test | N/A |
| l3_hbhcdat | Hemocue Hb Date (Today) | N/A |
| l3_hbhc | Hemocue Hb (Today) | g/dL |
| l3_hbhcoor | Hemocue Hb Out of Measuring Hemocue Range | N/A |
| l3_hbhcmethod | Hemocue Hb Method (Today) | N/A |
| l3_nadhbhc | Hemocue Hb (Nadir) (Calculated) | g/dL |
| l3_nadhbhcoor | Hemocue Hb Out of Measuring Range (Nadir) (Calculated) | N/A |
| l3_hbhcmaxfall | Hemocue Max Fall Hb (Calculated) | g/dL |
| l3_hbhcpcfall | Hemocue Max % Fall Hb (Calculated) | % |
| l3_hbdat | Hb Date (Today) | N/A |
| l3_hb | Hb (Today) | g/dL |
| l3_hboor | Hb Out of Measuring Range | N/A |
| l3_hbmethod | Hb Method (Today) | N/A |
| l3_nadhb | Hb (Nadir) (Calculated) | g/dL |
| l3_nadhboor | Hb Out of Measuring Range (Nadir) (Calculated) | N/A |
| l3_hbmaxfall | Max Fall Hb (Calculated) | g/dL |
| l3_hbpcfall | Max % Fall Hb (Calculated) | % |
| *g6pd* | *G6PD* | *U/g Hb* |
| *g6pdoor* | *G6PD Out of Measuring Range* | *N/A* |
| l3_methbdat | MetHb Date | N/A |
| l3_methb | MetHb % | % |
| l3_maxgrade | Max Graded Symptom | N/A |
| l3_maxgradedat | Max Graded Symptom – Date of Onset | N/A |
| l3_outcome | AE Outcome | N/A |
| l3_aesihaem | AESI Haemolysis | N/A |
| l3_aesihaemdat | AESI Haemolysis – Date of Onset | N/A |
| l3_aesihaemflag | AESI Haemolysis – Flags | N/A |
| l3_aesigast | AESI Gastrointestinal | N/A |
| l3_aesigastdat | AESI Gastrointestinal – Date of Onset | N/A |
| l3_aesigastflag | AESI Gastrointestinal – Flags | N/A |
| l3_aesimethb | AESI MetHb | N/A |
| l3_aesimethbdat | AESI MetHb – Date of Onset | N/A |
| l3_aesimethbflag | AESI MetHb – Flags | N/A |
| l3_sae | SAE | N/A |
| l3_saetype | SAE Type | N/A |
| l3_saetypeoth | Other SAE Type | N/A |
| l3_pqchange | Changes to PQ | N/A |
| l3_pqdose | PQ Daily Dose | mg |
| l3_pqday | Frequency of dosing | N/A |
| l3_pqduration | PQ Duration | days |
| l3_narrativeaesi | Narrative for AESI | N/A |
| l3_review | Planned Review | N/A |
| l3_reviewrtn | Planned Review Routine | N/A |
| l3_reviewoth | Other follow-up Day | N/A |
| l3_reviewothdat | Other follow-up Date | N/A |
| l3_refhospdat | Referred to Hospital Date | N/A |
| l3_refhosptim | Referred to Hospital Time | N/A |
| l3_notify | Notification | N/A |
| l3_comments | Comments | N/A |
| **Serious Adverse Event CRF** | | |
| *scopeid* | *SCOPE Study ID* | *N/A* |
| *studysite* | *Study Site* | *N/A* |
| sae_reviewdat | Date of Assessment | N/A |
| sae_reviewtim | Time of Assessment | N/A |
| sae_reptype | Report Type | N/A |
| sae_fupno | Follow-up Number | N/A |
| sae_location | Location of Assessment | N/A |
| *initials* | *Patient Initials* | *N/A* |
| *mcard* | *Indonesia: Malaria Card Number*  *PNG: Malaria ID* | *N/A* |
| *sex* | *Sex* | *N/A* |
| *age* | *Age (years)* | *years* |
| *agemonths* | *Age (months)* | *months* |
| *weight* | *Weight* | *kg* |
| *country* | *Country* | *N/A* |
| sae_pqdoses | Number of PQ doses taken before AE detected | N/A |
| sae_pqfood | Did the patient take their last PQ tablets with food? | N/A |
| sae_cm | Concomitant Medication | N/A |
| sae_cmname1 | Concomitant Medication 1 Name | N/A |
| sae_cmstdat1 | Concomitant Medication 1 Start Date | N/A |
| sae_cmenddat1 | Concomitant Medication 1 Stop Date | N/A |
| sae_cmstdose1 | Concomitant Medication 1 Dose | N/A |
| sae_cmfreq1 | Concomitant Medication 1 Frequency | N/A |
| sae_cmname2 | Concomitant Medication 1 Name | N/A |
| sae_cmstdat2 | Concomitant Medication 1 Start Date | N/A |
| sae_cmenddat2 | Concomitant Medication 1 Stop Date | N/A |
| sae_cmstdose2 | Concomitant Medication 1 Dose | N/A |
| sae_cmfreq2 | Concomitant Medication 1 Frequency | N/A |
| sae_cmname3 | Concomitant Medication 1 Name | N/A |
| sae_cmstdat3 | Concomitant Medication 1 Start Date | N/A |
| sae_cmenddat3 | Concomitant Medication 1 Stop Date | N/A |
| sae_cmstdose3 | Concomitant Medication 1 Dose | N/A |
| sae_cmfreq3 | Concomitant Medication 1 Frequency | N/A |
| sae_cmname4 | Concomitant Medication 1 Name | N/A |
| sae_cmstdat4 | Concomitant Medication 1 Start Date | N/A |
| sae_cmenddat4 | Concomitant Medication 1 Stop Date | N/A |
| sae_cmstdose4 | Concomitant Medication 1 Dose | N/A |
| sae_cmfreq4 | Concomitant Medication 1 Frequency | N/A |
| sae_cmname5 | Concomitant Medication 1 Name | N/A |
| sae_cmstdat5 | Concomitant Medication 1 Start Date | N/A |
| sae_cmenddat5 | Concomitant Medication 1 Stop Date | N/A |
| sae_cmstdose5 | Concomitant Medication 1 Dose | N/A |
| sae_cmfreq5 | Concomitant Medication 1 Frequency | N/A |
| sae_cmname6 | Concomitant Medication 1 Name | N/A |
| sae_cmstdat6 | Concomitant Medication 1 Start Date | N/A |
| sae_cmenddat6 | Concomitant Medication 1 Stop Date | N/A |
| sae_cmstdose6 | Concomitant Medication 1 Dose | N/A |
| sae_cmfreq6 | Concomitant Medication 1 Frequency | N/A |
| sae_maindx | AE Main Diagnosis | N/A |
| sae_stdat | Date AE Started | N/A |
| sae_serdat | Date AE met Serious Criteria | N/A |
| sae_detdat | Date SAE detected | N/A |
| sae_hospadmdat | Date of Hospitalisation | N/A |
| sae_hospna | Hospitalisation Not Applicable | N/A |
| sae_hospdisdat | Date of Discharge | N/A |
| sae_hosp | Still in Hospital |  |
| sae_narrativecp | Narrative |  |
| sae_med | Relevant Medical History | N/A |
| sae_medname1 | Medical Condition 1 Name | N/A |
| sae_medstdat1 | Medical Condition 1 Start Date | N/A |
| sae_medenddat1 | Medical Condition 1 End Date | N/A |
| sae_medong1 | Medical Condition 1 Ongoing | N/A |
| sae_medname2 | Medical Condition 2 Name | N/A |
| sae_medstdat2 | Medical Condition 2 Start Date | N/A |
| sae_medenddat2 | Medical Condition 2 End Date | N/A |
| sae_medong2 | Medical Condition 2 Ongoing | N/A |
| sae_medname3 | Medical Condition 3 Name | N/A |
| sae_medstdat3 | Medical Condition 3 Start Date | N/A |
| sae_medenddat3 | Medical Condition 3 End Date | N/A |
| sae_medong3 | Medical Condition 3 Ongoing | N/A |
| sae_medname4 | Medical Condition 4 Name | N/A |
| sae_medstdat4 | Medical Condition 4 Start Date | N/A |
| sae_medenddat4 | Medical Condition 4 End Date | N/A |
| sae_medong4 | Medical Condition 4 Ongoing | N/A |
| sae_hbhcdetdat | Hemocue Hb Date (At time of Detection) | N/A |
| sae_hbhcdet | Hemocue Hb (At time of Detection) | g/dL |
| sae_hbhcdetoor | Hemocue Hb Out of Measuring Range | N/A |
| sae_hbhcdetmeth | Hemocue Hb Method (At time of Detection) | N/A |
| sae_hbhcnaddat | Hemocue Hb Date (Nadir) | N/A |
| sae_hbhcnad | Hemocue Hb (Nadir) | g/dL |
| sae_hbhcnadmeth | Hemocue Hb Method (Nadir) | N/A |
| sae_g6pdtester | Indonesia: Staff ID (G6PD Tester)  PNG: ID of staff conducting G6PD test | N/A |
| sae_hbhc | Hemocue Hb (Current) | g/dL |
| sae_hbhcdat | Hemocue Hb Date (Current) | N/A |
| sae_hbhcmeth | Hemocue Hb Method (Current) | N/A |
| sae_hbhcoor | Hemocue Hb Out of Measuring Range | N/A |
| sae_hbhcmaxfall | Hemocue Max Fall Hb | g/dL |
| sae_hbhcpcfall | Hemocue Max % Fall Hb | % |
| sae_hbdetdat | Hb Date (At time of Detection) | N/A |
| sae_hbdet | Hb (At time of Detection) | g/dL |
| sae_hboor | Hb Out of Measuring Range | N/A |
| sae_hbdetmeth | Hb Method (At time of Detection) | N/A |
| sae_hbnaddat | Hb Date (Nadir) | N/A |
| sae_hbnad | Hb (Nadir) | g/dL |
| sae_hbnadmeth | Hb Method (Nadir) | N/A |
| sae_hbcur | Hb (Current) | g/dL |
| sae_hbcurdat | Hb Date (Current) | N/A |
| sae_hbcurmeth | Hb Method (Current) | N/A |
| sae_hbmaxfall | Max Fall Hb | g/dL |
| sae_hbpcfall | Max % Fall Hb | % |
| sae_curg6pddat | G6PD Date | N/A |
| sae_curg6pd | G6PD Result | U/g Hb |
| sae_g6pdoor | G6PD (Out of Measuring Range) | N/A |
| sae_g6pdcat | G6PD Category | N/A |
| sae_g6pdgen | G6PD Genotyping | N/A |
| sae_g6pdvariant | Genotyping Variant | N/A |
| sae_wbcdat | WBC Date | N/A |
| sae_wbc | WBC Result | x10^3^/µL |
| sae_pltdat | Plt Date | N/A |
| sae_plt | Plt Result | x10^3^/µL |
| sae_methbdat | MetHb Date | N/A |
| sae_methb | MetHb Result | % |
| sae_nadat | Na Date | N/A |
| sae_na | Na Result | mmol/L |
| sae_kdat | K Date | N/A |
| sae_k | K Result | mmol/L |
| sae_ureadat | Urea Date | N/A |
| sae_urea | Urea Result | mg/dL |
| sae_creatdat | Creatinine Date | N/A |
| sae_creat | Creatinine Result | mg/dL |
| sae_totbilidat | Total Bili Date | N/A |
| sae_totbili | Total Bili Result | µmol/L |
| sae_unconjbilidat | Unconj Bili Date | N/A |
| sae_unconjbili | Unconj Bili Result | µmol/L |
| sae_alpdat | ALP Date | N/A |
| sae_alp | ALP Result | µmol/L |
| sae_altdat | ALT Date | N/A |
| sae_alt | ALT Result | µmol/L |
| sae_ldhdat | LDH Date | N/A |
| sae_ldh | LDH Result | µmol/L |
| sae_testoth | Other Results | N/A |
| sae_saetype | SAE Type | N/A |
| sae_organs | Organ Systems Involved | N/A |
| sae_relpq | Relationship (causality) to PQ | N/A |
| sae_iv | IV Fluids | N/A |
| sae_bltr | Blood Transfusion | N/A |
| sae_bltrunits | Blood Transfusion – Number of units | N/A |
| sae_bltrdat | Blood Transfusion – Date | N/A |
| sae_dial | Dialysis | N/A |
| sae_dialtype | Dialysis Type | N/A |
| sae_pqchange | Changes to PQ | N/A |
| sae_pqdat | PQ Date Restarted | N/A |
| sae_pqdose | PQ Daily Dose | mg |
| sae_pqday | Frequency of dosing | N/A |
| sae_pqduration | PQ Duration | days |
| sae_narrativecm | Narrative | N/A |
| sae_outcome | Outcome | N/A |
| sae_rrs | Recovered/Resolved with Sequelae – Specify | N/A |
| sae_dod | Date of Death | N/A |
| sae_cod | Cause of Death | N/A |
| sae_lastdat | AE Stopped – Last date AE was present | N/A |
| sae_notify | Notification | N/A |
| sae_comments | Comments | N/A |

### Appendix 2

**LIST OF TABLES/FIGURES/LISTINGS**

| **Number** | **Title** |
| --- | --- |
| Table 1 | Baseline characteristics of patients enrolled in Stage 1 safety and feasibility study by health facility site |
| Table 2 | Baseline characteristics of patients enrolled in Stage 1 safety and feasibility study by treatment arm |
| Table 3 | Number of events for patients enrolled in Stage 1 safety and feasibility study by health facility site |
| Table 4 | Number of patients with at least one event enrolled in Stage 1 safety and feasibility study by health facility site |
| Table 5 | Number of patients with at least one event enrolled in Stage 1 safety and feasibility study by treatment arm |
| Table 6 | Baseline characteristics of individuals presenting to health facilities by pre-implementation and post-implementation (Stage 2) phases of study |
| Table 7 | Estimates of the effect of the implementation package on the incidence of symptomatic *P. vivax* malaria episodes |
| Table 8 | Baseline characteristics of individuals included in the cross-sectional surveys in the pre-implementation and post-implementation (Stage 2) phases of study |
| Table 9 | Estimates of the effect of the implementation package on the prevalence of *P. vivax* parasitaemia and severe anaemia (haemoglobin < 7g/dl) in patients presenting with fever in the cross-sectional surveys in the pre-implementation and post-implementation (Stage 2) phases of study |
| Table 10 | Descriptive statistics and 95% limits of agreement for haemoglobin measurements obtained by using Biosensor and HemoCue, stratified by day of measurement (day 0, 3, 7) and country |
| Table S1 | Protocol deviations for patients enrolled in Stage 1 safety and feasibility study by health facility site |
| Table S2 | Baseline characteristics of individuals presenting to health facilities by pre-implementation and post-implementation (Stage 2) phases of study for each health facility site |
| Table S3 | Estimates of the effect of the implementation package on the incidence of symptomatic *P. vivax* malaria episodes – alternate seasonality specification (adjustment for rainy/dry season) |
| Figure 1 | Study overview |
| Figure 2 | Incidence of symptomatic *P. vivax* malaria episodes over time. The introduction of the implementation package at 6 months is indicated by the dashed red vertical line. The trend lines for the pre- and post-implementation stages are presented as solid blue lines, and the counterfactual to the post-implementation trend presented as a dashed blue line.  *[Note: the example figure is given for a single health facility site]* |
| Figure 3 | Bland–Altman plots comparing the measurements haemoglobin obtained using HemoCue and SD Biosensor. |

### Appendix 3 – Dummy tables and figures

**Table 1. Baseline characteristics of patients enrolled in Stage 1 safety and feasibility study by health facility site**

|  | **Overall** | **HF1** | **HF2** | **HF3** | **HF4** |
| --- | --- | --- | --- | --- | --- |
|  | N= | N= | N= | N= |  |
| **Age (mean and range)** |  |  |  |  |  |
| <5yrs |  |  |  |  |  |
| 5-15 |  |  |  |  |  |
| >15yrs |  |  |  |  |  |
| Unknown |  |  |  |  |  |
| **Sex n (%)** |  |  |  |  |  |
| Male |  |  |  |  |  |
| Female |  |  |  |  |  |
| Unknown |  |  |  |  |  |
| **Malaria diagnosis n (%)** |  |  |  |  |  |
| Pv mono infection (microscopy) |  |  |  |  |  |
| Pv mixed infection (microscopy) |  |  |  |  |  |
| Pv mono/mixed (RDT) |  |  |  |  |  |
| Total Pv (mono or mixed) |  |  |  |  |  |
| Unknown |  |  |  |  |  |
| **G6PD value** |  |  |  |  |  |
| Normal |  |  |  |  |  |
| Intermediate All |  |  |  |  |  |
| Intermediate Females |  |  |  |  |  |
| Intermediate Males |  |  |  |  |  |
| Deficient |  |  |  |  |  |
| Unknown |  |  |  |  |  |
| **Treatment** |  |  |  |  |  |
| PQ7 |  |  |  |  |  |
| PQ14 |  |  |  |  |  |
| PQ8W |  |  |  |  |  |
| None |  |  |  |  |  |
| Unknown |  |  |  |  |  |

**Table 2. Baseline characteristics of patients enrolled in Stage 1 safety and feasibility study by treatment arm**

|  | **Overall** | **PQ7** | **PQ14** | **PQ8W** |
| --- | --- | --- | --- | --- |
|  | N= | N= | N= | N= |
| **Age (mean and range)** |  |  |  |  |
| <5yrs |  |  |  |  |
| 5-15 |  |  |  |  |
| >15yrs |  |  |  |  |
| **Sex n (%)** |  |  |  |  |
| Male |  |  |  |  |
| Female |  |  |  |  |
| Unknown |  |  |  |  |
| **Malaria diagnosis n (%)** |  |  |  |  |
| Pv mono infection (microscopy) |  |  |  |  |
| Pv mixed infection (microscopy) |  |  |  |  |
| Pv mono/mixed (RDT) |  |  |  |  |
| Total Pv (mono or mixed) |  |  |  |  |
| Unknown |  |  |  |  |

**Table 3. Number of events for patients enrolled in Stage 1 safety and feasibility study by health facility site**

|  | **Overall** | **HF1** | **HF2** | **HF3** | **HF4** |
| --- | --- | --- | --- | --- | --- |
| **Total Enrolled N** |  |  |  |  |  |
| Flagged Day 3-5 N(%) * |  |  |  |  |  |
| Flagged Day 6-10 N(%) * |  |  |  |  |  |
| Other Day Flagged N(%) * |  |  |  |  |  |
| **AEs n(%)** |  |  |  |  |  |
| Total Grade 3 |  |  |  |  |  |
| Total Grade 4 |  |  |  |  |  |
| **Total** |  |  |  |  |  |
| **AESI** |  |  |  |  |  |
| **Haemolysis** |  |  |  |  |  |
| AESI (SAE) |  |  |  |  |  |
| AESI (NonSAE) |  |  |  |  |  |
| **Total** |  |  |  |  |  |
| **Gastrointestinal** |  |  |  |  |  |
| AESI (SAE) |  |  |  |  |  |
| AESI (NonSAE) |  |  |  |  |  |
| **Total** |  |  |  |  |  |
| **Methaemoglobinaemia** |  |  |  |  |  |
| AESI (SAE) |  |  |  |  |  |
| AESI (NonSAE) |  |  |  |  |  |
| **Total** |  |  |  |  |  |
| **SAEs** |  |  |  |  |  |
| SAE – definitely related |  |  |  |  |  |
| SAE – probably related |  |  |  |  |  |
| SAE – possibly related |  |  |  |  |  |
| SAE – unlikely related |  |  |  |  |  |
| SAE – not related |  |  |  |  |  |
| **Total** |  |  |  |  |  |
| Related SAE which is also an AESI |  |  |  |  |  |
| Related SAE Other |  |  |  |  |  |

**Table 4. Number of patients with at least one event who are enrolled in Stage 1 safety and feasibility study by health facility site**

|  | **Overall** | **HF1** | **HF2** | **HF3** | **HF4** |
| --- | --- | --- | --- | --- | --- |
| **Total Enrolled N** |  |  |  |  |  |
| Flagged Day 3-5 N(%) * |  |  |  |  |  |
| Flagged Day 6-10 N(%) * |  |  |  |  |  |
| Other Day Flagged N(%) * |  |  |  |  |  |
| **Aes n(%)** |  |  |  |  |  |
| Total Grade 3 |  |  |  |  |  |
| Total Grade 4 |  |  |  |  |  |
| **Total** |  |  |  |  |  |
| **AESI** |  |  |  |  |  |
| **Haemolysis** |  |  |  |  |  |
| AESI (SAE) |  |  |  |  |  |
| AESI (NonSAE) |  |  |  |  |  |
| **Total** |  |  |  |  |  |
| **Gastrointestinal** |  |  |  |  |  |
| AESI (SAE) |  |  |  |  |  |
| AESI (NonSAE) |  |  |  |  |  |
| **Total** |  |  |  |  |  |
| **Methaemoglobinaemia** |  |  |  |  |  |
| AESI (SAE) |  |  |  |  |  |
| AESI (NonSAE) |  |  |  |  |  |
| **Total** |  |  |  |  |  |
| **SAEs** |  |  |  |  |  |
| SAE – definitely related |  |  |  |  |  |
| SAE – probably related |  |  |  |  |  |
| SAE – possibly related |  |  |  |  |  |
| SAE – unlikely related |  |  |  |  |  |
| SAE – not related |  |  |  |  |  |
| **Total** |  |  |  |  |  |
| Related SAE which is also an AESI |  |  |  |  |  |
| Related SAE Other |  |  |  |  |  |

**Table 5. Number of patients with at least one event enrolled in Stage 1 safety and feasibility study by treatment arm**

|  | **Overall** | **PQ7** | **PQ14** | **PQ8W** |
| --- | --- | --- | --- | --- |
| **Total Enrolled N** |  |  |  |  |
| Flagged Day 3-5 N(%) * |  |  |  |  |
| Flagged Day 6-10 N(%) * |  |  |  |  |
| Other Day Flagged N(%) * |  |  |  |  |
| **Aes n(%)** |  |  |  |  |
| Total Grade 3 |  |  |  |  |
| Total Grade 4 |  |  |  |  |
| **Total** |  |  |  |  |
| **AESI** |  |  |  |  |
| **Haemolysis** |  |  |  |  |
| AESI (SAE) |  |  |  |  |
| AESI (NonSAE) |  |  |  |  |
| **Total** |  |  |  |  |
| **Gastrointestinal** |  |  |  |  |
| AESI (SAE) |  |  |  |  |
| AESI (NonSAE) |  |  |  |  |
| **Total** |  |  |  |  |
| **Methaemoglobinaemia** |  |  |  |  |
| AESI (SAE) |  |  |  |  |
| AESI (NonSAE) |  |  |  |  |
| **Total** |  |  |  |  |
| **SAEs** |  |  |  |  |
| SAE – definitely related |  |  |  |  |
| SAE – probably related |  |  |  |  |
| SAE – possibly related |  |  |  |  |
| SAE – unlikely related |  |  |  |  |
| SAE – not related |  |  |  |  |
| **Total** |  |  |  |  |
| Related SAE which is also an AESI |  |  |  |  |
| Related SAE Other |  |  |  |  |

**Table 6. Baseline characteristics of individuals presenting to health facilities by pre-implementation and post-implementation (Stage 2) phases of study**

|  | Pre-implementation | Post-implementation (Stage 2) |
| --- | --- | --- |
| **Number of individuals** | N | N |
| **Number of individuals per health facility site** | x (x, x) [min, max] | x (x, x) [min, max] |
| **Age, years**  **(mean and range)** |  |  |
| <5yrs |  |  |
| 5-15 |  |  |
| >15yrs |  |  |
| Unknown |  |  |
| **Sex n (%)** |  |  |
| Male |  |  |
| Female |  |  |
| Unknown |  |  |
| **Malaria diagnosis n (%)** |  |  |
| Pv mono infection (microscopy) |  |  |
| Pv mixed infection (microscopy) |  |  |
| Pv mono/mixed (RDT) |  |  |
| Total Pv (mono or mixed) |  |  |
| Unknown |  |  |
| **G6PD value** |  |  |
| Normal |  |  |
| Intermediate All |  |  |
| Intermediate Females |  |  |
| Intermediate Males |  |  |
| Deficient |  |  |
| Unknown |  |  |
| **Treatment** |  |  |
| PQ7 |  |  |
| PQ14 |  |  |
| PQ8W |  |  |
| None |  |  |
| Unknown |  |  |

N = Total number; x (x, x) = median (lower quartile, upper quartile).

**Table 7. Estimates of the effect of the implementation package on the incidence of symptomatic *P. vivax* malaria episodes**

|  | **Pre-implementation** | **Post-implementation** | **Post- versus pre-implementation** |
| --- | --- | --- | --- |
| **Effect of intervention 6 months after date of implementation** | Est of IR (lb, ub) | Est of IR (lb, ub) | Est of IRR (lb, ub) |
| **Effect of intervention on slope after date of implementation** | Est of change in IR per month (lb, ub) | Est of change in IR per month (lb, ub) | Est of slope change (lb, ub) |

Est (lb, ub) = Estimate (lower bound of 95% CI, upper bound of 95% CI), IR – Incidence Rate, IRR – Incidence Rate Ratio

**Table 8. Baseline characteristics of individuals included in the cross-sectional surveys in the pre-implementation and post-implementation (Stage 2) phases of study**

|  | Pre-implementation survey | Post-implementation (Stage 2) survey |
| --- | --- | --- |
| **Number of individuals** | N | N |
| **Number of individuals per site** | x (x, x) [min, max] | x (x, x) [min, max] |
| **Age, years**  **(mean and range)** |  |  |
| <5yrs |  |  |
| 5-15 |  |  |
| >15yrs |  |  |
| Unknown |  |  |
| **Sex n (%)** |  |  |
| Male |  |  |
| Female |  |  |
| Unknown |  |  |
| **Malaria diagnostic method n (%)** |  |  |
| Film |  |  |
| RDT |  |  |
| Both |  |  |
| **Malaria diagnosis n (%)** |  |  |
| Pv |  |  |
| Pf |  |  |
| Po |  |  |
| Pm |  |  |
| Mixed species |  |  |
| Unknown |  |  |
| **Parasitaemia (geometric mean and range)** |  |  |
| Pv |  |  |
| Pf |  |  |
| Po |  |  |
| Pm |  |  |
| Mixed species |  |  |
| **Gametocytaemia n (%)** |  |  |
| Pv |  |  |
| Pf |  |  |
| Po |  |  |
| Pm |  |  |
| Mixed species |  |  |
| **G6PD value** |  |  |
| Normal |  |  |
| Intermediate All |  |  |
| Intermediate Females |  |  |
| Intermediate Males |  |  |
| Deficient |  |  |
| Unknown |  |  |
| **Haemoglobin (median, IQR, range)** |  |  |
| **Anaemia <7g/dL (n (%)** |  |  |

**Table 9. Estimates of the effect of the implementation package on the prevalence of *P. vivax* parasitaemia and severe anaemia (haemoglobin < 7g/dl) in patients presenting with fever in the cross-sectional surveys in the pre-implementation and post-implementation (Stage 2) phases of study**

|  | **Pre-implementation** | **Post-implementation** | **Post- versus pre-implementation** |
| --- | --- | --- | --- |
| ***P. vivax* parasitaemia** | Est of P (lb, ub) | Est of P (lb, ub) | Est of PR (lb, ub) |
| **Severe anaemia** | Est of P (lb, ub) | Est of P (lb, ub) | Est of PR (lb, ub) |

Est (lb, ub) = Estimate (lower bound of 95% CI, upper bound of 95% CI), P – Prevalence, IRR – Prevalence Ratio

**Table 10. Descriptive statistics and 95% limits of agreement for haemoglobin measurements obtained by using Biosensor and HemoCue, stratified by day of measurement (day 0, 3, 7) and country**

|  | **HemoCue haemoglobin (g/dL)** | **SD Biosensor haemoglobin (g/dL)** | **Difference between two measurements (g/dL)** | **95% limits of agreement (g/dL)** |
| --- | --- | --- | --- | --- |
|  | **Mean (SD); N** | **Mean (SD); N** | **Mean (SD) [95% CI]** |  |
| **Indonesia** |  |  |  |  |
| Day 0 |  |  |  |  |
| Day 3 |  |  |  |  |
| Day 7 |  |  |  |  |
| **PNG** |  |  |  |  |
| Day 0 |  |  |  |  |
| Day 3 |  |  |  |  |
| Day 7 |  |  |  |  |

**Table S1. Protocol deviations for patients enrolled in Stage 1 safety and feasibility study by site**

|  | **Overall** | **HF1** | **HF2** | **HF3** | **HF4** |
| --- | --- | --- | --- | --- | --- |
|  | N= | N= | N= | N= | N= |
| **PRESCRIBING DEVIATIONS** |  |  |  |  |  |
| **Pregnancy n (% of all female patients)** |  |  |  |  |  |
| Treated with PQ |  |  |  |  |  |
| **Breast Feeding infants (<6 months Indonesia, <12 months PNG) n (% of all female patients)** |  |  |  |  |  |
| Treated with PQ |  |  |  |  |  |
| **Age <1year (PNG), <6 months (Indonesia) n (% of all enrolled pts)** |  |  |  |  |  |
| Treated with PQ |  |  |  |  |  |
| **Patients with Pf (Mono) n (% of all enrolled pts)** |  |  |  |  |  |
| Treated with PQ |  |  |  |  |  |
| **G6PD normal >70% N(% of all enrolled pts)** |  |  |  |  |  |
| Treated with PQ14 n(%) |  |  |  |  |  |
| Treated with PQ8W n(%) |  |  |  |  |  |
| **G6PD Intermediate 30-70% N(% of all enrolled pts)** |  |  |  |  |  |
| Treated with PQ7 n(%) |  |  |  |  |  |
| Treated with PQ8W n(%) |  |  |  |  |  |
| **G6PD Deficient <30% N(% of all enrolled pts)** |  |  |  |  |  |
| Treated with PQ7 n(%) |  |  |  |  |  |
| Treated with PQ14 n(%) |  |  |  |  |  |
| **DOSING DEVIATIONS** |  |  |  |  |  |
| **PQ7 n(%)** |  |  |  |  |  |
| daily dose > 1.3mg/kg |  |  |  |  |  |
| daily dose < 0.5mg/kg |  |  |  |  |  |
| **PQ14 n(%)** |  |  |  |  |  |
| daily dose > 0.83mg/kg |  |  |  |  |  |
| daily dose < 0.25mg/kg |  |  |  |  |  |
| **PQ8W n(%)** |  |  |  |  |  |
| daily dose > 1.25mg/kg |  |  |  |  |  |
| daily dose < 0.38mg/kg |  |  |  |  |  |
| **FOLLOW UP DEVIATIONS** |  |  |  |  |  |
| Proportion of Patients without Day3 Review (Day 3-5) N(% **of all enrolled pts**) |  |  |  |  |  |
| Proportion of Patients without Day7 Review (Day 6-10) N(% **of all enrolled pts**) |  |  |  |  |  |

**Table S2. Baseline characteristics of individuals presenting to health facilities by pre-implementation and post-implementation (Stage 2) phases of study for each health facility site**

*Example table give for 2 health facility sites from PNG*

|  | **PNG health facility site 1** |  | **PNG health facility site 2** |  |
| --- | --- | --- | --- | --- |
|  | Pre-implementation | Post-implementation  (Stage 2) | Pre-implementation | Post-implementation  (Stage 2) |
| **Number of individuals** | N | N | N | N |
| **Number of individuals per health facility site** | x (x, x) [min, max] | x (x, x) [min, max] | x (x, x) [min, max] | x (x, x) [min, max] |
| **Age, years**  **(mean and range)** |  |  |  |  |
| <5yrs |  |  |  |  |
| 5-15 |  |  |  |  |
| >15yrs |  |  |  |  |
| Unknown |  |  |  |  |
| **Sex n (%)** |  |  |  |  |
| Male |  |  |  |  |
| Female |  |  |  |  |
| Unknown |  |  |  |  |
| **Malaria diagnosis n (%)** |  |  |  |  |
| Pv mono infection (microscopy) |  |  |  |  |
| Pv mixed infection (microscopy) |  |  |  |  |
| Pv mono/mixed (RDT) |  |  |  |  |
| Total Pv (mono or mixed) |  |  |  |  |
| Unknown |  |  |  |  |
| **G6PD value** |  |  |  |  |
| Normal |  |  |  |  |
| Intermediate All |  |  |  |  |
| Intermediate Females |  |  |  |  |
| Intermediate Males |  |  |  |  |
| Deficient |  |  |  |  |
| Unknown |  |  |  |  |
| **Treatment** |  |  |  |  |
| PQ7 |  |  |  |  |
| PQ14 |  |  |  |  |
| PQ8W |  |  |  |  |
| None |  |  |  |  |
| Unknown |  |  |  |  |

**Table S3. Estimates of the effect of the implementation package on the incidence of symptomatic *P. vivax* malaria episodes (adjusted for seasonality – rainy / dry season)**

|  | **Pre-implementation** | **Post-implementation** | **Post- versus pre-implementation** |
| --- | --- | --- | --- |
| **Effect of intervention 6 months after date of implementation** | Est of IR (lb, ub) | Est of IR (lb, ub) | Est of IRR (lb, ub) |
| **Effect of intervention on slope after date of implementation** | Est of change in IR per month (lb, ub) | Est of change in IR per month (lb, ub) | Est of slope change (lb, ub) |

Est (lb, ub) = Estimate (lower bound of 95% CI, upper bound of 95% CI), IR – Incidence Rate, IRR – Incidence Rate Ratio

**Figure 2. Incidence of symptomatic *P. vivax* malaria episodes over time. The introduction of the implementation package occurs at 6 months (dashed red vertical line). The trend lines for the pre- and post-implementation stages are presented as solid blue lines, and the counterfactual to the post-implementation trend presented as a dashed blue line.**

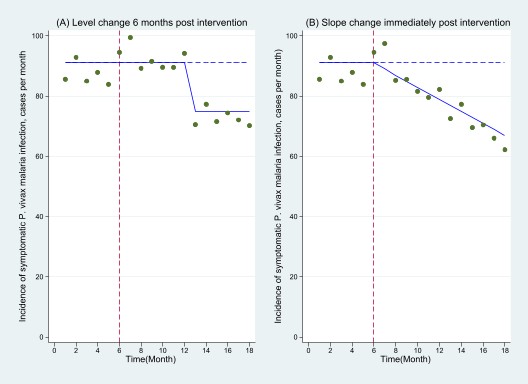
